# Antimicrobial resistance surveillance: can we estimate resistance in bloodstream infections from other types of specimen?

**DOI:** 10.1101/2020.10.12.20211243

**Authors:** Karina-Doris Vihta, Nicola Claire Gordon, Nicole Stoesser, T. Phuong Quan, Carina SB Tyrrell, Manivanh Vongsouvath, Elizabeth A Ashley, Vilada Chansamouth, Paul Turner, Clare L Ling, David Eyre, Nicholas J White, Derrick Crook, Tim Peto, Ann Sarah Walker

**Affiliations:** University of Oxford, Nuffield Department of Clinical Medicine, Oxford, United Kingdom; London School of Hygiene and Tropical Medicine; University of Cambridge, Cambridge, United Kingdom; Lao-Oxford-Mahosot Hospital-Wellcome Trust Research Unit, Vientiane, Laos; Centre for Tropical Medicine & Global Health, Nuffield Department of Medicine, University of Oxford, Oxford, UK; Cambodia Oxford Medical Research Unit, Angkor Hospital for Children, Siem Reap, Cambodia; Shoklo Malaria Research Unit, Mahidol-Oxford Tropical Medicine Research Unit, Faculty of Tropical Medicine, Mahidol University, Mae Sot, Thailand; Mahidol Oxford Tropical Medicine Research Unit, Faculty of Tropical Medicine, Mahidol University, Bangkok, Thailand; Oxford National Institute for Health Research Health Protection Research Unit, Oxford, United Kingdom; University Hospitals Birmingham NHS Foundation Trust, Birmingham, United Kingdom

**Author notes:** Corresponding author: Karina-Doris Vihta, Microbiology Level 7, John Radcliffe Hospital, Headley Way, Oxford, OX3 9DU.

## Abstract

**Background:** Antimicrobial resistance (AMR) surveillance of bloodstream infections is challenging in low- and middle-income countries (LMICs), limited laboratory capacity preventing routine patient-level susceptibility testing. Other specimen types could provide an effective approach to surveillance.

**Objectives:** Our study aims to systematically evaluate the relationship between resistance prevalence in non-sterile sites and bloodstream infections.

**Methods:** Associations between resistance rates in *Escherichia coli* and *Staphylococcus aureus* isolates from blood and other specimens were estimated in Oxfordshire, UK, 1998-2018, comparing proportions resistant in each calendar year using time series cross-correlations and across drug-years. We repeated analysis across publicly-available data from four high-income and 12 middle-income countries, and in three hospitals/programmes in LMICs.

**Results:** 8102 *E. coli* bloodstream infections, 322087 *E. coli* urinary tract infections, 6952 *S. aureus* bloodstream infections and 112074 *S. aureus* non-sterile site cultures were included from Oxfordshire. Resistance trends over time in blood versus other specimens were strongly correlated (maximum cross-correlation 0.51-0.99, strongest associations in the same year for 18/27 pathogen-drug combinations). Resistance prevalence was broadly congruent across drug-years for each species. 276/312 (88%) species-drug-years had resistance prevalence in other specimen types within ±10% of that blood isolates. Results were similar across multiple countries and hospitals/programmes in high/middle/low income-settings.

**Conclusions:** Resistance in bloodstream and less invasive infections are strongly related over time, suggesting the latter could be a surveillance tool for AMR in LMICs. These infection sites are easier to sample and cheaper to obtain the necessary numbers of susceptibility tests, providing more cost-effective evidence for decisions including empiric antibiotic recommendations.

## Introduction

Antimicrobial resistance (AMR) is among the top ten global health threats,^1^ and is particularly acute in low-to middle-income countries (LMICs).^2–4^ Surveillance is a key tool to combat rising AMR, as highlighted by the World Health Organization (WHO)^5^ and formalised in its Global Antimicrobial Resistance Surveillance System (GLASS),^6,7^ particularly in LMICs where lack of laboratory capacity prevents routine patient-level antimicrobial susceptibility testing.^8^ The lack of surveillance data was identified as a key contributor to global AMR,^9^ since the broad-spectrum empiric treatment approach that is pragmatic in hospitals with limited laboratory facilities^2,10^ generally results in overtreatment.^11^ Strengthening surveillance capacity in LMICs is therefore a major focus of international initiatives. To date, these programmes have generally focused on improving capacity for blood culture surveillance due to their high mortality; however, this requires substantial capital investment and sustainable funding for LMICs, where baseline laboratory functioning is poor.^12^ Blood cultures are comparatively costly, require trained staff due to the invasive nature of blood sampling, and can have slow turnaround times, reducing their ability to inform individual clinical management. Additionally, the low positivity rate means that very high sample throughput is needed to confidently estimate resistance prevalence. Consequently, empirical treatment guidelines are largely uninformed by local resistance rates in LMICs.

One way to address this would be to assess resistance rates using data from cheaper, easier non-invasive samples. Reflecting this, the 2020 WHO GLASS report includes resistance prevalence in urine cultures for key Gram-negative pathogens. However, associations between resistance rates in blood and other specimens have been rarely studied longitudinally. A 2018 report for South Korea observed similar resistance rates in *E. coli* blood cultures and urine isolates in 2016.^13^ A 2018 study showed statistical non-equivalence of antimicrobial susceptibility in nontyphoidal *Salmonella enterica* from blood, urine and faecal samples from 2015 U.S. National Antimicrobial Resistance Monitoring System data.^14^ Longitudinal surveillance of *Escherichia coli* bloodstream and urinary infections in a large UK region (Oxfordshire) found similar trends in co-amoxiclav resistance over 19 years.^15^ Another Oxfordshire study also found similar trends over time in nosocomial methicillin-resistant *Staphylococcus aureus* isolates from blood and non-blood clinical samples between 1998 and 2006,^16^ but changes in trends were identified earlier from non-blood samples due to higher numbers. This raises the broader question as to whether, for multiple pathogen-drug combinations, the proportion of resistant bloodstream infections could be estimated by the proportion of resistant isolates from other body-sites, that are easier to sample from, and more often culture-positive, reducing the costs per drug phenotype obtained. Using the latter as a surveillance method for the former is attractive because setting up a laboratory and providing the infrastructure (e.g. reagents, personnel, electricity etc.) to conduct blood culture surveillance in LMICs approximately doubles the costs compared to urine culture surveillance.^17^ Their large numbers make them ideal for developing locally relevant empiric treatment guidelines.

We therefore investigated the annual prevalence of resistance in isolates from blood and other body-sites for multiple pathogen-drug combinations in Oxfordshire (regional analysis, high-income country (HIC)). We then extended the analysis, first to consider concurrent or previous isolates in the same patient, and second to consider other world regions through publicly available datasets (Antimicrobial Testing Leadership and Surveillance (ATLAS)^18^) and collaborating hospitals/programmes in LMICs with available data.

## Materials and methods

We obtained antibiograms for *E. coli, Klebsiella* spp., *Staphylococcus aureus, Streptococcus pneumoniae*, and *Enterococcus* spp. from the Infections in Oxfordshire Research Database (IORD),^19^ from 1 January 1998 to 31 December 2018, which includes all microbiology tests performed for the region of approximately 680,000 individuals, and has Research Ethics Committee and Confidentiality Advisory Group approval (19/SC/0403, 19/CAG/0144) as a de-identified electronic research database. We compared *E. coli* and *Klebsiella* bloodstream infections (BSI) versus urinary tract infections (UTI); and isolates from blood versus all other body-sites of infection for other pathogens (non-blood infections; excluding surveillance swabs), further sub-divided into sterile versus non-sterile for *S. aureus* (Supplementary Methods). We estimated yearly resistance prevalence for all antimicrobials with susceptibilities for >65% samples in that year (mostly >80%). Testing used manual disk diffusion before 2013, and thereafter automated testing with the BD Phoenix™ Automated Microbiology System, Beckton Dickinson. Resistance was as defined by the laboratory when the test was done, using the same breakpoints regardless of specimen type, following European Committee on Antimicrobial Susceptibility testing (EUCAST) recommendations for bloodstream infections in each year.^20^ We did not de-duplicate isolates by patient, reflecting data that would be easily available from routine laboratory systems.

For each pathogen-drug combination, we first estimated the proportion resistant in blood versus other samples in each calendar year (with 95% confidence intervals (CI)) using Lin’s concordance correlation coefficient (CCC) for comparisons.^21^ We used time-series cross-correlation functions to identify the time differences at which the correlation between resistance prevalence was strongest. As perfect agreement is unrealistic, we also considered agreement within what might be considered clinically acceptable error (±5% and ±10% difference in prevalence), and agreement within arbitrary categories which may nevertheless be clinically helpful, specifically: <5% resistance (most would readily prescribe an antibiotic), 5-10% (most would prescribe for mild infections), 10-20% (unclear) and >20% (many would not prescribe if other options were available). We used logistic random-effects meta-analysis to estimate the overall difference between resistance prevalence in the two sample types, treating every calendar year as an independent study, assessing heterogeneity across years using the I^2^ statistic.^22^ We used meta-regression to estimate the effect of calendar year on the proportion of resistant bloodstream isolates (with its standard error), assuming the proportion of resistant isolates from other infection sites was known (given their greater numbers). We also used meta-regression to directly estimate the association between log odds of resistance in bloodstream infections (outcome, with its standard error) and the log odds of resistance in other infection sites (explanatory variable, fixed). (Full details in the Supplementary Material.)

One possibility is that resistance rates differ between blood and other infection sites because of differences in the underlying populations being sampled. A secondary analysis therefore considered matched pairs of isolates from the same patient, selecting the closest culture from a different site up to 3 days before or 2 days after the blood culture,^15^ assessing concordance using McNemar’s test. We also considered whether susceptibility of a previous *E. coli* UTI could predict susceptibility of a subsequent *E. coli* BSI, by selecting the temporally closest urine culture taken between 3-90 days before the blood culture.^15,23^

We also analysed pathogen-drug combinations at the country level (pooling multiple hospitals/regions) using the ATLAS dataset, comprising 633,820 isolates from 73 countries between 2004-2017.^18^ We considered all years where at least 30 isolates were tested for a given drug in LMICs (95% CI width around prevalence always <37%); and 100 isolates for HIC (width <20%) given greater numbers from HIC and strong inverse association between numbers tested and resistance prevalence in HIC, suggesting preferential testing of resistant isolates. Finally, we also analysed microbiology data from Angkor Hospital for Children (Siem Reap, Cambodia), Mahosot Hospital (Vientiane, Laos), and the Shoklo Malaria Research Unit (Mae Sot, Thailand, serving migrant and refugee populations on the Thailand-Myanmar border), including antibiotics tested for >40% of the isolates across all study years. Antimicrobial susceptibility testing was conducted using disk diffusion and results interpreted using Clinical and Laboratory Standards Institute (CLSI) 2019 breakpoints. Intermediate isolates were considered non-susceptible. Extended Spectrum Beta-Lactamase (ESBL) status was confirmed using the double disk method (ceftazidime+/-clavulanate and cefotaxime+/-clavulanate) as a ≥5mm increase in a zone diameter for either agent with clavulanic acid.

We conducted all analyses using R 3.5.1, using the metafor package for meta-analyses.^24^

## Results

We focused initially on *E. coli* and *S. aureus* as two of the most common bacteria causing BSI,^2,25,26^ and often causing urine or skin and soft tissue infections, respectively. We included 8102 *E. coli* BSI and 322087 *E. coli* UTI (40-fold higher) from Oxfordshire (**Supplementary Figure 1**); 4 antibiotics (amoxicillin, co-amoxiclav, ciprofloxacin, and trimethoprim) were consistently tested in both between 1998-2018. Whilst absolute resistance prevalence was generally slightly higher in bloodstream isolates, trends in the proportion of resistant BSI and UTI were very similar, notably tracking substantial fluctuations over time in trimethoprim resistance (**Figure 1, Supplementary Table 1, Supplementary Figure 2**), with resistance rates in the same year most strongly correlated (maximum cross-correlation at lag 0). For these four antibiotics, there was no evidence of strong variation in the relationship between resistance rates over time (**Supplementary Table 1**).

**Figure 1.**
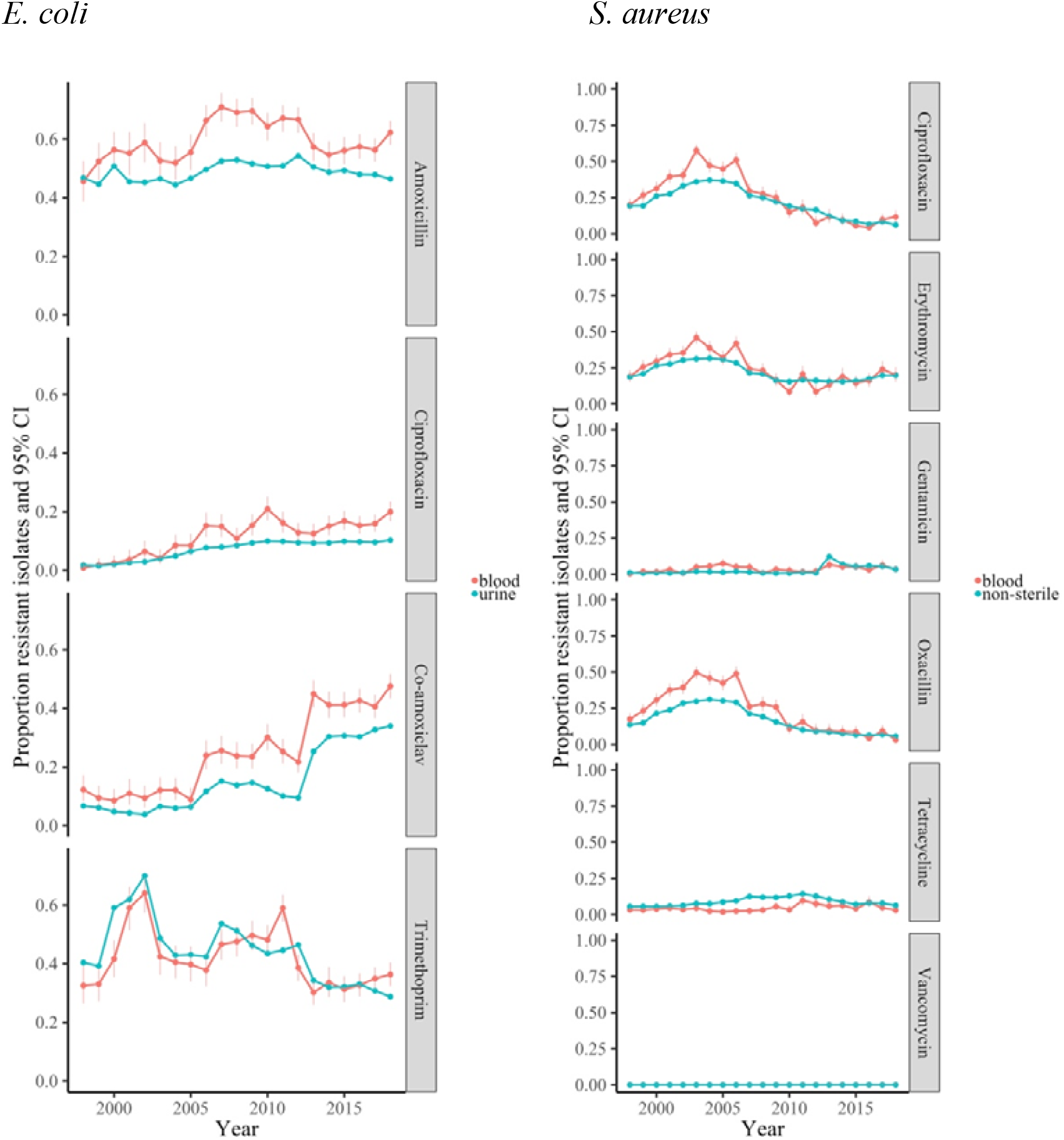
Resistance prevalence in E. coli and S. aureus in blood versus non-blood cultures in Oxfordshire, 1998-2018 Note: maximum cross-correlation at time lag 0 in 3/4 drugs for *E. coli* and 4/6 drugs for *S. aureus*; cross-correlation 0.77 at lag 1 (0.75 at lag 0), 0.95, 0.96, 0.80 for *E. coli* and 0.95, 0.93, 0.54, 0.75, 0.69 at lag −4, 0.60 at lag 3 for *S. aureus* from top to bottom respectively.

Including a further six antibiotics (aztreonam, ceftazidime, ceftriaxone, co-trimoxazole, ertapenem, gentamicin, meropenem and piperacillin-tazobactam) consistently tested between 2013-2018 (**Supplementary Figure 3, Supplementary Table 1**) increased the range of resistance prevalences observed. Again, resistance rates were slightly higher in BSI and showed reasonable concordance with resistance in UTI, with prevalences within 10% of each other in 109/132 (89%) drug-years (**Figure 2**, left-hand panel, **Table 1**). Overall resistance prevalence in BSI was highly correlated with resistance prevalence in UTI (CCC=0.93 (95% CI 0.91-0.95) (**Table 1)**. Agreement was also relatively high across our four pre-defined resistance categories, with 83/132 (63%) drug-years in the same resistance category and 45/132 (34%) in adjacent categories (**Figure 2**, right-hand panel). Although numbers were smaller, broadly similar results were seen for *Klebsiella* spp. (**Figure 2, Table 1, Supplementary Figures 4&5, Supplementary Tables 1&2**).

**Figure 2.**
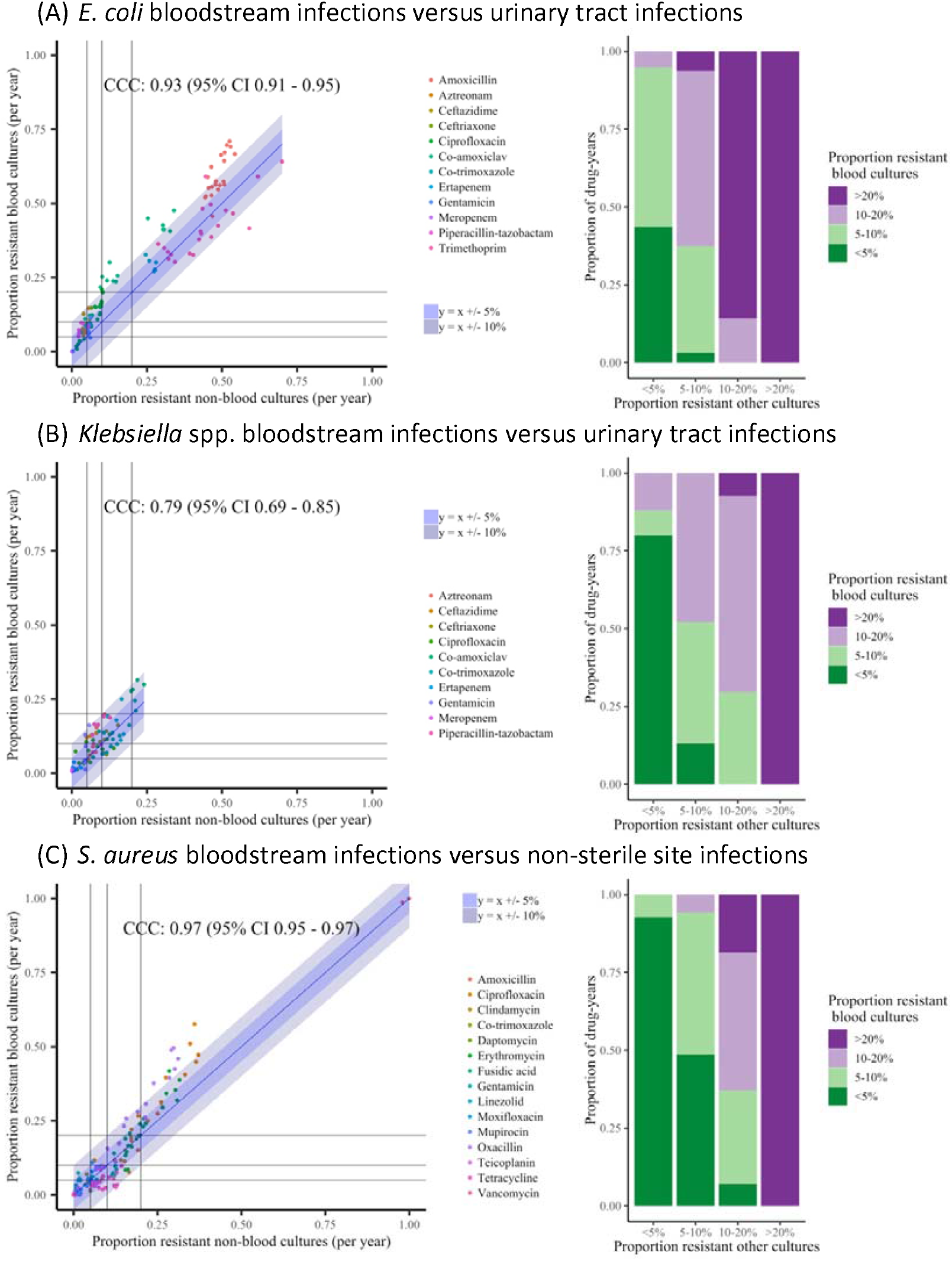
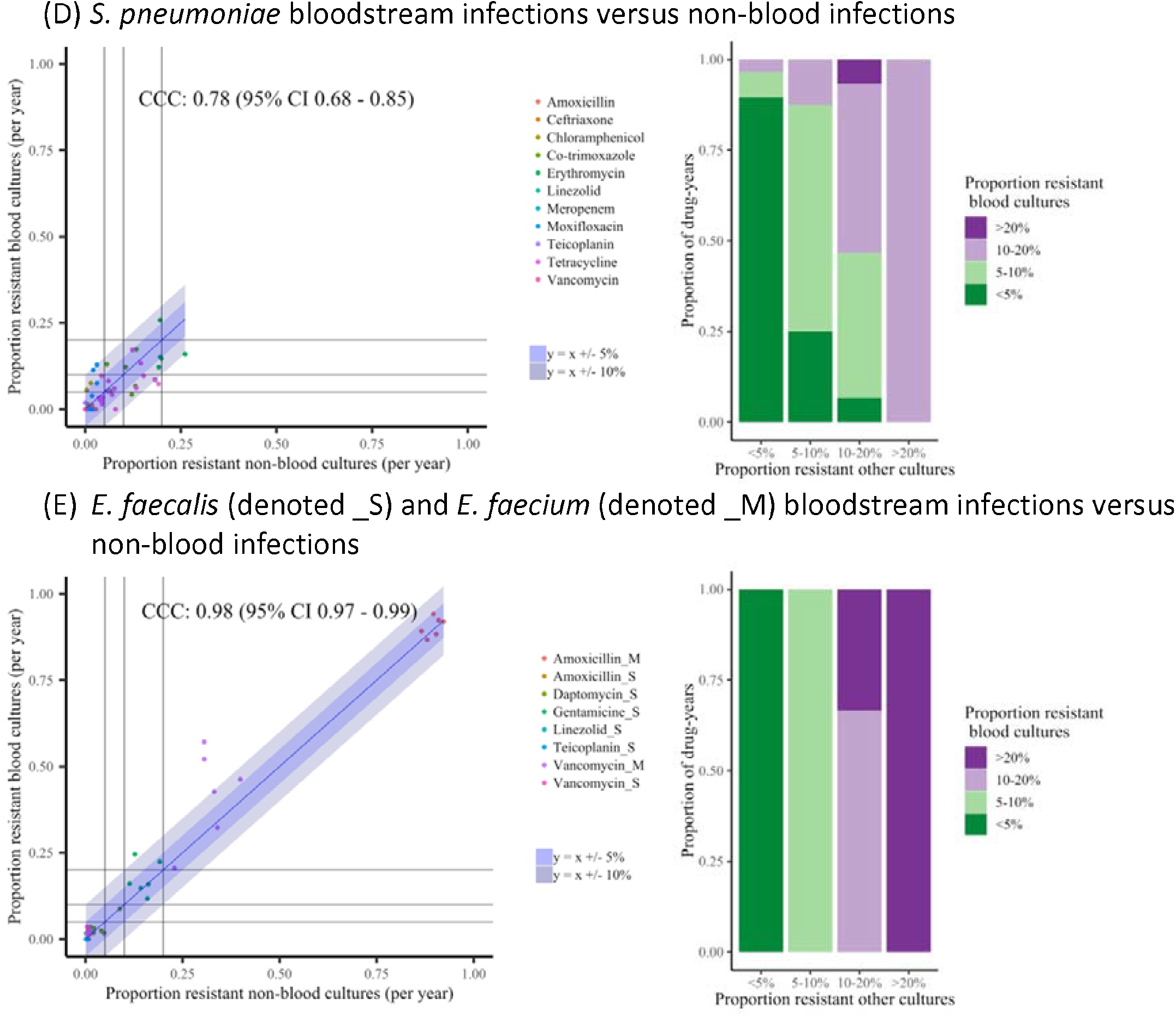
Resistance in blood and non-blood cultures for (A) E. coli, (B) Klebsiella spp., (C) S. aureus, (D) S. pneumoniae, (E) E. faecalis and E. faecium, Oxfordshire 1998-2018 Note: Left: Proportion resistant non-blood cultures versus proportion resistant blood cultures per year for all antibiotics. Each dot reflects resistance prevalence for one antibiotic in one specific year. Line of identity, +/- 5% and 10% shown in blue. Lines are drawn at 0.05 (5%), 0.1 (10%) and 0.2 (20%) resistance prevalence in each of blood versus non-blood cultures for ease of visualisation of agreement between clinically meaningful resistance categories in these two types of samples. Right: Grouping points from left column into clinically meaningful resistance categories; classifying resistance prevalence in blood cultures by the proportion in urine/non-sterile site cultures.

**Table 1.** Resistance prevalence in blood and non-blood cultures across drug-years, Oxfordshire 1998-2018

| Pathogen | Lin's concordance coefficient (95% CI) | <5% difference in prevalence | <10% difference in prevalence | Same resistance category* | One resistance category* different | Two resistance categories* different |
| --- | --- | --- | --- | --- | --- | --- |
| **Oxfordshire** |  |  |  |  |  |  |
| *E. coli* | 0.93 (0.91-0.95) | 67/132 (51%) | 109/132 (83%) | 83/132 (63%) | 45/132 (34%) | 4/132 (3%) |
| *Klebsiella* spp. | 0.79 (0.69-0.85) | 54/80 (68%) | 79/80 (99%) | 51/80 (64%) | 26/80 (32%) | 3/80 (4%) |
| *S. aureus* | 0.97 (0.95-0.97) | 140/180 (78%) | 167/180 (93%) | 132/180 (73%) | 45/180 (25%) | 3/180 (2%) |
| *S. pneumoniae* | 0.78 (0.68-0.85) | 63/81 (78%) | 79/81 (98%) | 63/81 (78%) | 15/81 (19%) | 3/81 (4%) |
| *E. faecalis* | 0.90 (0.82-0.94) | 35/36 (97%) | 35/36 (97%) | 34/36 (94%) | 2/36 (6%) | 0/36 (0%) |
| *E. faecium* | 0.92 (0.77-0.97) | 8/12 (67%) | 9/12 (75%) | 12/12 (100%) | 0/12 (0%) | 0/12 (0%) |
| **ATLAS high-income countries** |  |  |  |  |  |  |
| *E. coli* | 0.97 (0.96-0.97) | 302/401 (75%) | 376/401 (94%) | 326/401 (81%) | 74/401 (18%) | 1/401 (0%) |
| *S. aureus* | 0.99 (0.99-0.99) | 140/155 (90%) | 147/155 (95%) | 154/155 (99%) | 1/155 (1%) | 0/155 (0%) |
| *K. pneumoniae* | 0.89 (0.86-0.92) | 135/186 (73%) | 172/186 (92%) | 134/186 (72%) | 51/186 (27%) | 1/186 (1%) |
| *P. aeruginosa* | 0.89 (0.80-0.94) | 27/35 (77%) | 34/35 (97%) | 25/35 (71%) | 10/35 (29%) | 0/35 (0%) |
| **ATLAS middle-income countries** |  |  |  |  |  |  |
| *E. coli* | 0.95 (0.94-0.96) | 157/268 (59%) | 213/268 (79%) | 234/268 (87%) | 31/268 (12%) | 3/268 (1%) |
| *S. aureus* | 0.96 (0.95-0.97) | 260/344 (76%) | 308/344 (90%) | 289/344 (84%) | 47/344 (14%) | 8/344 (2%) |
| *K. pneumoniae* | 0.86 (0.82-0.89) | 87/228 (38%) | 146/228 (64%) | 170/228 (75%) | 53/228 (23%) | 5/228 (2%) |
| *P. aeruginosa* | 0.77 (0.60-0.88) | 20/36 (56%) | 31/36 (86%) | 32/36 (89%) | 4/36 (11%) | 0/36 (0%) |
| *E. cloacae* | 0.96 (0.93-0.97) | 31/61 (51%) | 45/61 (74%) | 49/61 (80%) | 10/61 (16%) | 2/61 (3%) |
| **Cambodia** |  |  |  |  |  |  |
| *E. coli* | 0.67 (0.53-0.78) | 16/70 (23%) | 27/70 (39%) | 63/70 (90%) | 4/70 (6%) | 3/70 (4%) |
| *S. aureus* | 0.97 (0.95-0.98) | 22/48 (46%) | 39/48 (81%) | 30/48 (62%) | 15/48 (31%) | 3/48 (6%) |
| **Laos** |  |  |  |  |  |  |
| *E. coli* | 0.83 (0.64-0.92) | 3/20 (15%) | 10/20 (50%) | 20/20 (100%) | 0/20 (0%) | 0/20 (0%) |
| *S. aureus* | 0.96 (0.85-0.99) | 4/10 (40%) | 8/10 (80%) | 7/10 (70%) | 3/10 (30%) | 0/10 (0%) |

| Pathogen | Lin's concordance coefficient<br>(95% CI) | <5% difference in prevalence | <10% difference in prevalence | Same resistance category* | One resistance category* different | Two resistance categories* different |
| --- | --- | --- | --- | --- | --- | --- |
|  | **Thailand** |  |  |  |  |  |
| *E. coli* | 0.91 (0.87-0.94) | 48/120 (40%) | 77/120 (64%) | 77/120 (64%) | 34/120 (28%) | 9/120 (8%) |
| *S. aureus* | 0.97 (0.96-0.98) | 56/84 (67%) | 72/84 (86%) | 55/84 (65%) | 21/84 (25%) | 8/84 (10%) |

\* Resistance categories are: <5%, 5-10%, 10-20%, >20%
Note: in the Oxfordshire dataset, non-blood cultures are urine for *E. coli* and *Klebsiella* spp., other non-sterile sites for *S. aureus* and all other body sites for other gram- negatives and *S. pneumoniae*, *E. faecalis* and *E. faecium*; in the ATLAS dataset non-blood cultures are all other body sites for all pathogens; in the three hospitals from LMICs non-blood cultures are urine for *E. coli* and swabs/pus for *S. aureus*

Considering matched pairs of isolates from individual patients, there was no evidence that the modestly different resistance rates seen in *E. coli* BSIs and UTIs overall were due to differential sampling of intrinsically different underlying populations, given high but similarly imperfect agreement in susceptibility (86-100% across 11 antibiotics for concurrent infections, 80-94% for previous infections, **Supplementary Table 5**). In particular, considering the closest prior *E. coli* UTI 3-90 days before a BSI, >80% resistant UTIs also had a resistant BSI across these four drugs.

Between 1998-2018, there were 6952 *S. aureus* BSIs and 166179 *S. aureus* non-blood cultures: 54105 (33%) from sterile sites and 112074 (67%) from non-sterile sites (8- and 16-fold higher respectively, **Supplementary Figure 1)**. Ciprofloxacin, erythromycin and oxacillin presented similar trends in the proportions resistant over time in the different sample types, which varied substantially over time (**Figure 1, Supplementary Table 1, Supplementary Figure 6**), again supporting the plausibility of using non-blood cultures as a proxy for blood culture surveillance. Agreement was poorer for gentamicin and tetracycline resistance, although rates were low (<15%). As might be expected, agreement was slightly better between resistance prevalence in blood and other sterile site cultures than blood and non-sterile site cultures (**Supplementary Figure 7**). As agreement was reasonable in both, and non-sterile site cultures would be more likely to be taken in LMICs, we focussed on this group of infection sites. Overall, resistance prevalence was more similar in bloodstream and other non-sterile infection sites in *S. aureus* than *E. coli* (**Supplementary Table 1**), but there were also slightly stronger effects of calendar time for *S. aureus*, reflecting generally higher rates of resistance in blood compared with non-sterile site cultures during the MRSA epidemic in the mid-2000s, which then became more similar (**Supplementary Figure 3**).

For *S. aureus*, including other antibiotics tested between 2013-2018 only, resistance prevalences in blood and non-sterile site cultures were within 10% of each other in 167/180 (93%) drug-years, with 132 (73%) drug-years in the same resistance category and 45 (25%) in adjacent categories (**Figure 2, Table 1**). Overall resistance prevalence in blood was highly correlated with resistance prevalence in non-sterile isolates (CCC=0.97 (0.95-0.97)) (**Table 1)**. Overall agreement between resistance in concurrent blood and non-sterile cultures within a patient was extremely high (96-100%) for *S. aureus*, with ciprofloxacin, erythromycin and oxacillin having <5% of resistant blood cultures with susceptible non-sterile cultures and <4% susceptible blood cultures with resistant non-sterile sites cultures (**Supplementary Table 5**). Again, although numbers were smaller, broadly similar results were seen for *S. pneumoniae, E. faecalis* and *E. faecium* (**Figure 2, Table 1, Supplementary Figure 8, Supplementary Tables 1&2**).

In the ATLAS dataset, there were 401 and 155 country-drug-years with >100 *E. coli* and *S. aureus* isolates from HICs, respectively, from both bloodstream and urinary/non-sterile infections (**Supplementary Table 3**). For *E. coli*, time trends could be estimated over 7 years or more in four countries for 11 antibiotics, and were broadly similar (**Figure 3**; *S. aureus* in **Supplementary Figure 9**). Across all drugs, prevalence of resistance in *E. coli* BSIs and UTIs were within 10% of each other in 376/401 (94%) country-drug-years (**Figure 4**, left-hand panel, **Table 1**); 326 (81%) country-drug-years were in the same resistance category and 74 (18%) in adjacent categories (**Figure 4**, right column). For *S. aureus*, respective figures were 147/155 (95%), 154 (99%) and 1 (1%). Agreement was similar in middle-income countries where smaller numbers of isolates were available (**Figure 4, Table 1, Supplementary Figures 10&11**) and in other pathogens (**Supplementary Figures 12-16**).

**Figure 3.**
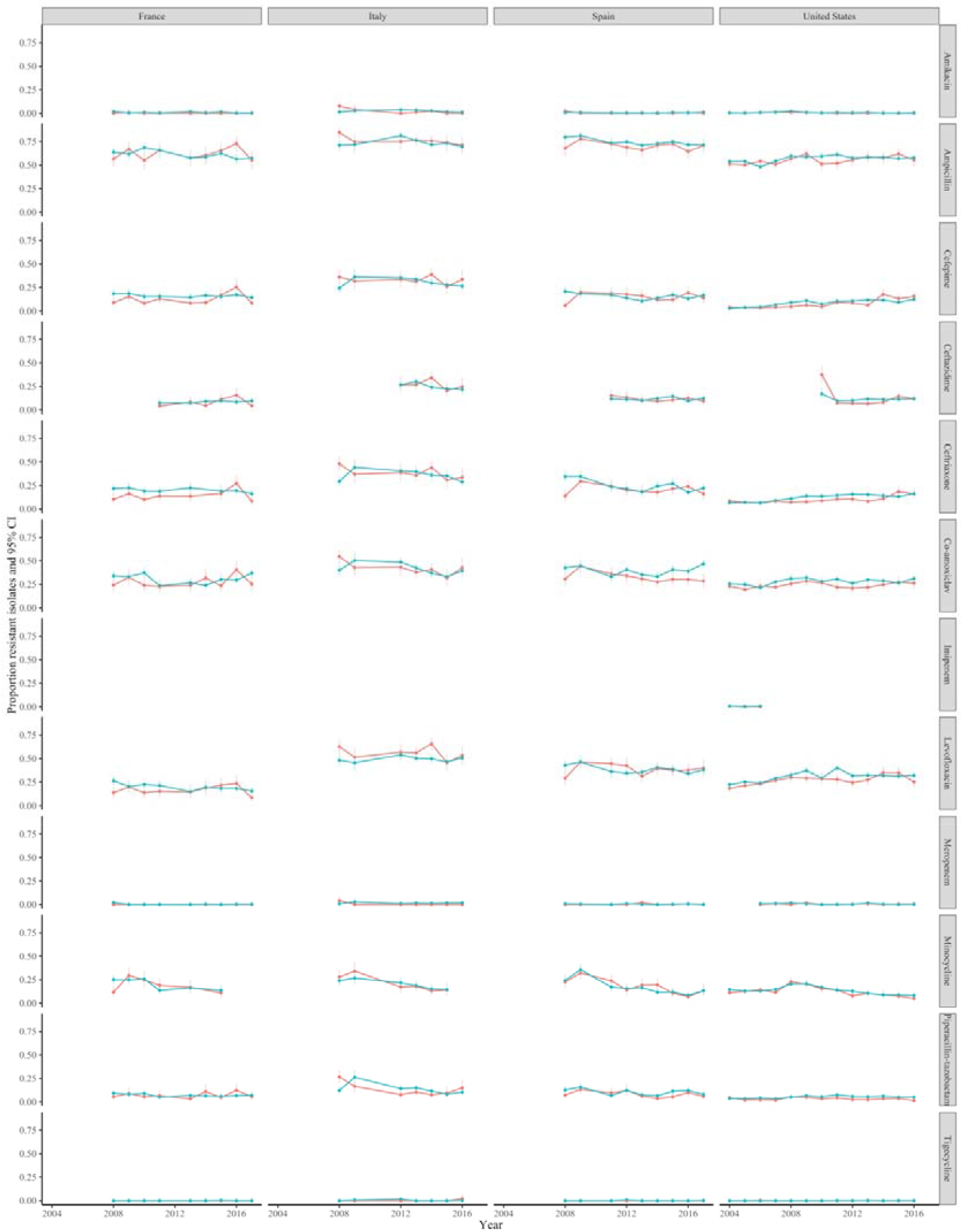
Resistance prevalence in E. coli in blood versus non-blood cultures in high-income countries present in the ATLAS dataset, 2004-2016

Considering hospital/programme-level LMIC datasets, numbers were smaller (**Supplementary Table 4**) and hence estimates more variable, but both time trends (**Figure 5, Supplementary Figure 17**) and comparisons of individual country-drug-years (**Figure 6, Table 1, Supplementary Figure 18**) supported overall findings that resistance profiles in non-blood culture isolates correlate well with those in blood culture isolates over time.

**Figure 4.**
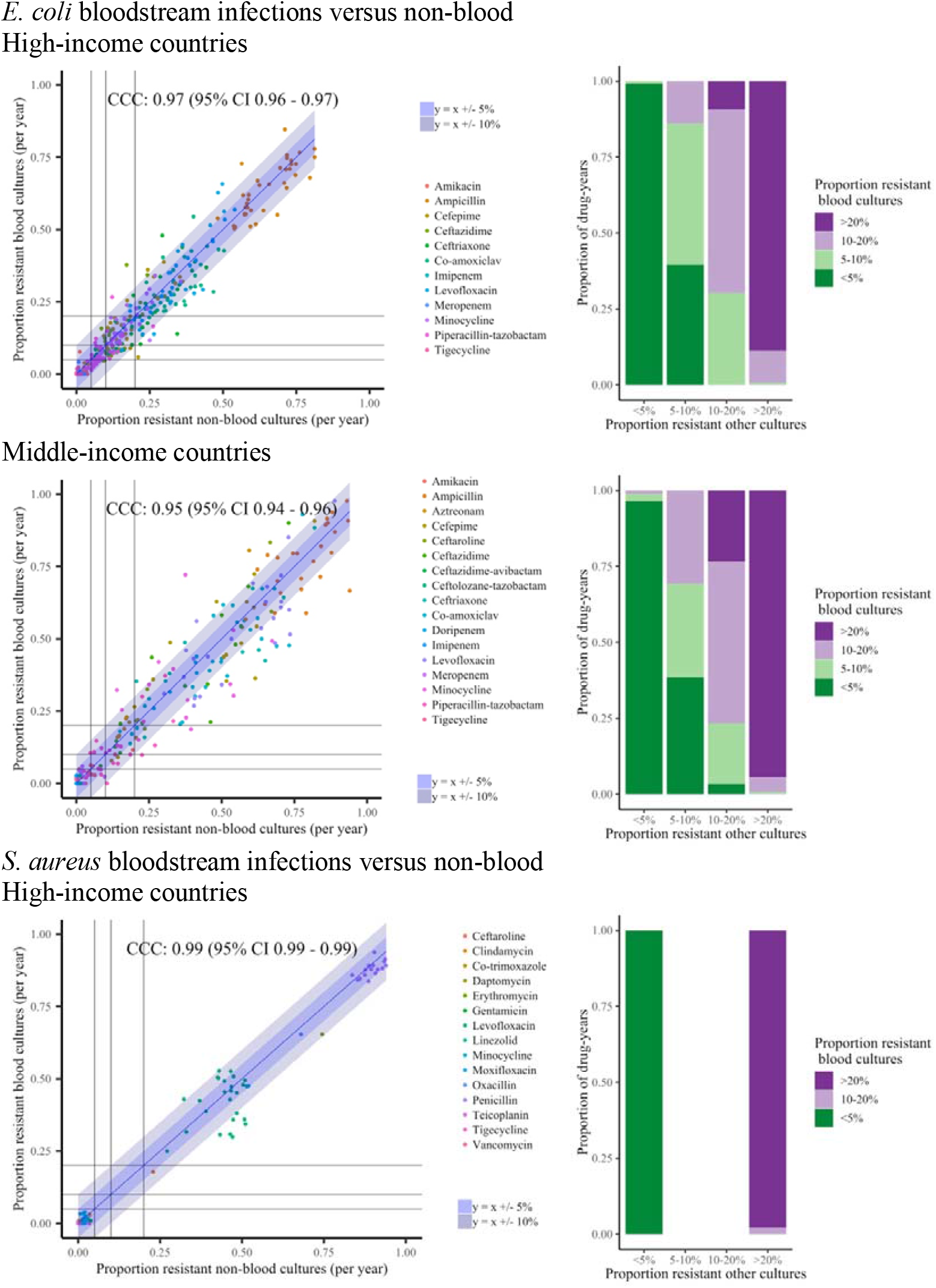
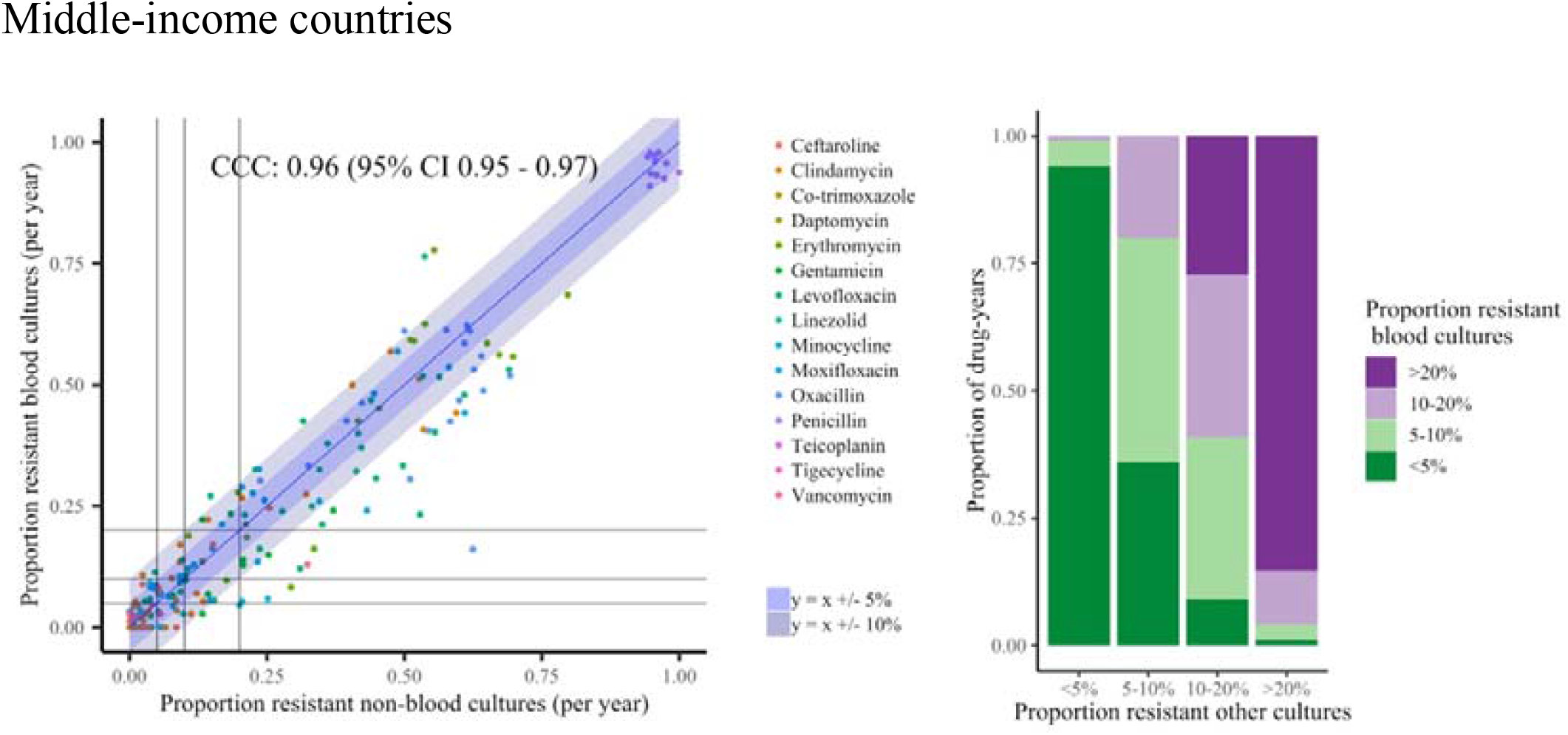
Resistance in blood and non-blood cultures (A) E. coli, (B) S. aureus, ATLAS dataset split into high-income countries and middle-income countries Note: Left: Proportion resistant non-blood cultures versus proportion resistant blood cultures per year for all antibiotics. Each dot reflects resistance prevalence for one antibiotic in one specific year. Line of identity, +/- 5% and 10% shown in blue. Lines are drawn at 0.05 (5%), 0.1 (10%) and 0.2 (20%) resistance prevalence in each of blood versus non-blood cultures for ease of visualisation of agreement between clinically meaningful resistance categories in these two types of samples. Right: Grouping points from left column into clinically meaningful resistance categories; classifying resistance prevalence in blood cultures by the proportion in urine/non-sterile site cultures.

**Figure 5.**
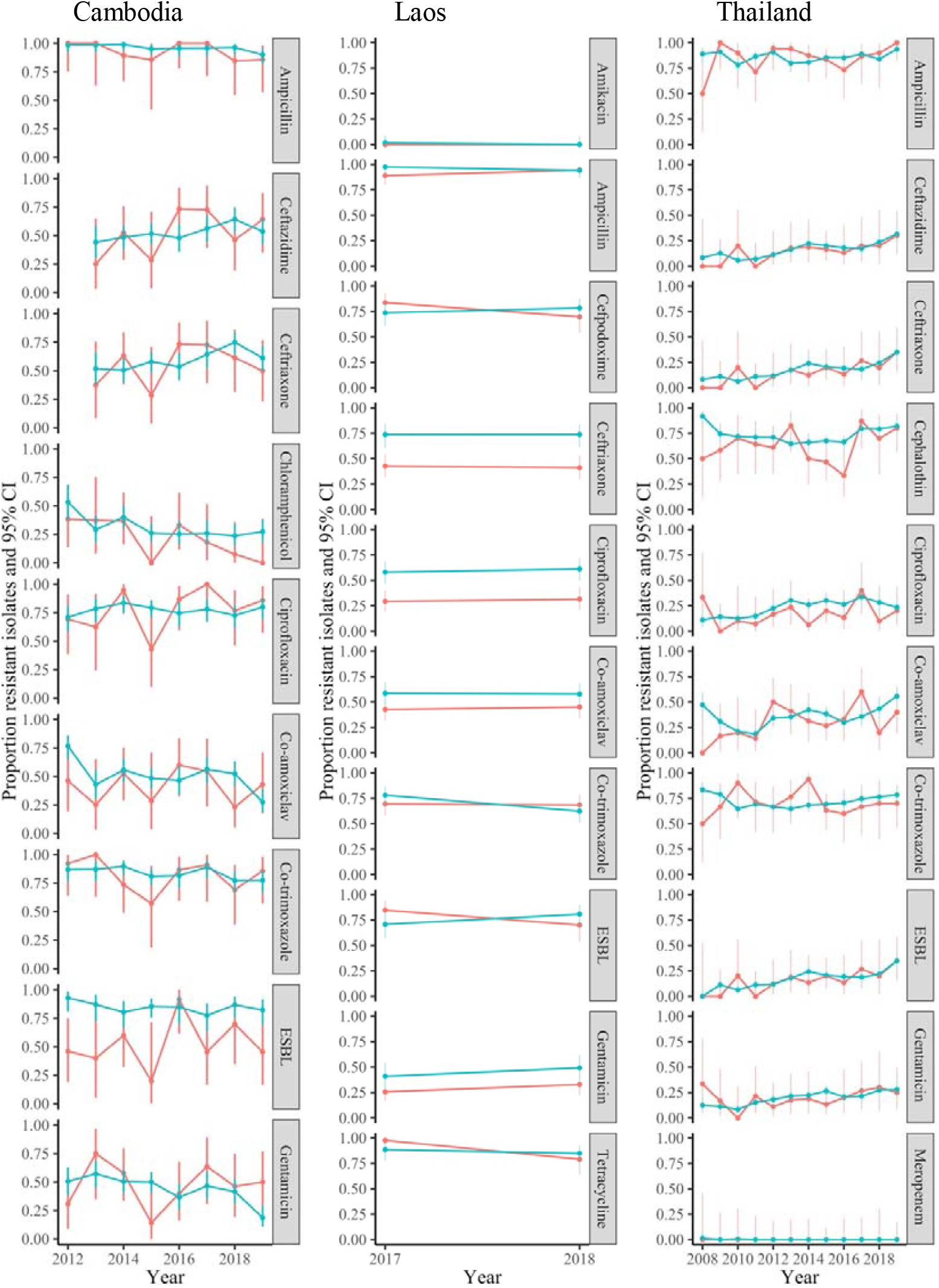
Resistance prevalence in E. coli in blood versus non-blood cultures in three hospitals/programmes in LMICs

**Figure 6.**
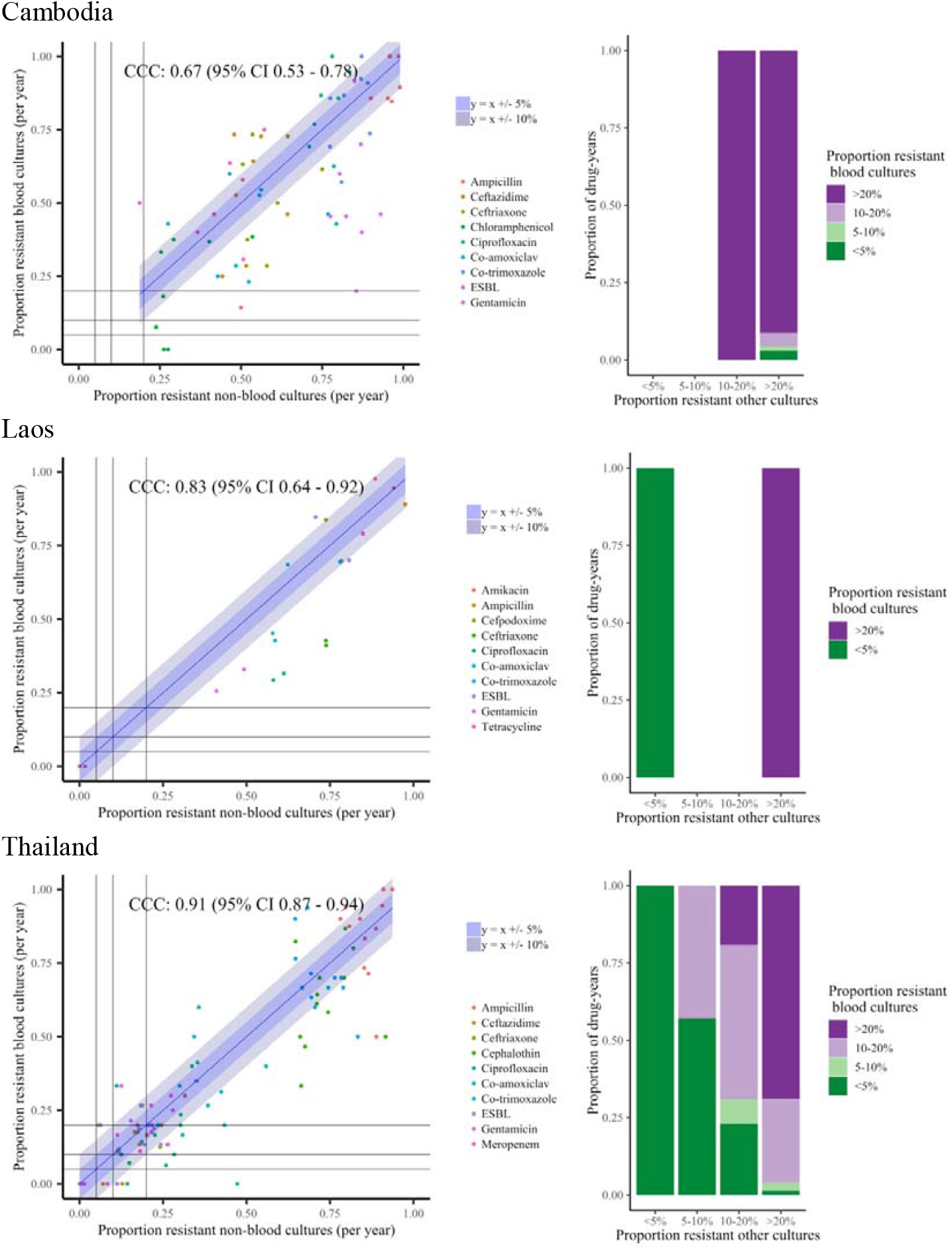
Resistance in E. coli blood and urine cultures in three hospitals/programmes in LMICs Note: Left: Proportion resistant non-blood cultures versus proportion resistant blood cultures per year for all antibiotics. Each dot reflects resistance prevalence for one antibiotic in one specific year, with 21 dots per antibiotic (one for each year 1998-2018). Line of identity, +/- 5% and 10% shown in blue. Lines are drawn at 0.05 (5%), 0.1 (10%) and 0.2 (20%) resistance prevalence in each of blood versus non-blood cultures for ease of visualisation of agreement between clinically meaningful resistance categories in these two types of samples. Right: Grouping points from left column into clinically meaningful resistance categories; classifying resistance prevalence in blood cultures by the proportion in urine/non-sterile site cultures.

## Discussion

Here, we show that AMR prevalence from infections in sites other than blood could potentially be used to infer AMR in bloodstream infections, which could reduce the initial costs of AMR surveillance in LMICs. Focusing on two of the most common bacteria causing serious infections, *E. coli* and *S. aureus*, we found that, whilst not perfect, agreement between the resistant rates in blood cultures and in other clinical cultures was good for most antibiotics, in terms of actual values, but, most importantly for surveillance, over time. This suggests that changes in resistance over time could be monitored using samples that are easier to collect than blood, and could also inform appropriateness of different empirical antimicrobial therapies.

Whilst many studies estimate resistance prevalence in various infection sites, to our knowledge, very few studies have directly compared resistance in blood versus other infection sites, either focussing on one specific drug-pathogen combination,^16^ or on Enterobacterales and investigating metagenomics from pooled human samples from the UK, Kenya and Cambodia as a predictor of resistance in invasive infections.^27^ In 2011, Health Protection Scotland and National Services Scotland published a protocol for the surveillance of AMR in UTIs, arguing that it would be more appropriate to monitor resistance in other sites where an infection initially starts before entering the bloodstream.^28^ However, the hypothesised relationship was not analysed using the data collected.^29^ WHO piloted a similar approach but did not compare with resistance rates in BSI.^30^

Current efforts on AMR surveillance in LMICs are directed towards building laboratories and training staff to carry out antimicrobial susceptibility testing on blood cultures, arguing that if this can be achieved, other types of culture will become possible too. However, setting up such systems is very expensive, both in terms of infrastructural and running costs, and it will be many years until sufficient data on blood cultures have been accumulated for surveillance purposes, particularly given initial low uptake and low rates of positivity in these samples. While longitudinal surveillance data is available at a limited number of research sites, these programmes cover a limited geographic area and may not be representative of other locations in a country. Incorporating blood culture testing into public hospitals may be further biased by differences in ability to pay.^2^ Collecting specimens from urine/skin surface/genital sites is cheaper and less invasive than sampling blood, and therefore substantially easier to do on large numbers from different communities, as well as having higher positivity rates, increasing their utility for AMR surveillance. This is true even in HICs, with clearly narrower confidence intervals from larger numbers of non-blood Oxfordshire isolates. Even allowing for the greater manual processing needed for non-blood cultures, for example, obtaining 100 *E. coli* isolates would only require testing 361 urine samples assuming 37% are culture-positive and 75% of these are *E. coli*, but 7693 blood cultures, assuming 13% are positive and 10% of these are *E. coli* (estimates based on Oxfordshire data 1998-2016). Whilst blood cultures may be perceived as having greater diagnostic utility, this may be limited practically, e.g. by long turnaround times.

One strength of our study is the different datasets used, and the robustness of findings across these, including continuous surveillance of one large region (Oxfordshire), and global analysis (ATLAS) albeit of a selection of infections within each country. The hospital/programme-level LMIC datasets are highly curated, but relatively small. One crucial underpinning assumption is that pathogens causing other infections are representative of pathogens causing BSI in terms of antibiogram, either because the source of infection is commonly from commensal colonising organisms that become pathogenic opportunistically or because both types of infection arise from a similar reservoir. Even though this may hold within the populations studied, it is possible, although fairly unlikely, that this may not generalise more widely. Generalisability is supported by the good country-level agreement in ATLAS, even though pooling different regions within countries, which could have potentially differed further between blood and other infection sites. This raises an important limitation of our approach, namely that different people were sampled for blood and other specimens. However, our analysis of samples from the same patients suggests this is unlikely to cause major bias. Another limitation is that we have only considered surveillance of the proportion of resistant infections, not numbers or population-level rates of resistance; however, in terms of empiric treatment recommendations, proportions rather than absolute numbers that are generally of interest. We have not considered surveillance of AMR across the entire antibiogram, i.e. patterns of resistance within one isolate across different antibiotics. We also pooled all samples together: in future work we could consider whether relationships are generalizable between different groups of patients (e.g. by age, nosocomial/community infections). Finally, the thresholds we considered were arbitrary, which is why we considered two different approaches (varying margins of error and varying categorical thresholds).

One question is how good the agreement between resistance rates in isolates from blood versus other specimens should be for surveillance of resistance in samples from other body-sites to be useful – clinically and for understanding overall trends. In our dataset, different resistance rates were observed in different sample types for many pathogen-drug combinations, with some variation over time and less than perfect agreement. However, cross-correlations were generally high, as was agreement within 10% and across categories that could inform empirical antimicrobial therapy. Several biological reasons for the lack of perfect agreement are plausible: for example, a large proportion of *E. coli* BSIs have a urinary focus, but susceptibility and appropriate empiric treatment generally limit progression to BSI, in contrast to resistant, and perhaps sub-optimally treated UTIs. Hence one might expect the proportion of drug-resistant BSIs to be higher due to inadequate empiric treatment of a drug-resistant UTI, but underlying time trends to track each other, as we observed. One could ask whether blood is the best sample for AMR surveillance. BSIs are the most serious infection, so risks associated with inappropriate antibiotic treatment are highest, but potentially treating the many other less serious infections correctly might mean fewer develop into BSI.

In HICs, susceptibility testing is still not instantaneous, with microbiology results generally taking at least 48 hours, time during which empiric antimicrobials are administered, potentially leading to poorer outcomes if the infection is caused by a resistant organism.^3^ However, increasing use of electronic health records means it is becoming much simpler to check for previous UTIs and use this to guide empiric treatment. Our paired analysis illustrates the potential of this approach based on an arbitrarily chosen interval of 3-90 days prior to the BSI; future work could investigate how the strength of this association varies further back in time, or using the most resistant rather than most recent urinary isolate. Similarly, Yelin *et al*.^31^ found that incorporating demographic information with susceptibility results of previous UTIs and antibiotic purchases in a machine learning model improved guidance of empirical treatment for new community-acquired UTIs.

AMR surveillance plays a key role in optimising the use of antibiotics and hence reducing resistance: asking clinicians to reduce antibiotic use and running successful stewardship programmes cannot be done in an information vacuum, and lack of microbiology facilities and unwillingness of patients to have samples taken (often due to cost) have been identified as key factors influencing antibiotic use.^32,33^ LMICs face numerous challenges in setting up surveillance systems similar to those in HICs, including lack of coherent governance, budget, technical expertise, information technology systems and co-ordination. Our study shows that body-sites which are easier to sample, cheaper, faster and easier to grow from compared to blood could provide a feasible approach to AMR surveillance, providing evidence for empiric treatment recommendations. They could also be amenable to survey-type approaches which could be particularly valuable as setting up a local laboratory infrastructure for successful and useful antibiogram production is challenging. Using population level surveillance as a substitute for individual patient testing could be considered a pragmatic steppingstone in improving a site’s diagnostic capacity for individual patient management. Nevertheless, the urgent threat posed by AMR demands that we not let the perfect be the enemy of the good.

## Data Availability

Oxfordshire data comes from the Infections in Oxfordshire Research Database https://oxfordbrc.nihr.ac.uk/research-themes-overview/antimicrobial-resistance-and-modernising-microbiology/infections-in-oxfordshire-research-database-iord/,
the ATLAS dataset is publicly available here:
https://www.synapse.org/#!Synapse:syn17009517/wiki/585653
The data from Cambodia, Laos and Thailand is not publicly available.

## Acknowledgements

We thank all the people of Oxfordshire who contribute to the Infections in Oxfordshire Research Database. Research Database Team: R Alstead, C Bunch, DCW Crook, J Davies, J Finney, J Gearing (community), L O’Connor, TEA Peto (PI), TP Quan, J Robinson (community), B Shine, AS Walker, D Waller, D Wyllie. Patient and Public Panel: G Blower, C Mancey, P McLoughlin, B Nichols.

We thank Liam Shaw for discussion about the ATLAS dataset and feedback on the manuscript.

We thank the diagnostic microbiology laboratory staff at Angkor Hospital for Children, Mahosot Hospital Microbiology Laboratory, and the Shoklo Malaria Research Unit clinic and laboratory staff.

## Transparency declaration

Karina-Doris Vihta affirms that this manuscript is an honest, accurate, and transparent account of the study being reported; that no important aspects of the study have been omitted; and that any discrepancies from the study as planned (and, if relevant, registered) have been explained.

## Contributors

K-DV, NJW, TEAP, and ASW designed the study. TPQ prepared extracts from the Infections in Oxfordshire Research Database. K-DV obtained data from the Antimicrobial Testing Leadership and Surveillance (ATLAS). Data from the Angkor Hospital for Children was provided by PT, from the Mahosot Hospital Microbiology Laboratory by MV, EAA, VC, and from the Shoklo Malaria Research Unit by CL. K-DV, TEAP and ASW analysed the data. K-DV, TEAP, and ASW prepared the figures. K-DV, DE, TEAP, and ASW prepared the first draft of the manuscript. All authors commented on the data and its interpretation, revised the content critically, and approved the final version. *NIHR Health Protection Research Unit Steering Committee:* J Coia, N French, C Marwick, M Sharland.

