## Supplementary Material for "Antimicrobial resistance surveillance: can we estimate resistance in bloodstream infections from other types of specimen?"

**Supplementary Methods**

***Additional details of pathogens considered***

For *S. aureus* isolates, as previously,^1^ we excluded cultures from vascular tips due to their relationship with blood cultures, and methicillin-resistant *S. aureus* (MRSA) screening cultures because only MRSA could be isolated from these. We then categorised the remaining *S. aureus* cultures into bloodstream infections and *S. aureus* non-blood cultures. Non-blood cultures were further subdivided into:

- sterile sites: clinical samples cultured from respiratory and normally sterile sites taken under aseptic technique, such as ascites, joint aspirates, cerebrospinal fluid, pre-prosthetic material, and collections of pus from deep sites, which one could hypothesise would be more similar to blood cultures in terms of resistance since they also reflect invasive infection. They are still potentially harder to obtain (e.g. needing sampling during operations, etc), but once obtained easier to culture.
- non-sterile sites: all other clinical samples, typically surface swabs, skin surface, usually superficial abscess swabs, genital, urine, which are easier to obtain and therefore more likely to be available, but may reflect colonisation with a commensal rather than infection with a pathogen.

The majority of *S. aureus* cultures were tested for susceptibility to vancomycin; however, there were no resistant bloodstream infections, only 2 resistant sterile site cultures and 4 resistant non-sterile site cultures. We therefore did not consider vancomycin resistance for individual drug analyses.

For *Enterococcus* spp., non-blood cultures were not routinely speciated before 2013 and therefore the analyses were only carried out for 2013-2018.

For *Klebsiella* spp., urine cultures were not routinely speciated between 2007-2012 and therefore these years were excluded from comparisons as numbers were very small and resistance prevalence unrealistically high, suggesting non-random speciation of highly resistant samples.

In the Cambodia, Laos and Thailand hospitals/programmes extended spectrum beta-lactamase (ESBL) testing should only be carried out if ceftriaxone zones are less than or equal to 25mm and/or ceftazidime zones are less than or equal to 22mm. Not taking this into account leads to a small proportion of isolates being tested and a large percentage being called positive. We therefore recoded those that were not tested and that should have not been tested according to the rules above, negative, leaving the ones that were not tested and that should have been tested as missing.

***Additional details of statistical methods***

In comparisons of resistance prevalence between blood vs non-blood samples, we considered the number differing by more than 5% or 10%, and also whether the 95% CIs lay within these boundaries, to estimate usefulness as a surveillance tool. For surveillance, we also considered the numbers agreeing within fixed prevalence groups that might influence decisions on empiric antibiotic treatment, specifically under 5%, 5-10%, 10-20% and 20% or more.

Lin’s concordance correlation coefficient (CCC)^2^ quantifies the agreement between the proportion of resistant non-blood cultures and the gold standard, here the proportion of resistant blood cultures, analogous to the Spearman’s correlation coefficient, but also taking into account the fact that the expectation is that the two variables being compared should be the same. It combines measures of precision and accuracy in order to determine how far the observed values deviate from the line of perfect concordance, overcoming some of the limitations of correlation coefficients in general, paired t-tests and least squares approaches.^2^ However, it does not allow for the fact that resistance prevalences are estimated with error, in bloodstream infections and, to a much lesser degree (due to larger numbers) in other infection sites. In order to estimate associations taking these standard error estimates into account, we considered three linear mixed models (or special cases of these).

First, we conducted a standard meta-analysis: in traditional meta-analysis terms, we treated every calendar year as an independent study, resistant vs susceptible as the outcome, the blood cultures as the “intervention” group and the other sites of infection as the “control” group. We considered the log odds ratio as a measure of the difference between resistance prevalence in these two groups. Fixed effect models gave very similar estimates to random effects models, so only the latter are presented, which assume that each year comes from a normal distribution centred on the average difference between the two sample types across the years. We also considered the degree of inconsistency/heterogeneity across years using I^2^ statistics.^3^ Similar to the methods above, these models estimate an overall fixed difference between the two sample types over all calendar years, but do allow individual years to vary around this.

We also conducted two different types of meta-regression. The first meta-regression was a standard extension of the meta-analysis above, modelling the log odds ratio for resistance in blood vs other sample types (with its standard error) as the response variable, and including year as an explanatory variable to assess whether there was an evidence of trends over calendar time in the difference between sample types in resistance prevalence – for example, rates rising faster or slower in bloodstream compared with other infections. The second directly modelled the log odds of resistance in bloodstream infections as the outcome variable (allowing for uncertainty), against the log odds of resistance in other sample types as the explanatory variable. Errors in the estimate of resistance in other sample types was assumed to be zero in these models (i.e. resistance prevalence was considered known exactly) given their very high numbers. The advantage is that, rather than assuming the same fixed difference between the proportion resistant in blood and other sample types as the standard meta-analysis above, meta-regression allows for the relationship between these proportions to vary. For example, the difference between them could be smaller (or larger) at high (or low) resistance prevalence.

**Supplementary Results**

**Supplementary Figures 2, 3, 4, 6 and 7** illustrate details of the analyses presented in the main text. For each antibiotic considered, the left panel illustrates trends in proportions of resistant bloodstream infections and other infections per year between1998-2018. These follow very similar trends. The middle panels show the resistance prevalences in the two sample types plotted against each other, with a meta-regression of resistance prevalence in blood cultures on resistance prevalence in other cultures (estimates shown in **Supplementary Table 1**; perfect agreement represented by an estimate of 1.00 with a very tight 95% CI). As the relationship was modelled as linear in the log odds, transforming back to the proportion scale leads to a slight curvature in the association. **Supplementary Table 2** shows the number of drug-years where the proportions of resistant bloodstream infections lay within the 95% prediction interval based on the proportion of resistant other infections (which also takes into account variability in this estimate). The third panel in these Supplementary Figures is a forest plot of the meta-analysis of the log odds ratio for resistance in bloodstream vs other cultures over study years, with variability between the pooled odds ratio under the assumption of a random effects model and estimates from the individual years represented by the I^2^ statistic. For perfect agreement the point estimate from this meta-analysis would have contained 1 (or at least the 95% CI contained 1). The meta-regression on calendar year in **Supplementary Table 1** assesses evidence for variation in this odds ratio between blood and other isolates over calendar time.

In **Supplementary Table 1**, the cross-correlation function reflects the association between the two time series shown in left panels of the Supplementary Figures above, allowing for different lags between the two time series; for example, a lag of 1 year is the cross-correlation between the proportion of resistant urines in one year and proportion of resistant blood cultures in the following year. The maximum cross-correlation between the proportion of resistant bloodstream infections and the proportion of resistant urinary tract infections is shown for each pathogen-drug combination – generally these occurred at lag 0, that is, no displacement.

***Individual-level analyses***

One reason for less than perfect agreement between resistance prevalence in blood and other cultures could be that different patients are included in the two estimates, which are simply based on all bloodstream isolates and all isolates from other samples, regardless of patient population (since this total dataset would be simple to obtain from a microbiology laboratory). In contrast, here we focus on using susceptibility results in other sites of infections to predict resistance in bloodstream infections within the same patient – the major disadvantage of this approach is reduced numbers, but it enables an assessment of the impact of population sampling on our main results.

Of the total 8102 *E. coli* bloodstream infections (not all with susceptibility test results for all antibiotics, hence denominators are slightly smaller in individual figures), only 2144 (26%) had a *E. coli* urinary tract infection within [-3,+2] days of the blood culture **(Supplementary Figure 1**), and only 1139 (14%) had one within [-90,-3] days. Similarly of 6952 *S. aureus* bloodstream infections, only 1820 (26%) had a *S. aureus* skin surface/genital/urine culture within [-3,+2] days of the blood culture. The percentages were much higher when considering *S. aureus* from other clinical cultures with 4188 (60%) of the *S. aureus* bloodstream infections having one within [-3,+2] days.

In **Supplementary Table 5**, we present contingency tables for all antibiotics for *E. coli* and *S. aureus*, together with performance metrics which are classically used when a replacement diagnostic tool is tested on the same sample (that is, using paired data).

Focusing on the performance of using susceptibility results of the closest *E. coli* urinary tract infection taken at any time from 3 days before up to 2 days after the bloodstream infection isolate within the same patient, interestingly, even on this paired data, resistance to trimethoprim was higher in urinary than blood isolates for trimethoprim (McNemar p<0.0001), but was lower in urinary than blood isolates for amoxicillin (p=0.018) and co-amoxiclav (p<0.0001) (similarly to results in all isolates described above). There was no asymmetry in resistance for ciprofloxacin (p=0.33) with high overall agreement (99%). Guidance on susceptibility testing tools^4^ require a major error (ME) of less than 3% and a very major error (VME) of less than 7.5% (upper 95% confidence limit for the true VME) which were only satisfied for ciprofloxacin when considering *E. coli*. But, for *E. coli* at least, there was no evidence that the modestly different resistance rates seen in blood and urine isolates in the overall analysis were due to differential sampling of intrinsically different underlying populations.

We next considered the concordance of susceptibility results from the closest prior *E. coli* urinary tract infection taken between 3 and 90 days before the bloodstream infection within a patient, since even within a high-income country, this could be a potential indicator of inappropriate empiric therapy. Whilst resistance to amoxicillin and co-amoxiclav was still lower in urinary than blood isolates (McNemar p<0.0001), interestingly there was no difference in trimethoprim resistance between prior urinary isolates and current blood isolates (p=0.69) potentially implicating inadequate treatment of a prior urinary tract infection with trimethoprim as contributing to bloodstream infection. Diagnostic performance was lower for all antibiotics compared with concurrent isolates described above; however, positive predictive value (PPV) was 80% or higher for all drugs including co-amoxiclav, meaning that four out of five patients with a prior co-amoxiclav resistant urinary tract infection would turn out to have a co-amoxiclav resistant bloodstream infection.

For *S. aureus* overall accuracy between resistance from concurrent blood and non-blood cultures was even higher than for *E. coli*, at (97-99%) with ciprofloxacin, erythromycin and oxacillin having VME and ME under 3% when looking at other clinical cultures, and only slightly higher values when considering only non-sterile site cultures (VME under 5% and ME under 4%). For these three drugs there was some evidence of asymmetry (McNemar p-values between 0.0067-0.01), but numbers were small, and if anything resistance was higher in non-blood cultures. The ME for gentamicin and tetracycline were only 1%, however the VME were 29% and 8% respectively.

1.	Walker S, Peto TEA, O’Connor L, Crook DW, Wyllie D. Are there better methods of monitoring MRSA control than bacteraemia surveillance? An observational database study. PLoS One [Internet]. 2008;3(6):e2378. Available from: http://www.pubmedcentral.nih.gov/articlerender.fcgi?artid=2405929&tool=pmcentrez&rendertype=abstract

2.	Lin LI. A Concordance Correlation Coefficient to Evaluate Reproducibility. Biometrics. 1989;45(1):255–68.

3.	Higgins JPT, Thompson SG, Deeks JJ, Altman DG. Measuring inconsistency in meta-analyses. BMJ [Internet]. 2003;327(7414):557–60. Available from: http://www.ncbi.nlm.nih.gov/pubmed/12958120%0Ahttp://www.pubmedcentral.nih.gov/articlerender.fcgi?artid=PMC192859

4.	U.S. Department of Health and Human Services Food and Drug Administration Center for Devices and Radiological Health. Guidance for Industry and FDA: Class II Special Controls Guidance Document : Antimicrobial Susceptibility Test ( AST ) Systems. 2009;1–42.

Supplementary Figure 1 Venn diagram of E. coli urinary tract infections, E. coli bloodstream infections, S. aureus non-sterile site cultures, S. aureus non-blood cultures and S. aureus bloodstream infections, Oxfordshire 1998-2018

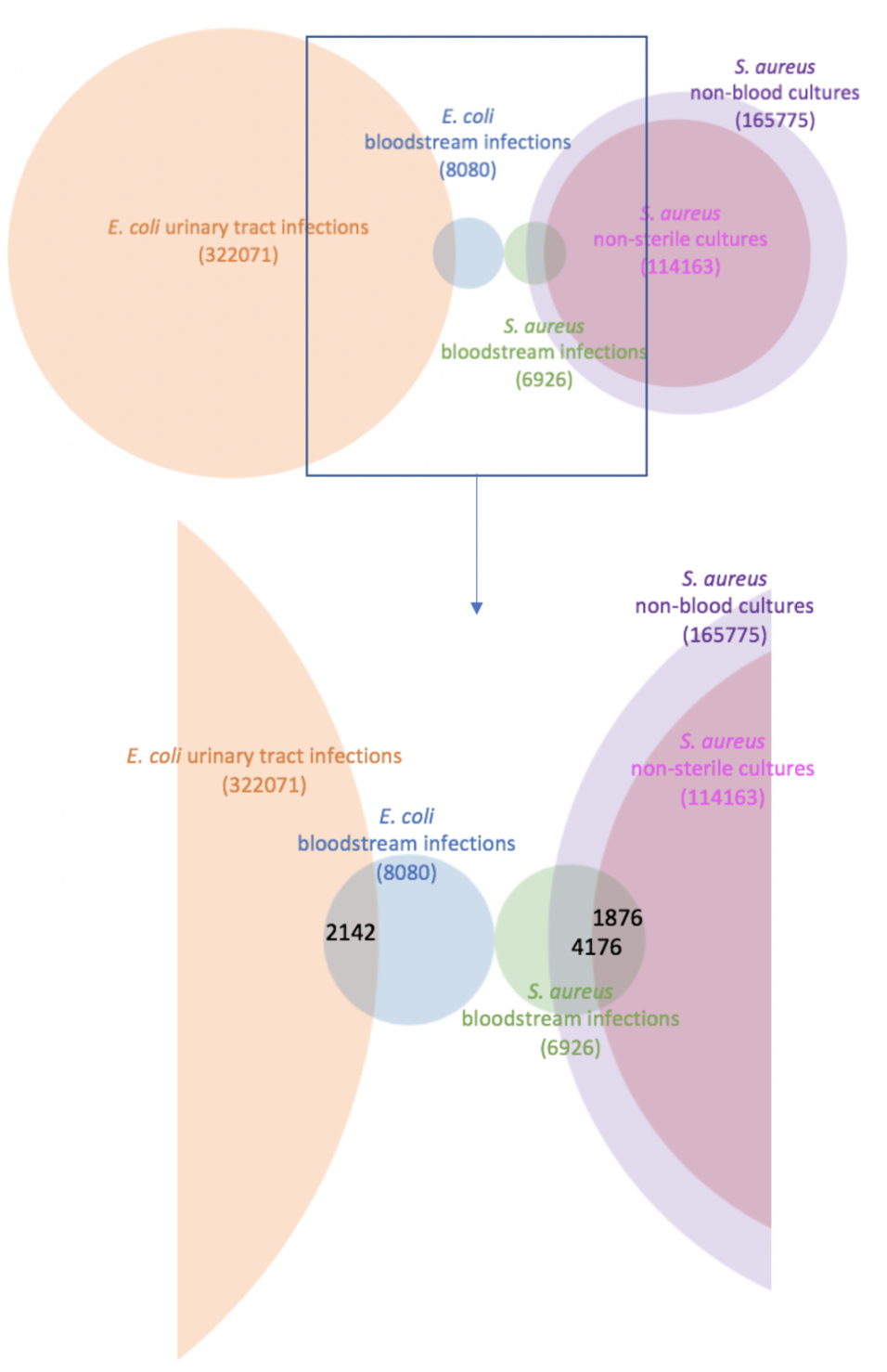

8102

**166179**

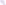

**112074**

**166179**

**4188**

**1820**

**6952**

**112074**

**6952**

**2144**

322087

8102

322087

Supplementary Figure 2 Agreement between proportion of resistant E. coli blood and urine cultures, for amoxicillin, co-amoxiclav, ciprofloxacin and trimethoprim, Oxfordshire 1998-2018

Amoxicillin resistance

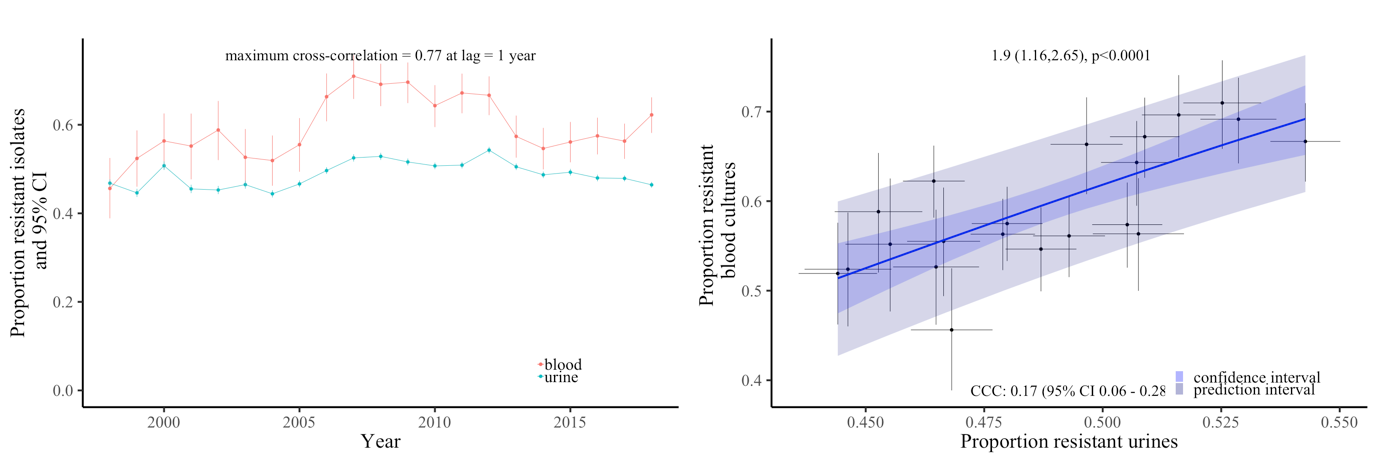

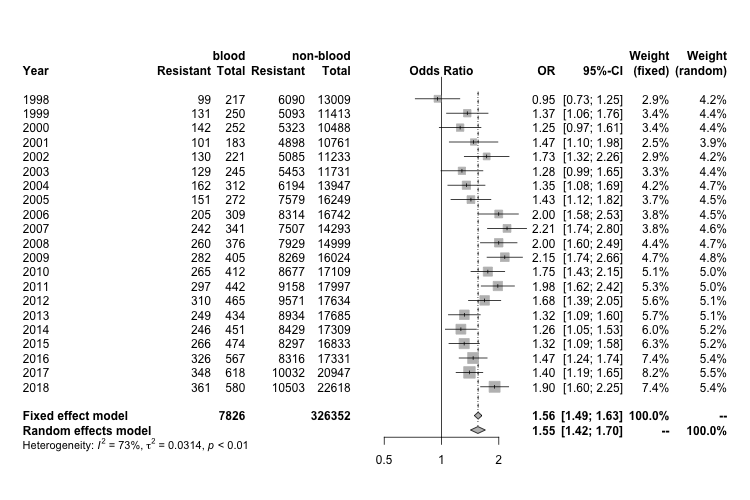

Co-amoxiclav resistance

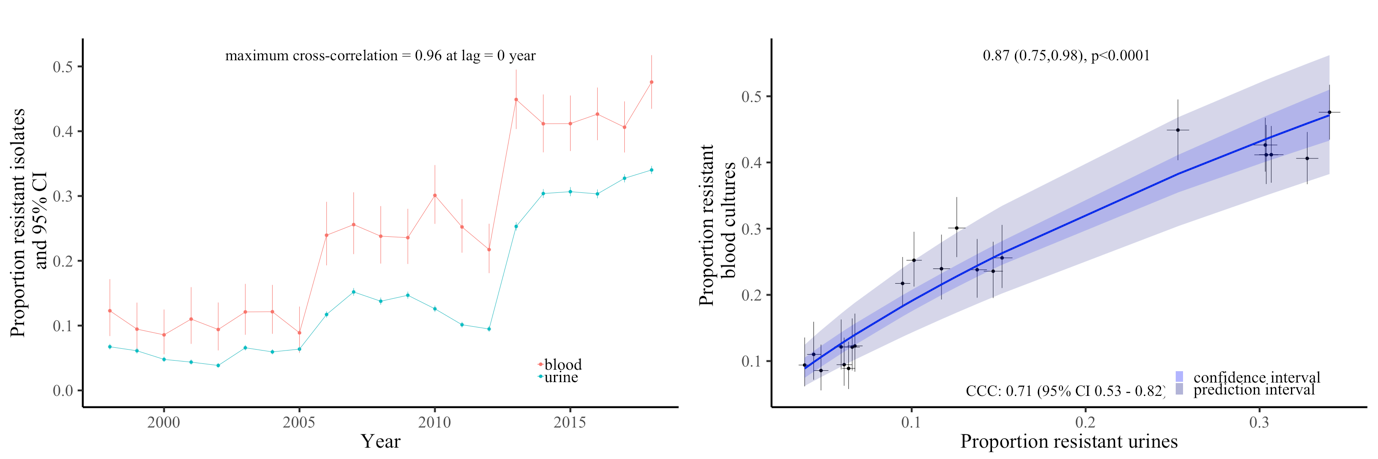

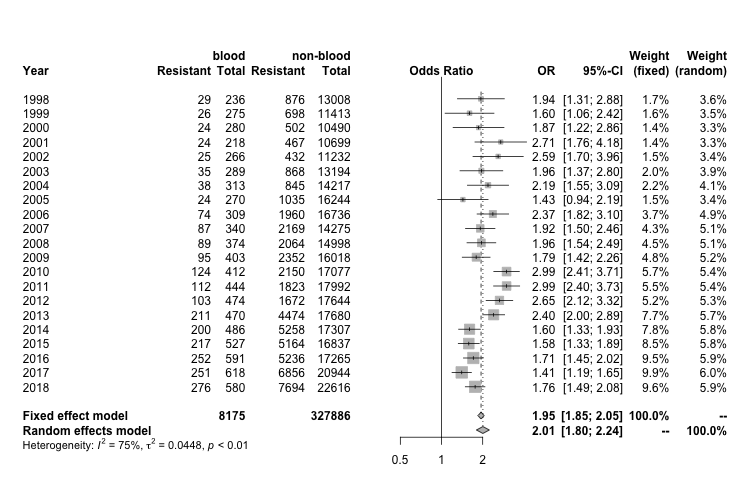

Note: In the middle plot the x and y axes are on different scales.

Ciprofloxacin resistance

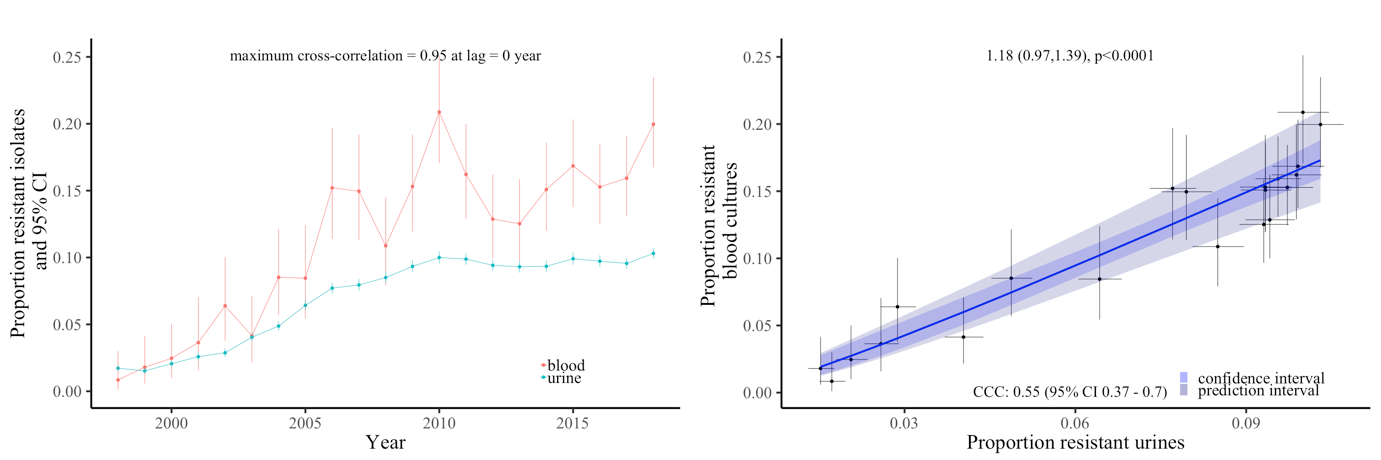

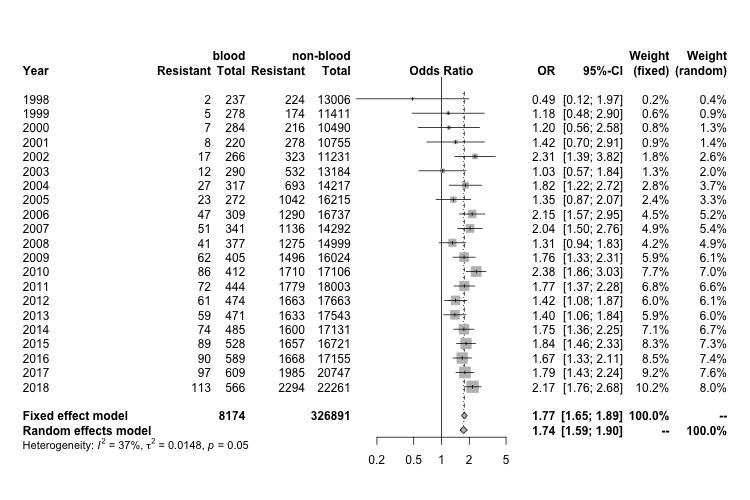

Trimethoprim resistance

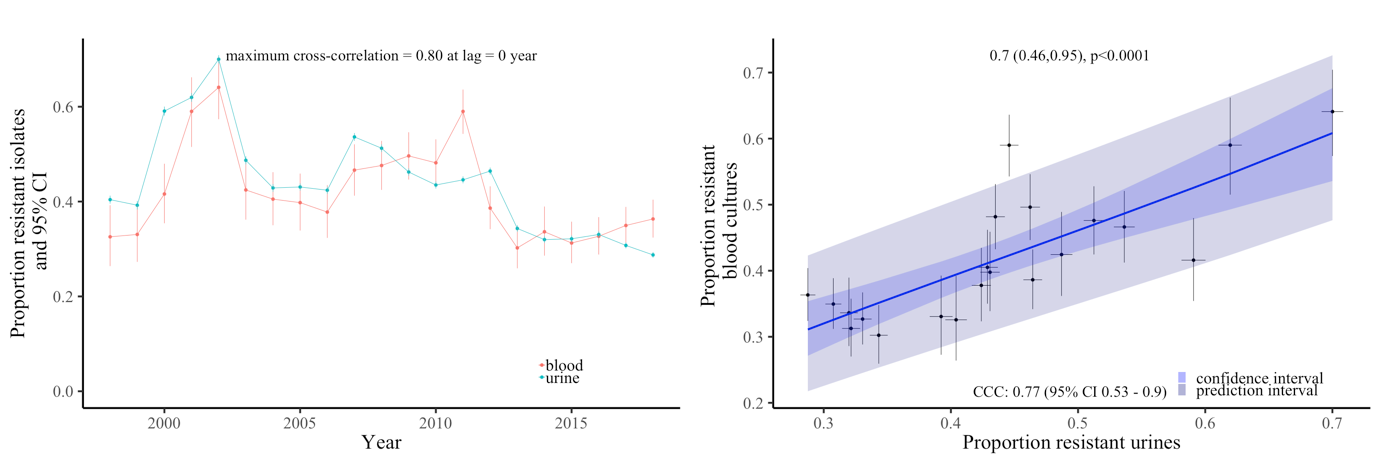

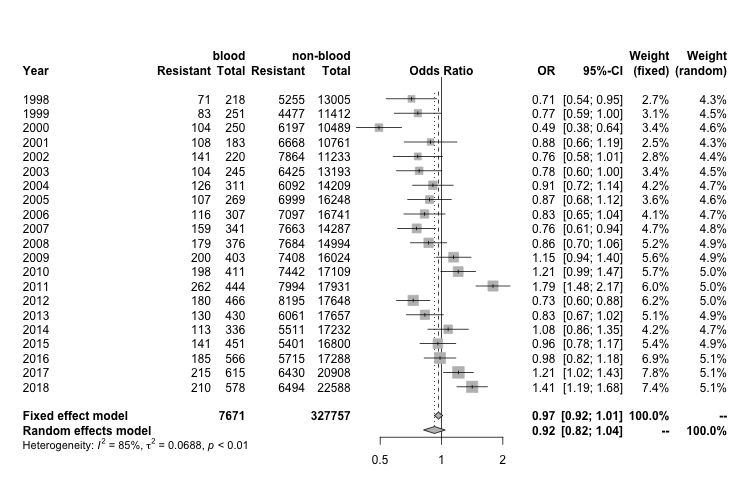

Note: In the middle plot the x and y axes are on different scales.

Supplementary Figure 3 Resistance prevalence in E. coli (left-hand panel) and S. aureus (right-hand panel) in blood and urine, respectively non-sterile site cultures in Oxfordshire, for antibiotics tested only 2013-2018

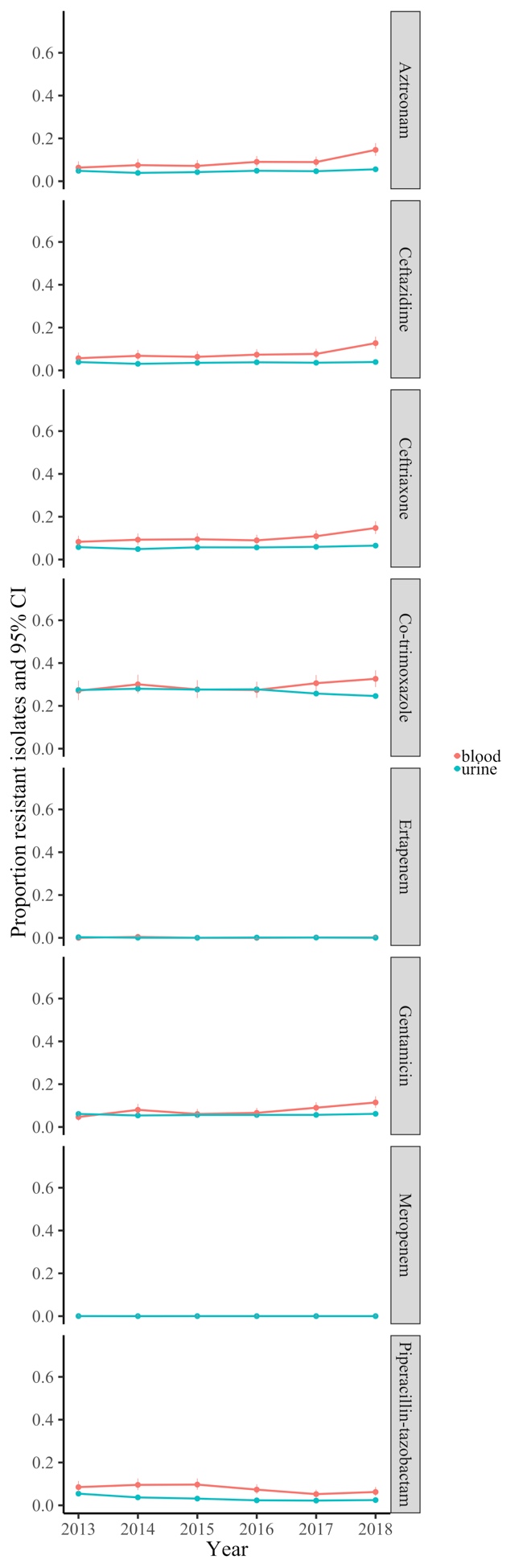

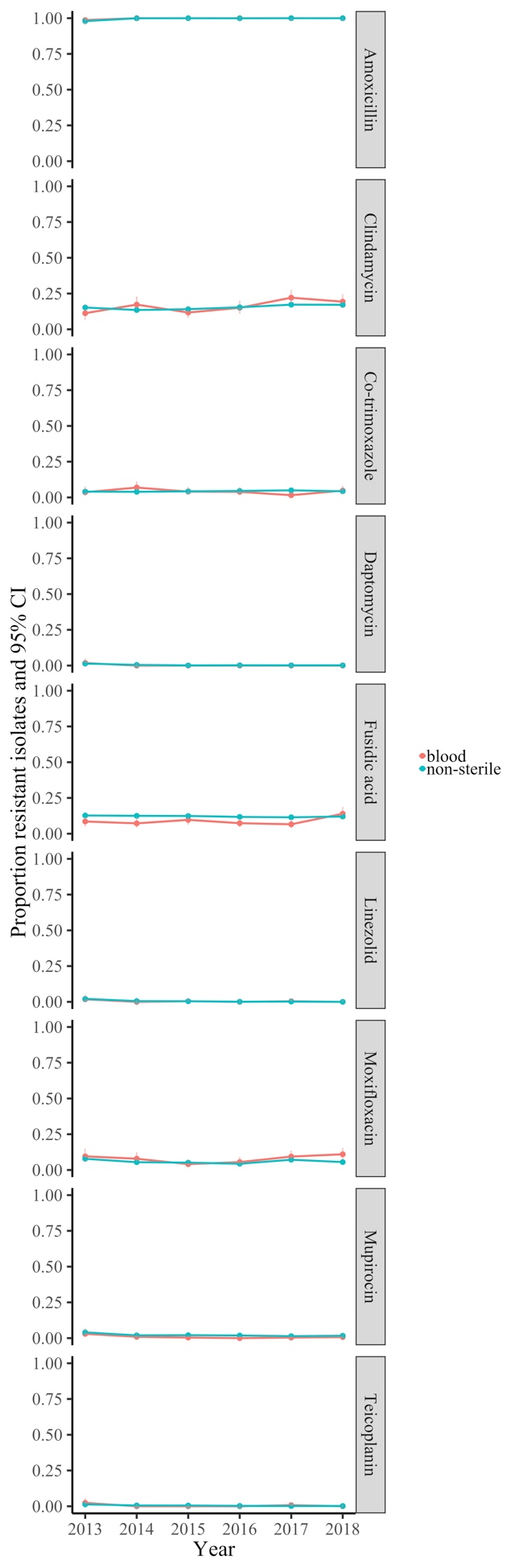

Supplementary Figure 4 Agreement between proportion of resistant Klebsiella bloodstream infections and proportion of resistant Klebsiella urinary tract infections, for co-amoxiclav and ciprofloxacin

Co-amoxiclav resistance

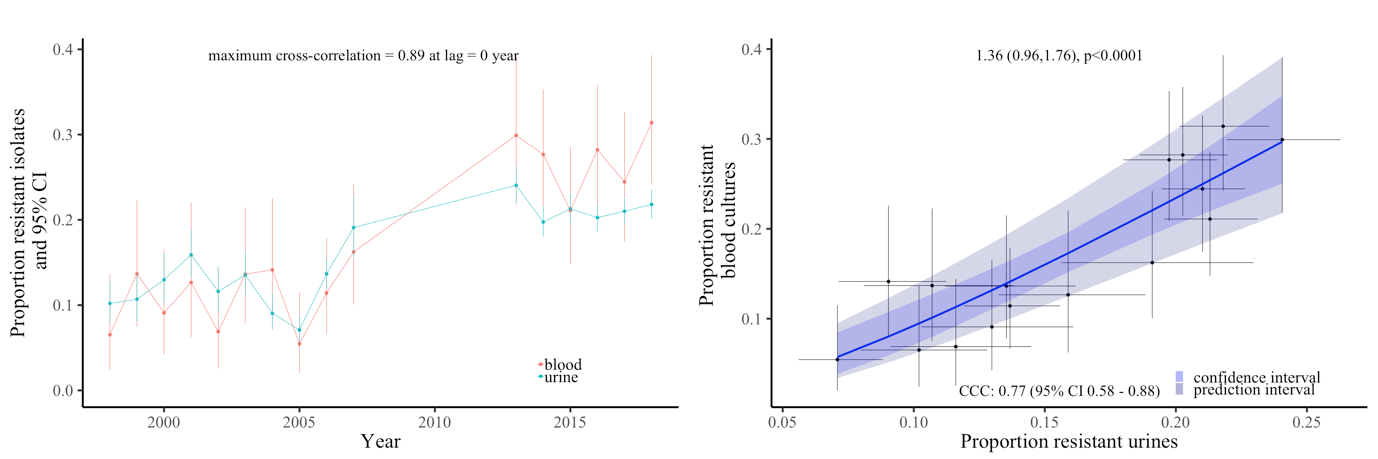

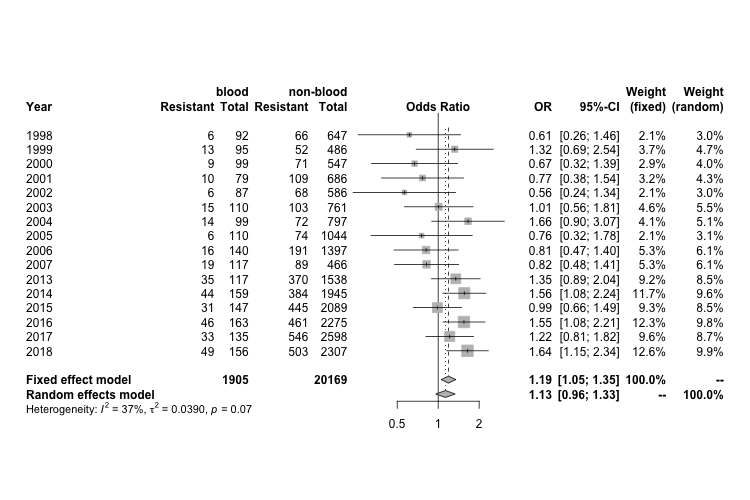

Note: In the middle plot the x and y axes are on different scales.

Ciprofloxacin resistance

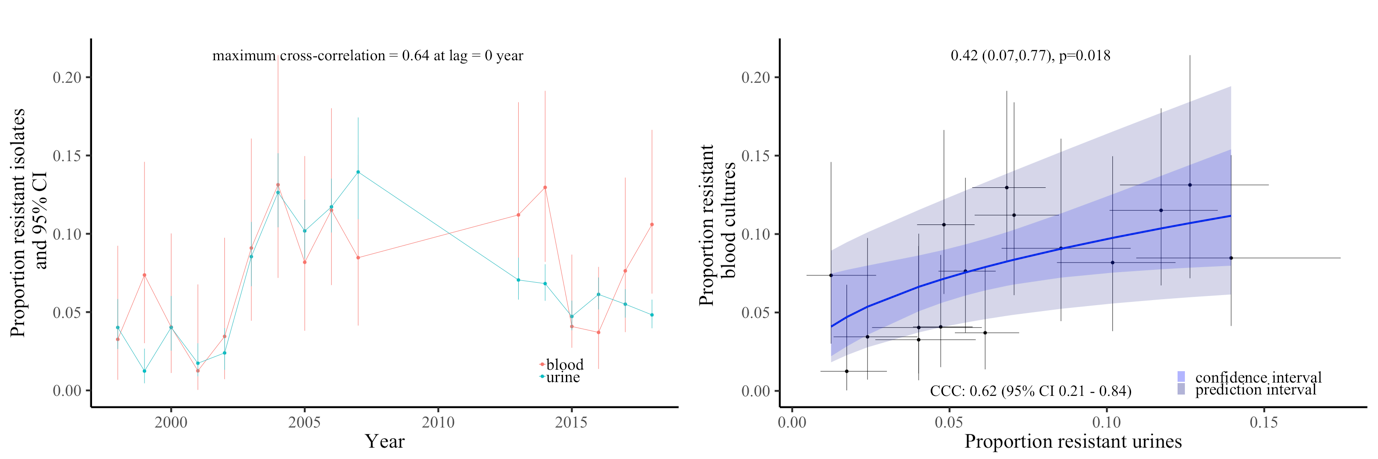

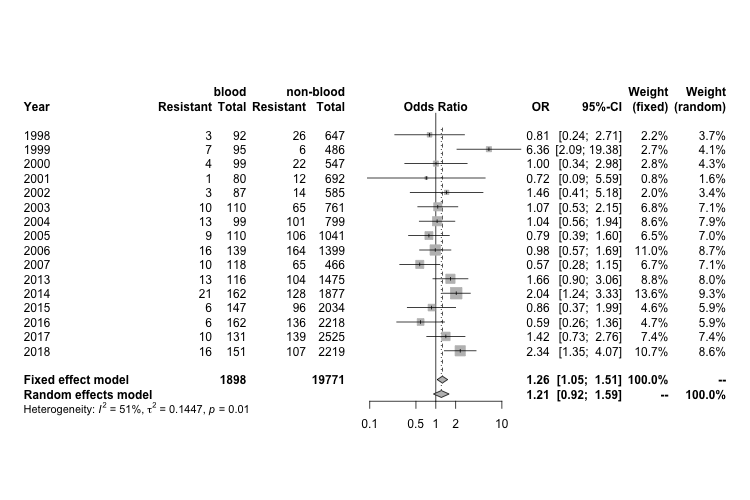

Supplementary Figure 5 Resistance prevalence in Klebsiella spp. in blood and non-blood cultures in Oxfordshire, 1998-2018

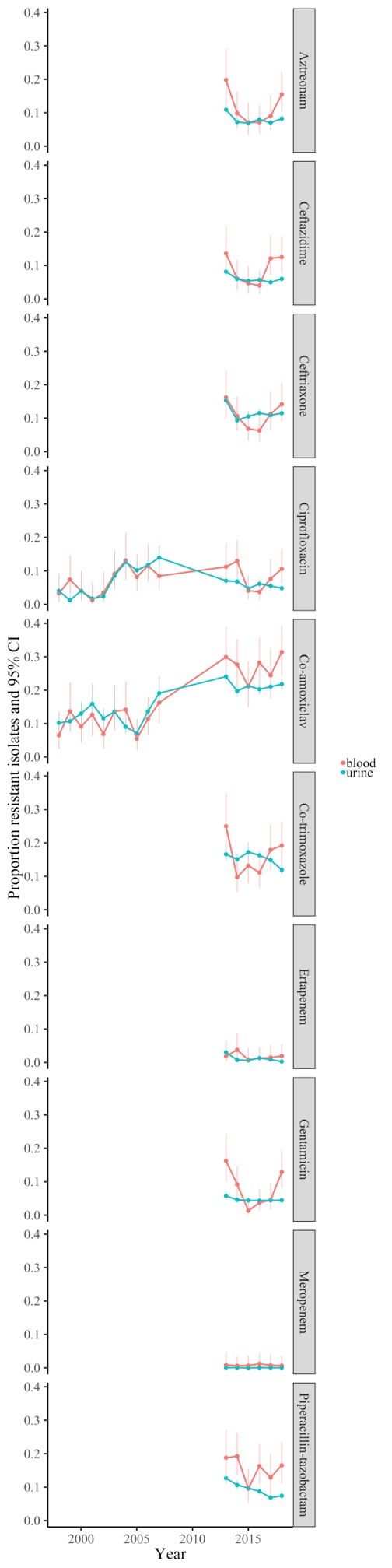

Supplementary Figure 6 Agreement between proportion of resistant S. aureus blood and non-sterile site cultures, for ciprofloxacin, erythromycin, oxacillin, gentamicin and tetracycline, Oxfordshire 1998-2018

Ciprofloxacin resistance

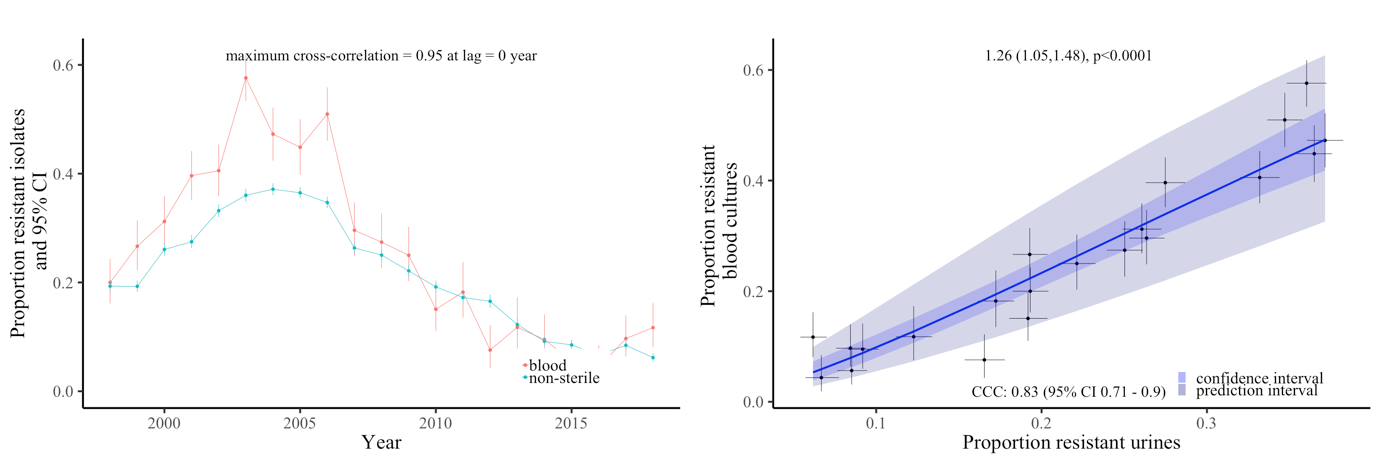

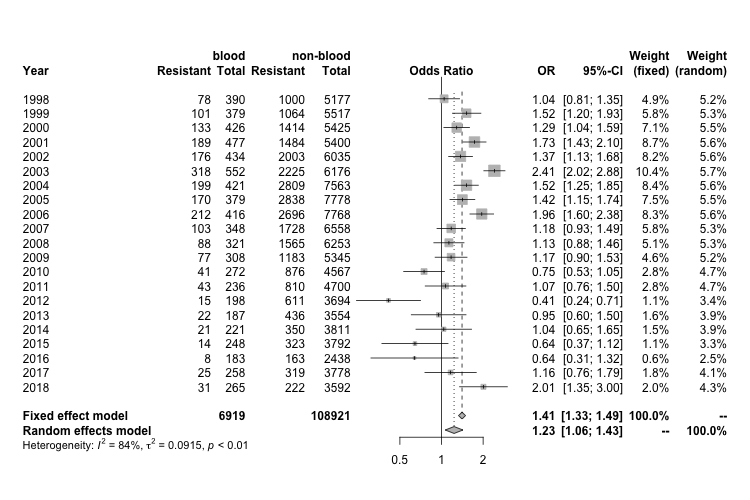

Erythromycin resistance

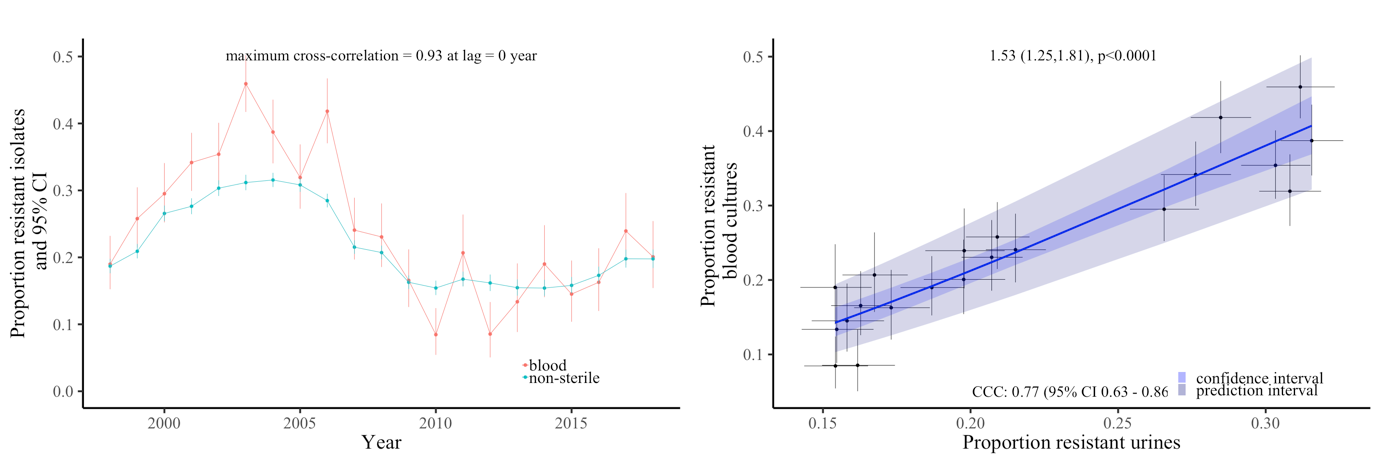

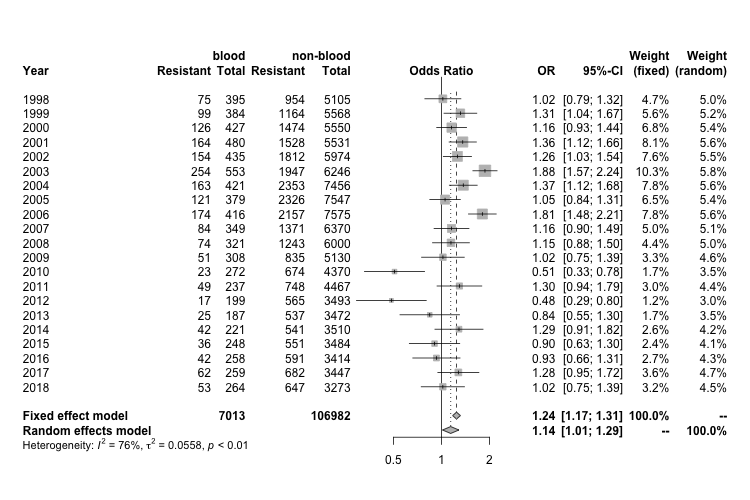

Note: In the middle plot the x and y axes are on different scales.

Gentamicin resistance

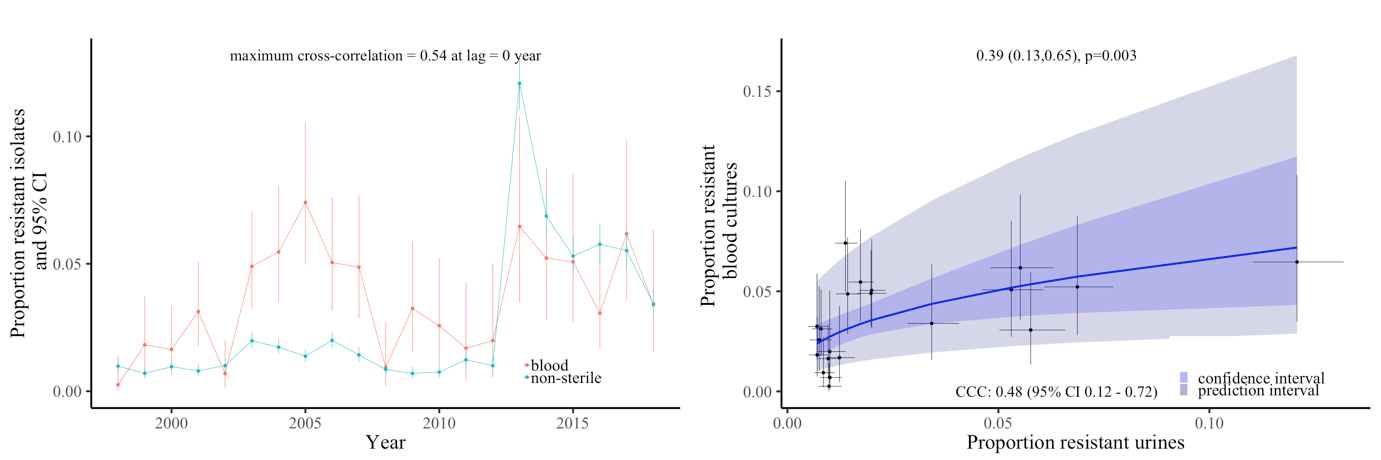

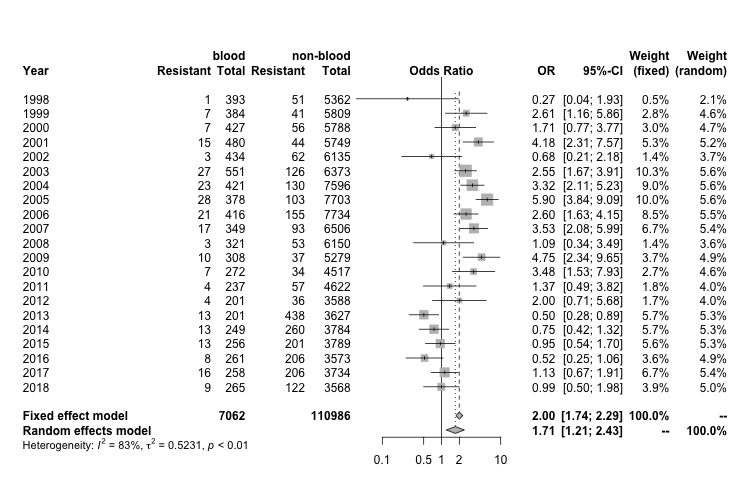

Oxacillin resistance

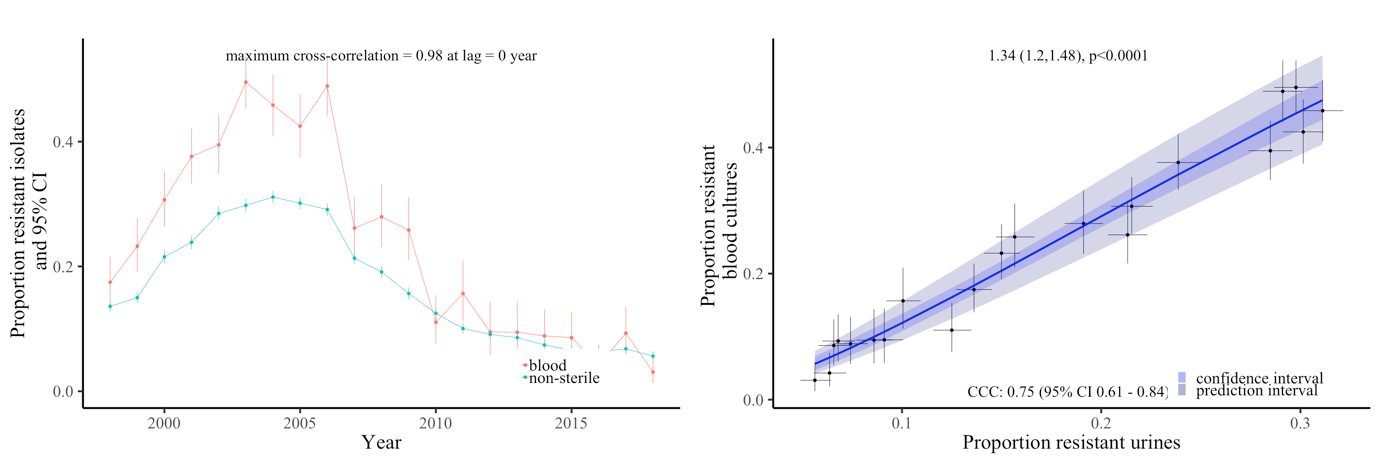

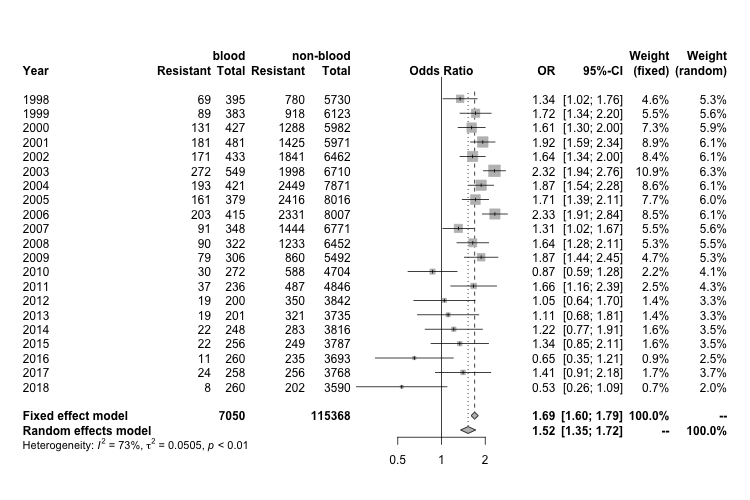

Note: In the middle plot the x and y axes are on different scales.

Tetracycline resistance

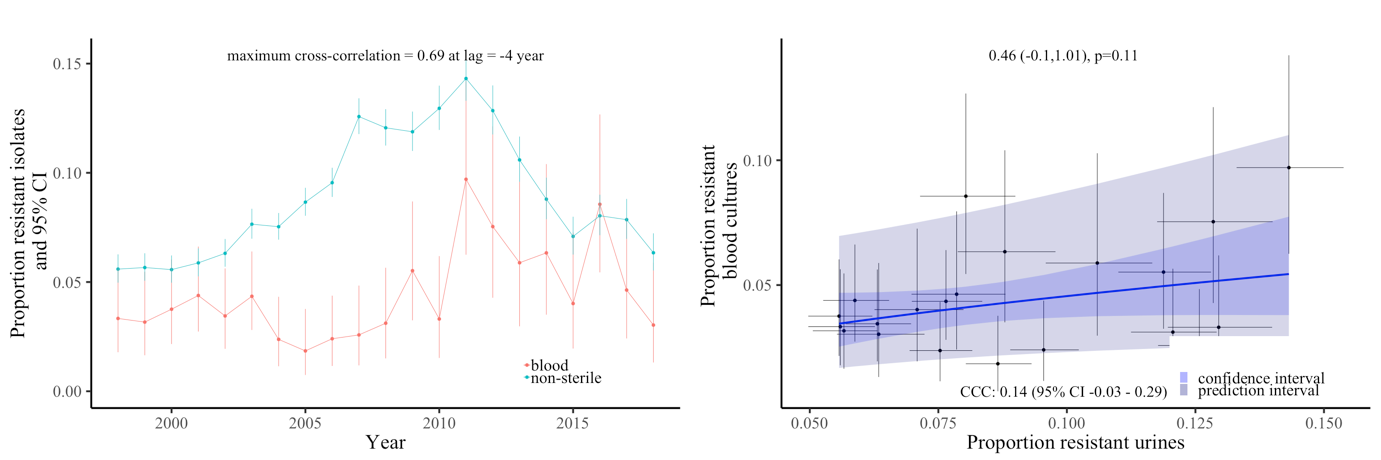

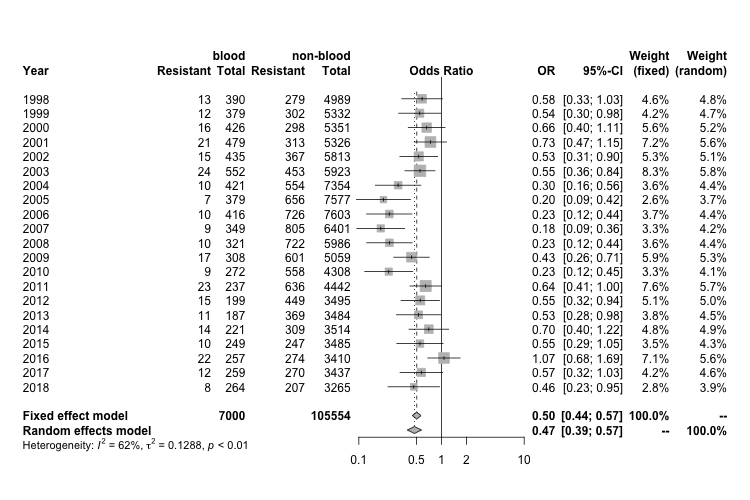

Note: In the middle plot the x and y axes are on different scales.

Supplementary Figure 7 Agreement between proportion of resistant S. aureus blood and, non-sterile site and sterile site cultures respectively, for erythromycin, gentamicin and oxacillin, Oxfordshire 1998-2018

(A) Erythromycin

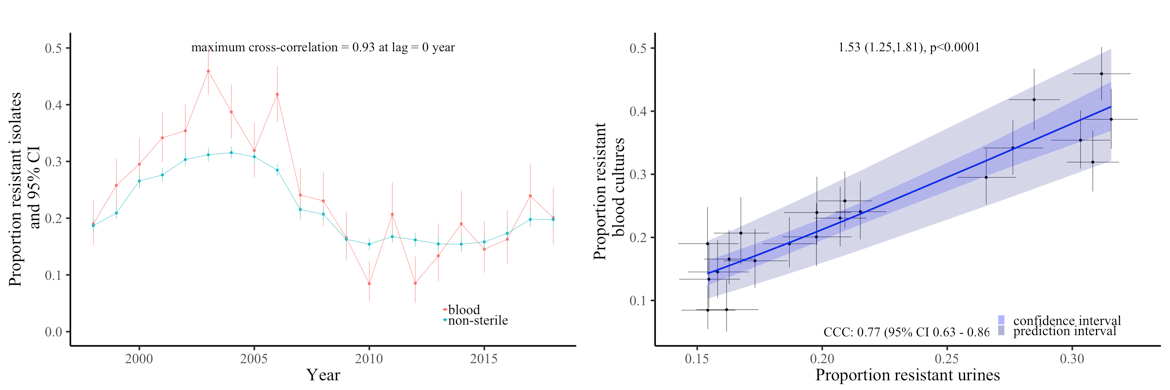

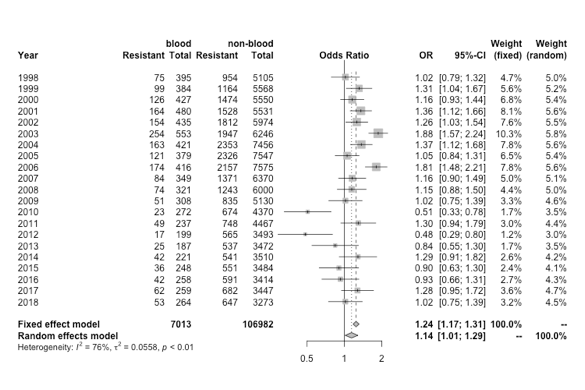

*S. aureus*

erythromycin-resistant blood cultures

vs

skin surface/genital/urine cultures

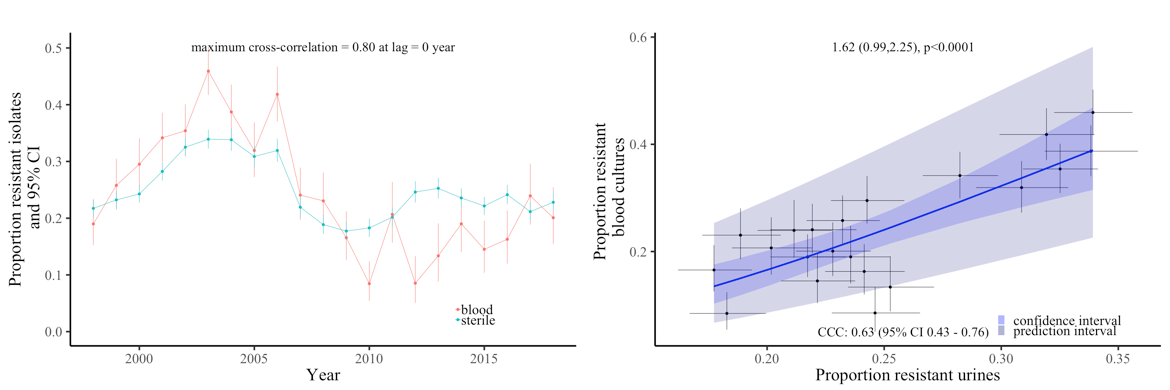

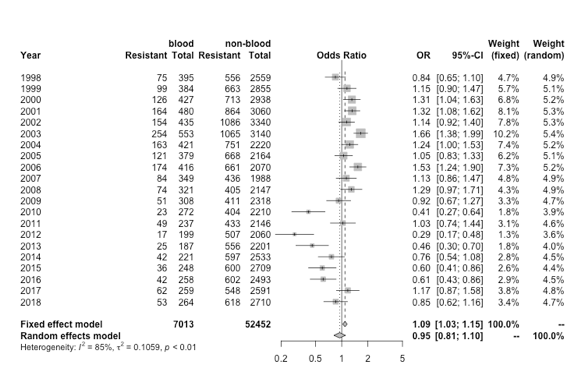

*S. aureus*

erythromycin-resistant blood cultures

vs

sterile/pus/wound/ tissue/respiratory

Note: In the middle plot the x and y axes are on different scales.

(B) Gentamicin

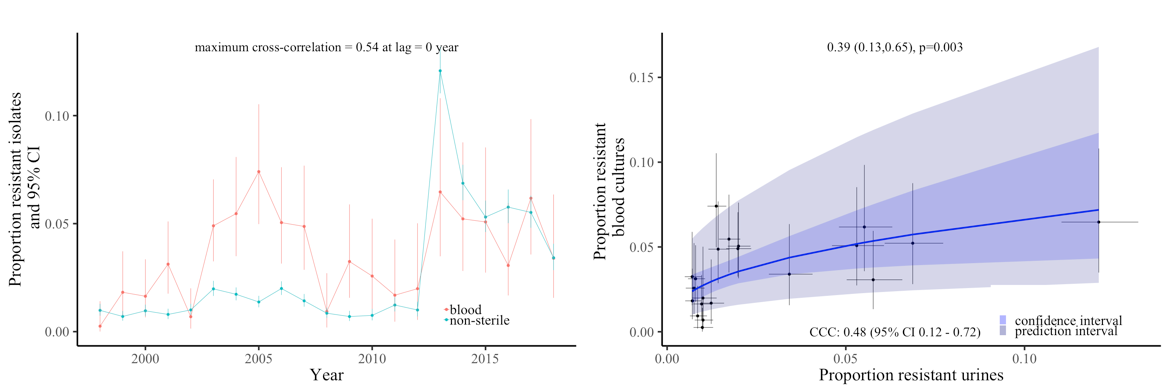

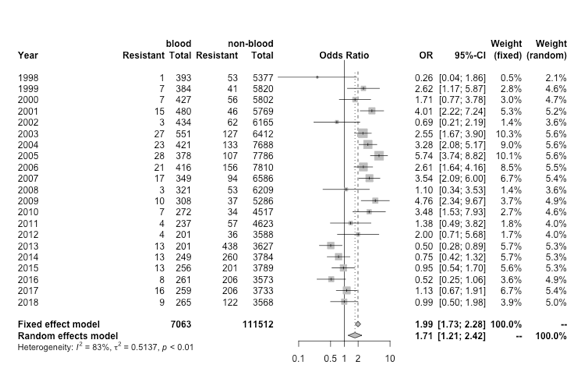

*S. aureus*

gentamicin-resistant blood cultures

vs

skin surface/genital/urine cultures

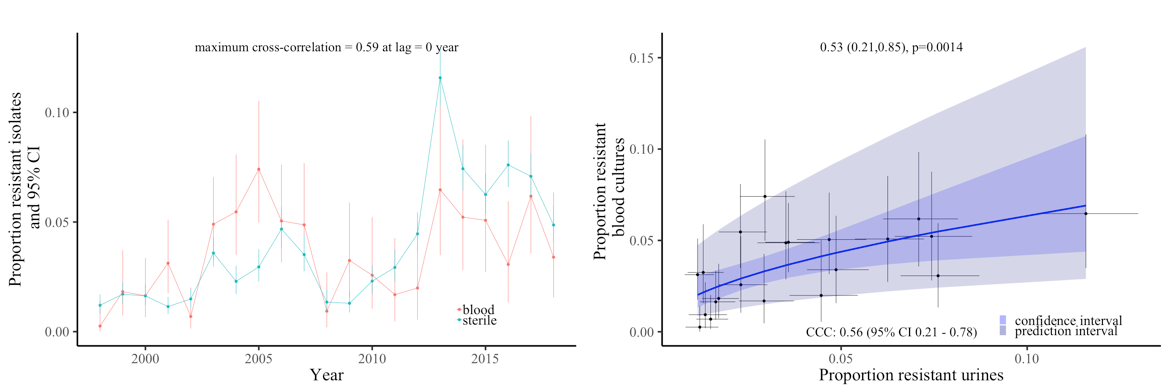

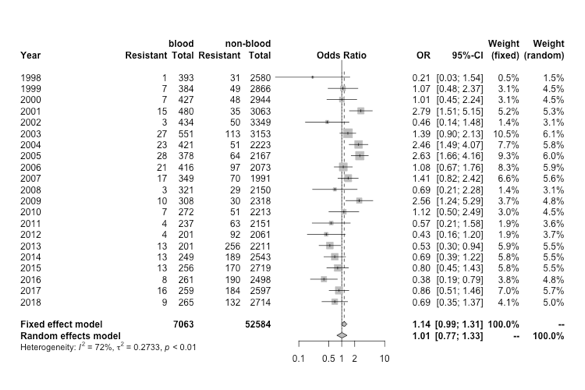

*S. aureus*

gentamicin-resistant blood cultures

vs

sterile/pus/wound/ tissue/respiratory

Note: In the middle plot the x and y axes are on different scales.

(C) Oxacillin

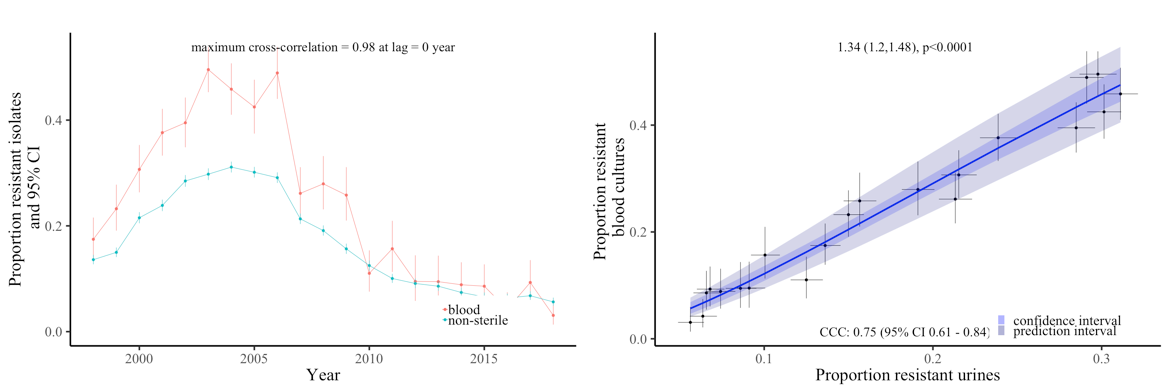

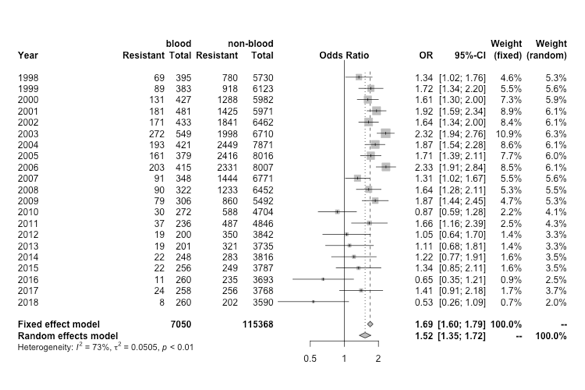

*S. aureus*

oxacillin-resistant blood cultures

vs

skin surface/genital/urine cultures

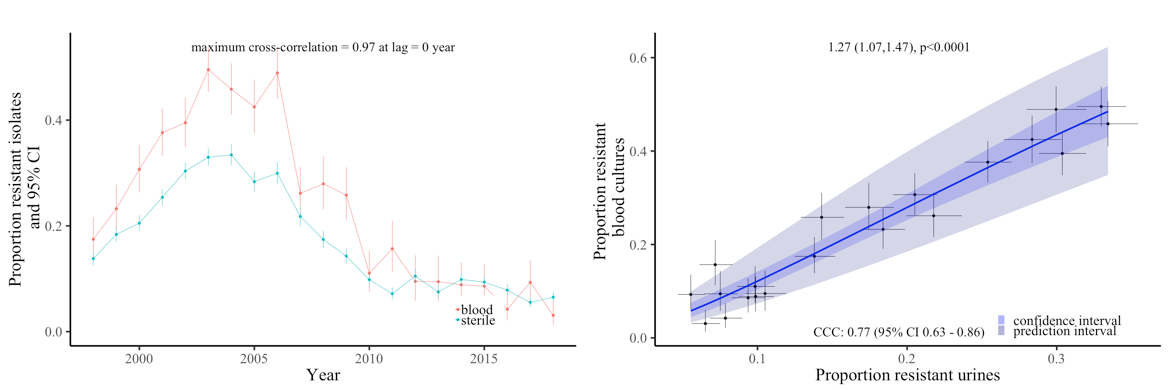

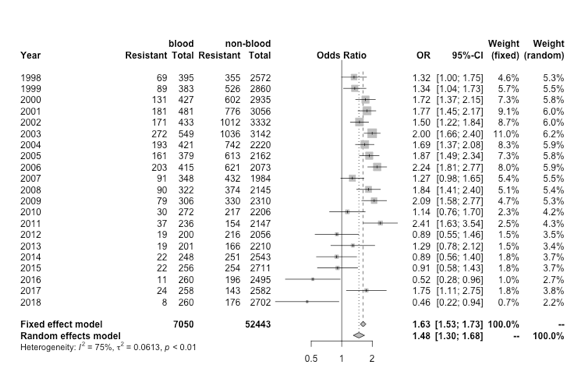

*S. aureus*

oxacillin-resistant blood cultures

vs

sterile/pus/wound/ tissue/respiratory

Note: In the middle plot the x and y axes are on different scales.

Supplementary Figure 8 Resistance prevalence in S. pneumoniae (left-hand panel), E. faecalis (middle panel), E. faecium (right-hand panel) in blood and non-blood cultures in Oxfordshire, 1998-2018

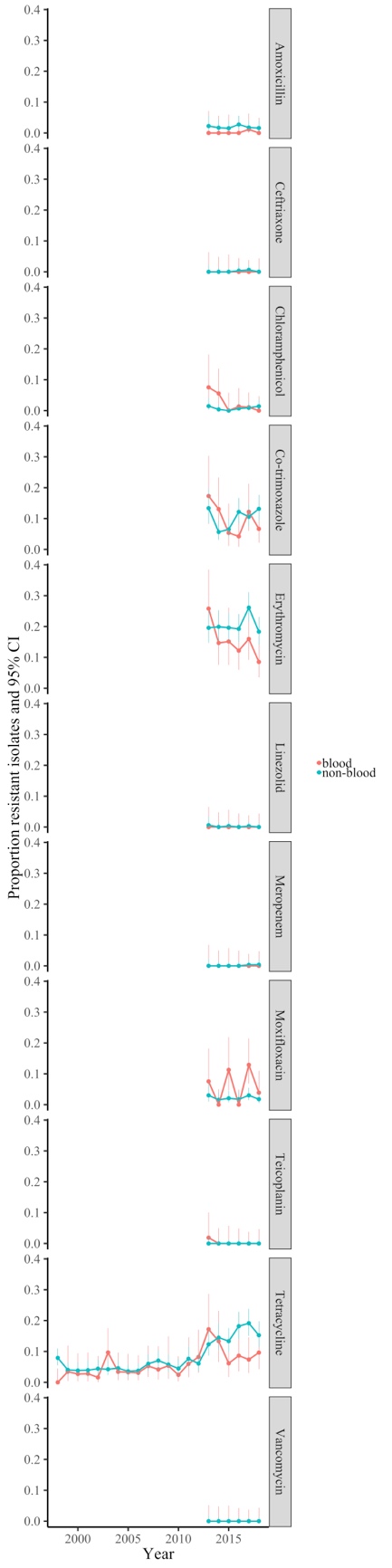

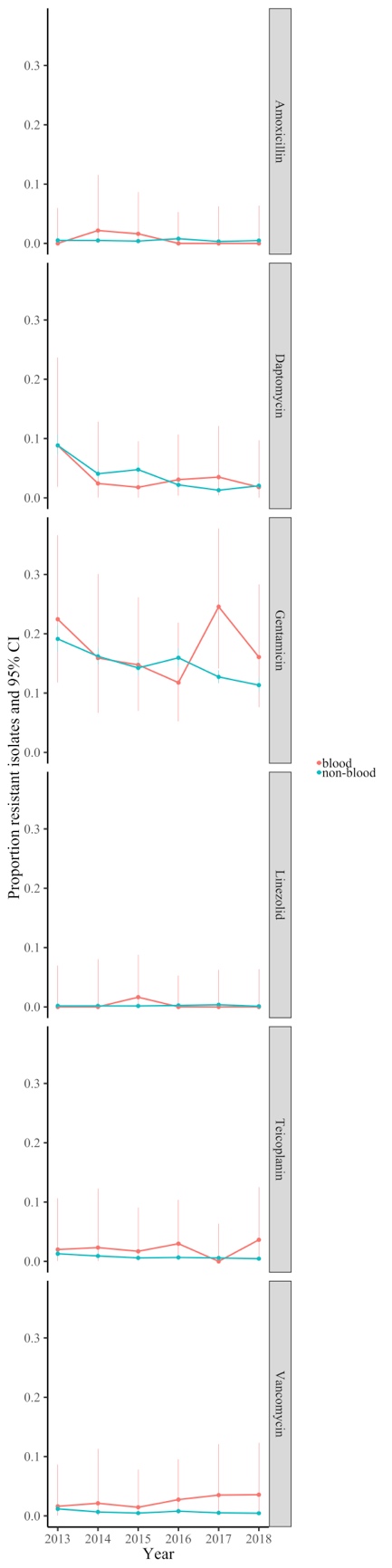

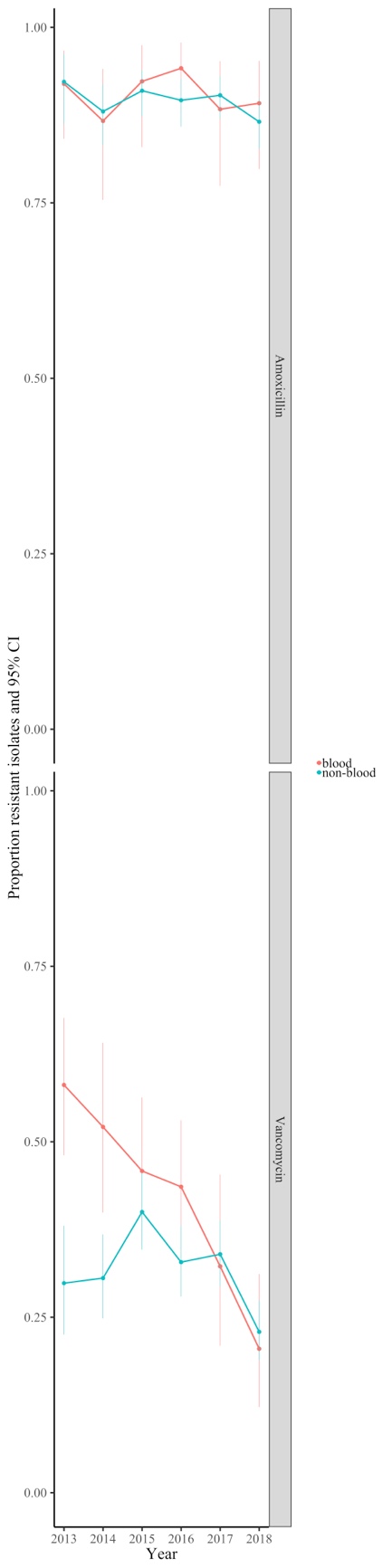

Supplementary Figure 9 Resistance prevalence in S. aureus in blood and non-blood cultures in high-income countries present in the ATLAS dataset

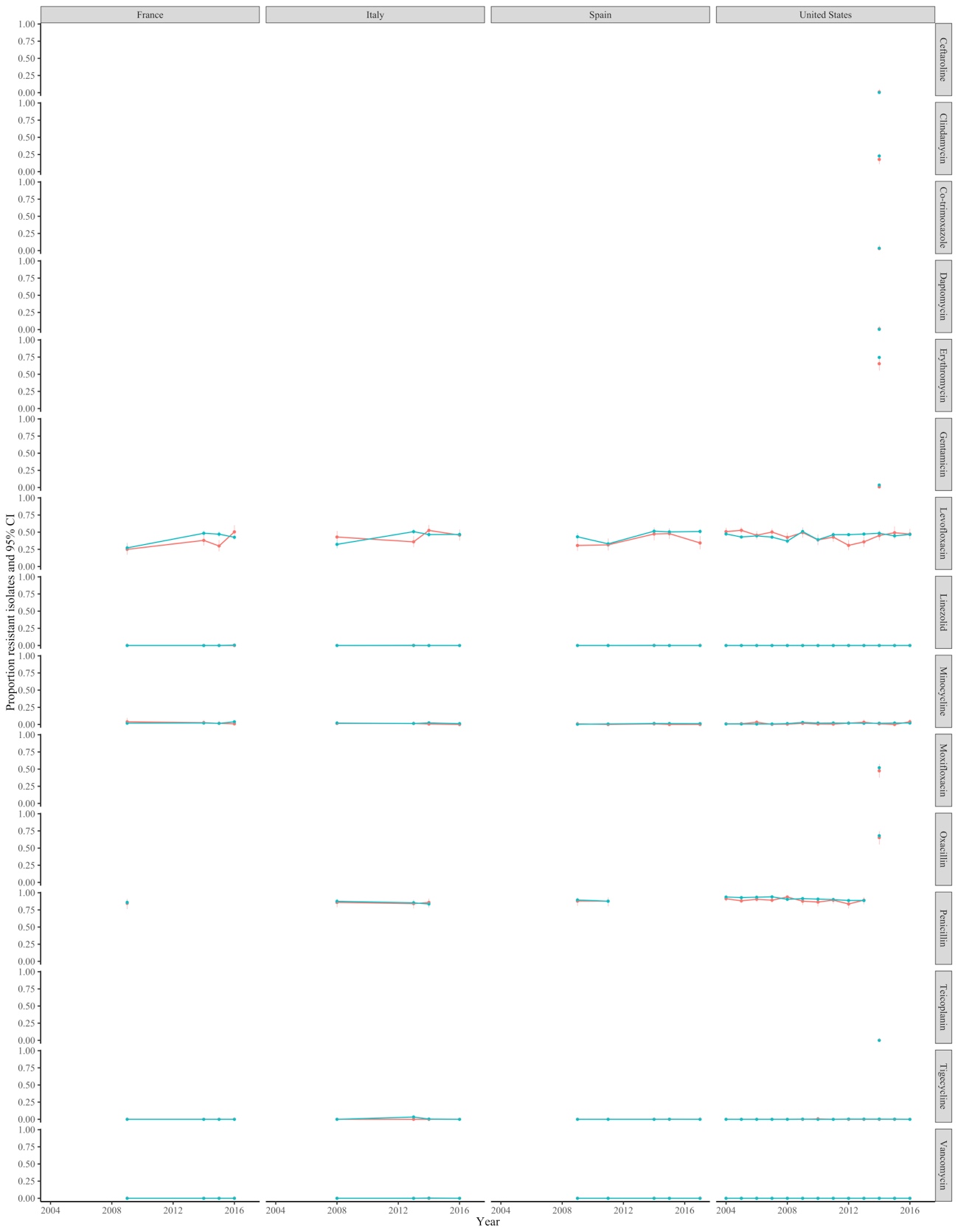

Supplementary Figure 10 Resistance prevalence in E. coli in blood and non-blood cultures in middle-income countries present in the ATLAS dataset

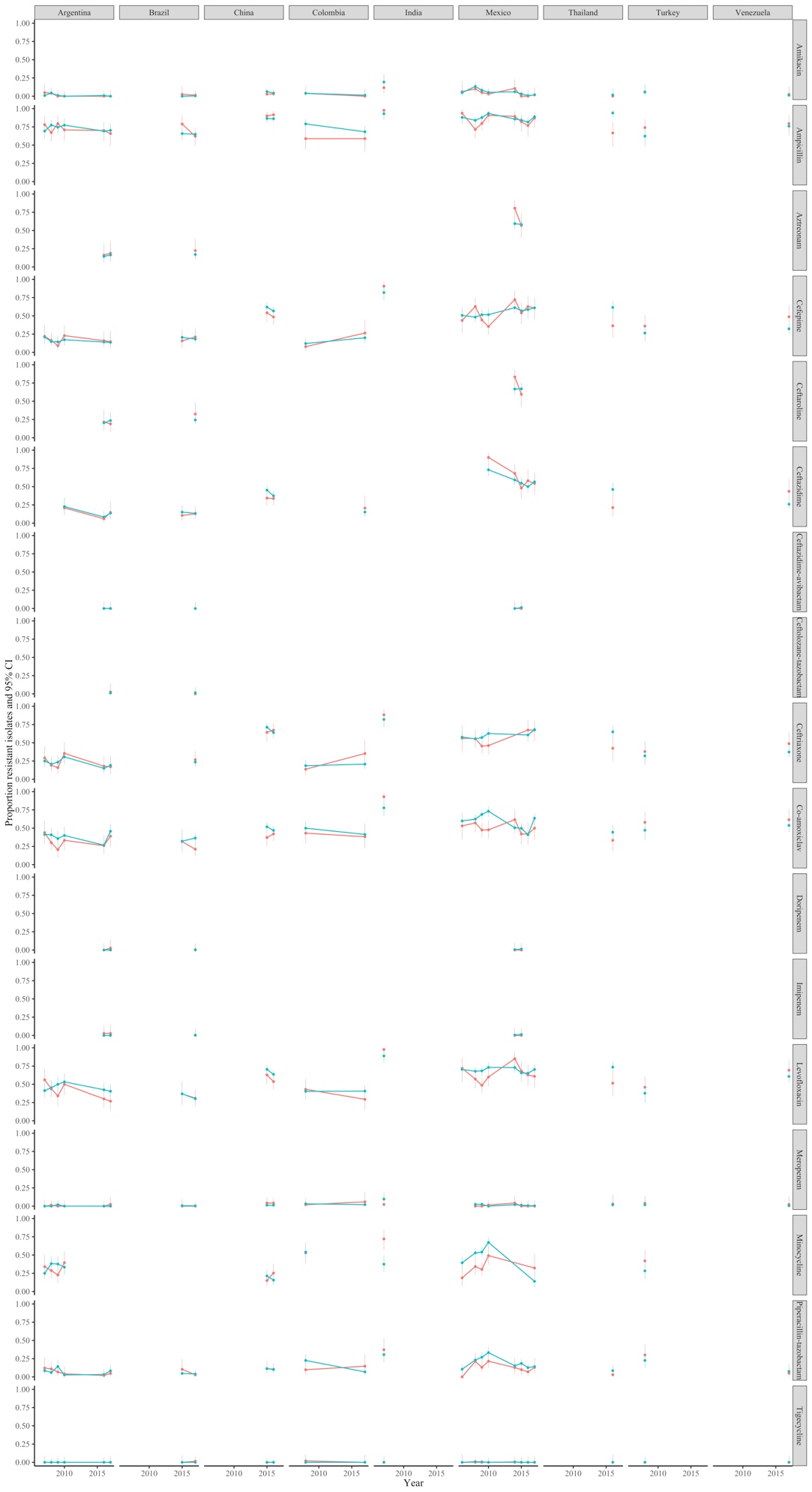

Supplementary Figure 11 Resistance prevalence in S. aureus in blood and non-blood cultures in middle-income countries present in the ATLAS dataset

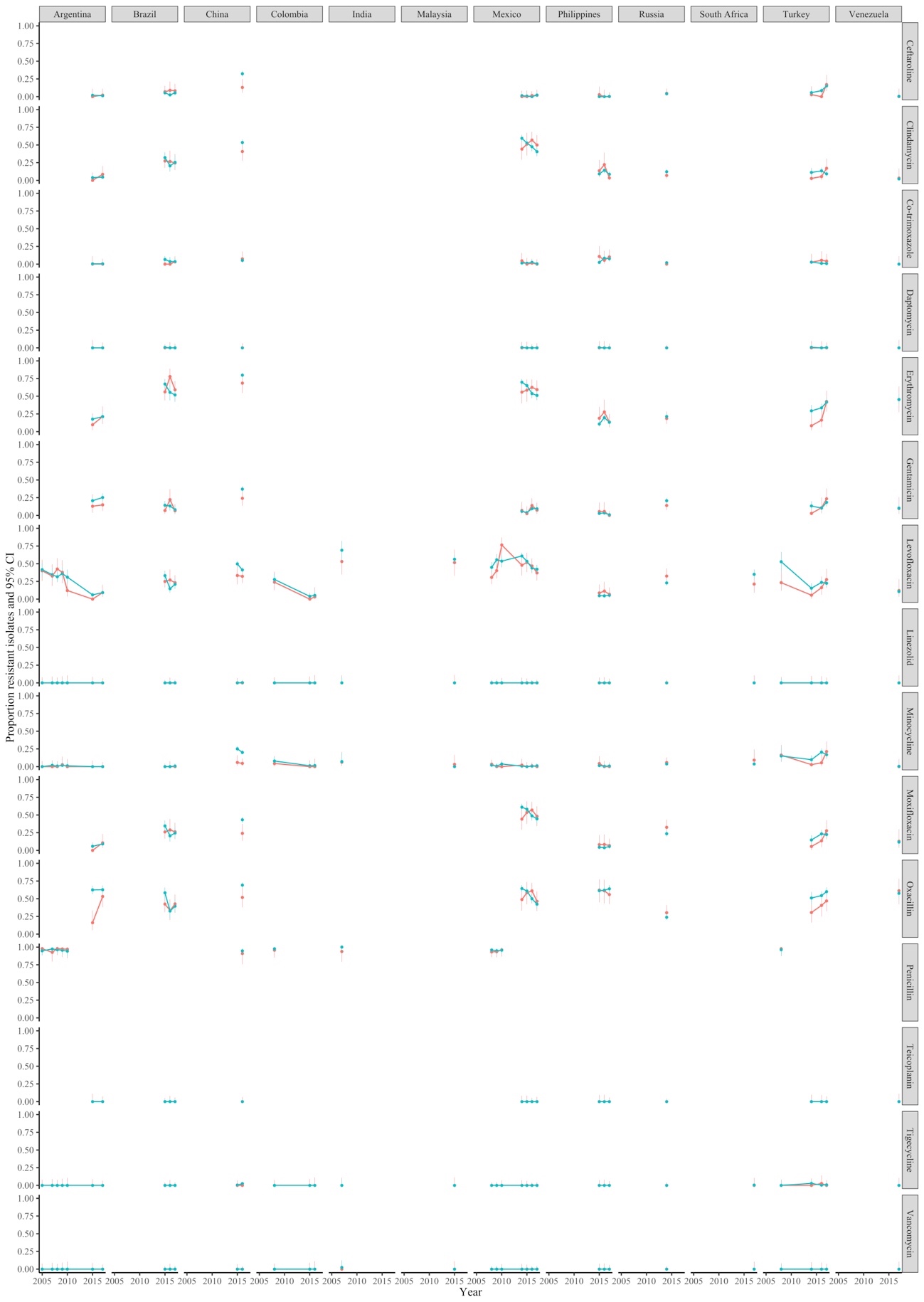

Supplementary Figure 12 Resistance prevalence in Klebsiella pneumoniae in blood and non-blood cultures in high-income countries present in the ATLAS dataset

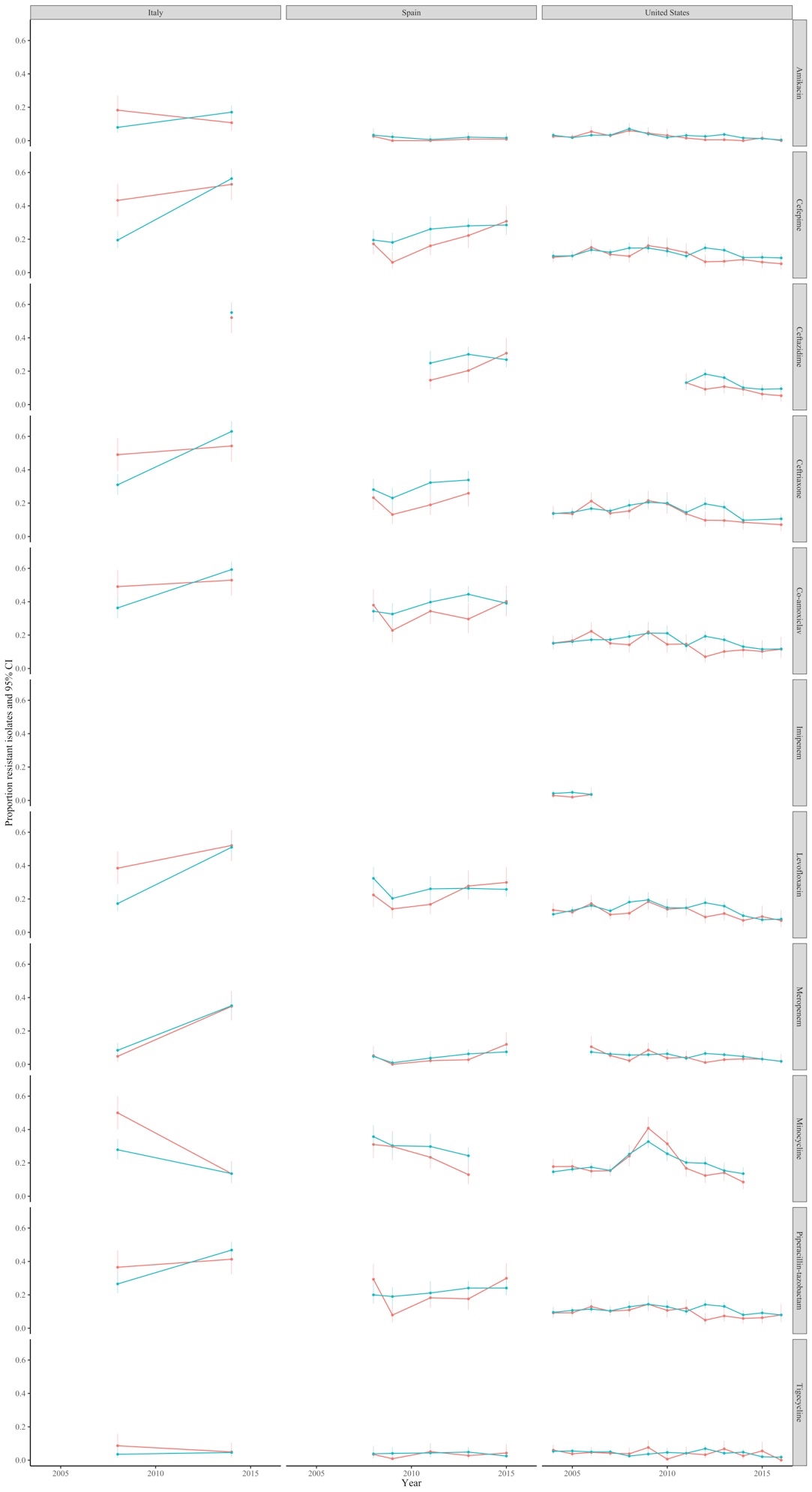

Supplementary Figure 13 Resistance prevalence in Klebsiella pneumoniae in blood and non-blood cultures in middle-income countries present in the ATLAS dataset

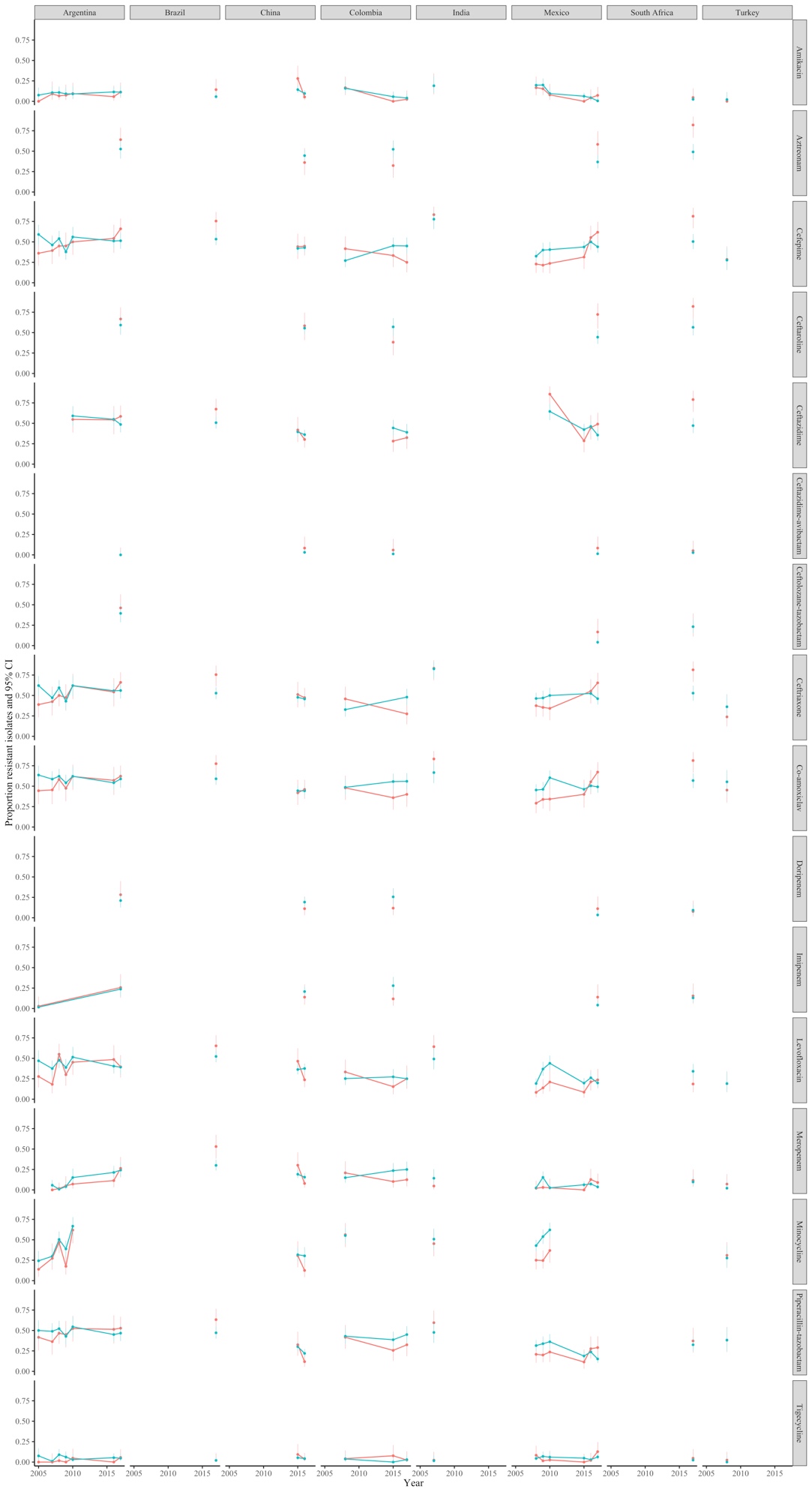

Supplementary Figure 14 Resistance prevalence in Pseudomonas aeruginosa in blood and non-blood cultures in high-income countries present in the ATLAS dataset

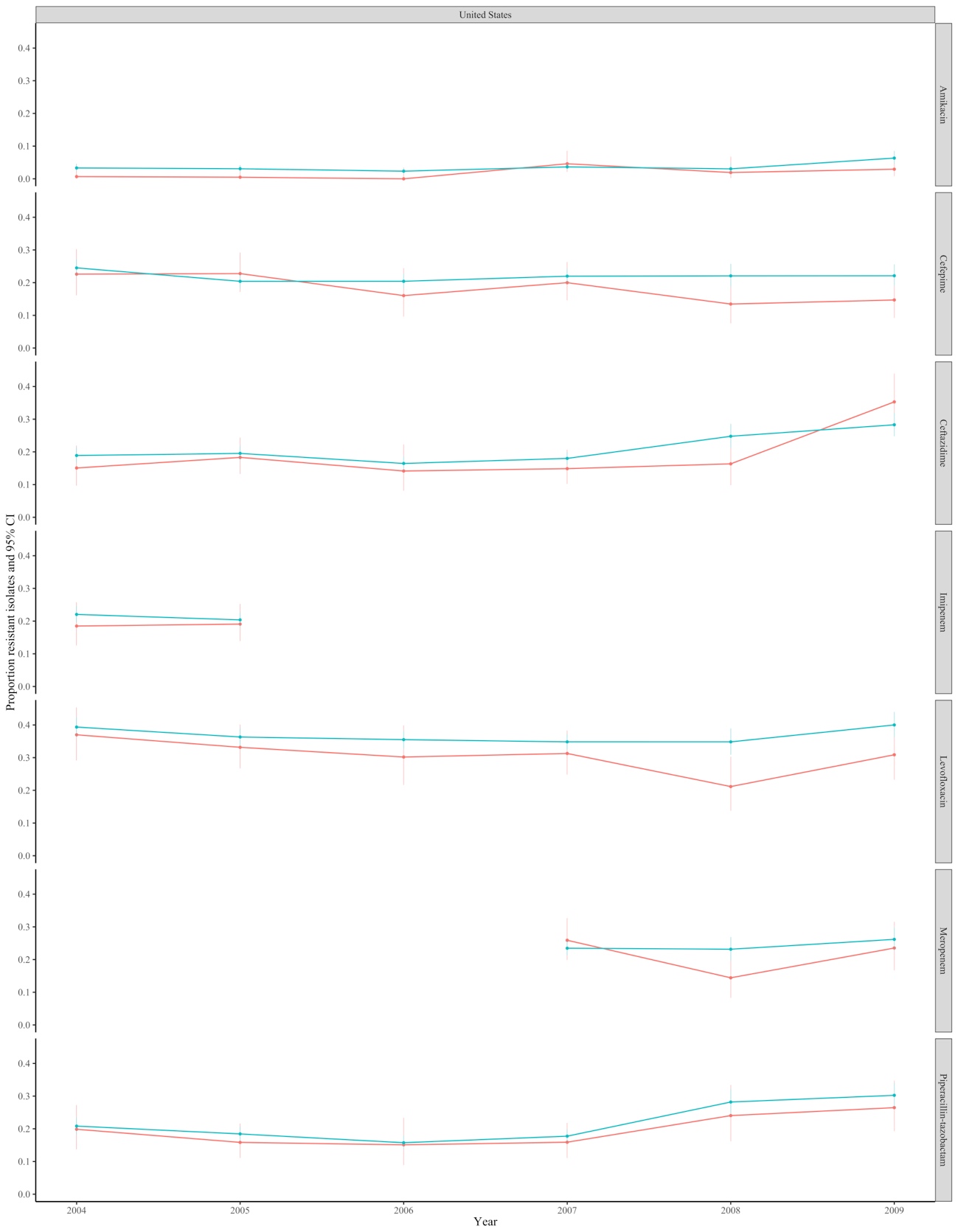

Supplementary Figure 15 Resistance prevalence in Pseudomonas aeruginosa in blood and non-blood cultures in middle-income countries present in the ATLAS dataset

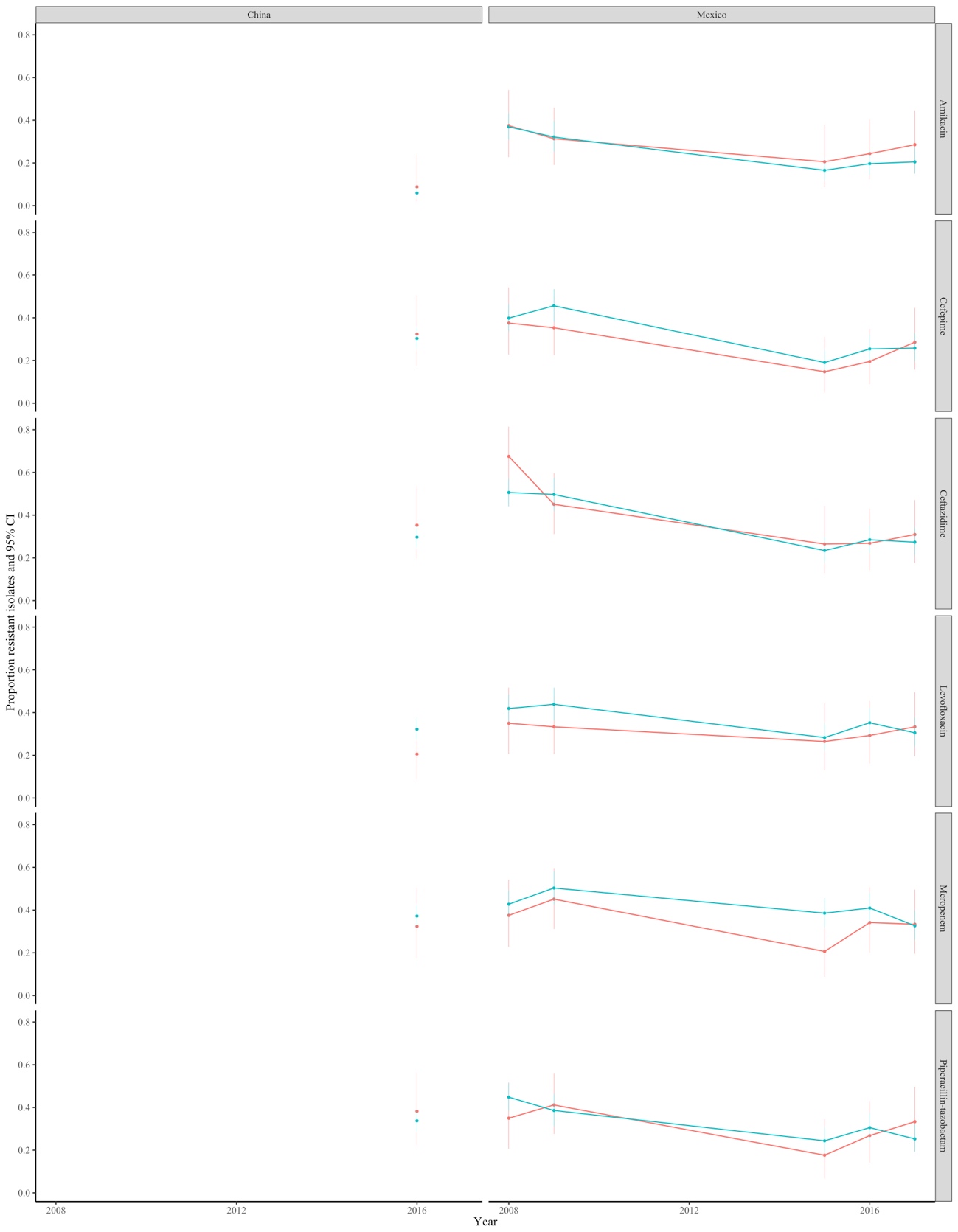

Supplementary Figure 16 Resistance in blood and non-blood cultures (A) Klebsiella pneumoniae, (B) Pseudomonas aeruginosa ATLAS dataset split into high-income countries and middle-income countries

(A) *K. pneumoniae*

High-income countries

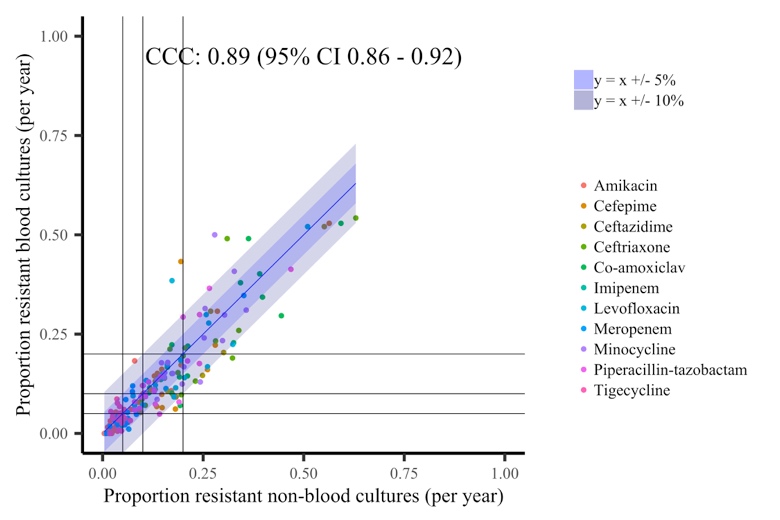

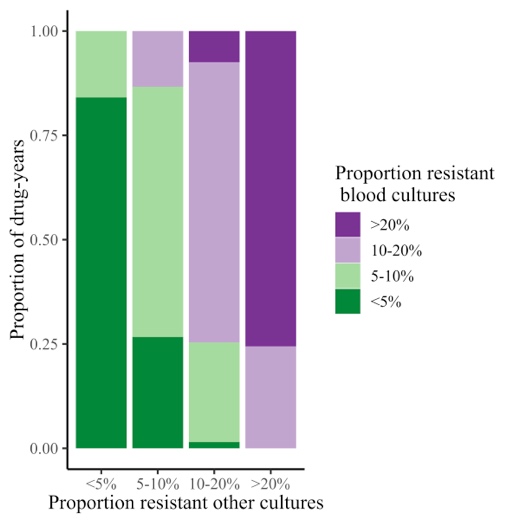

Middle-income countries

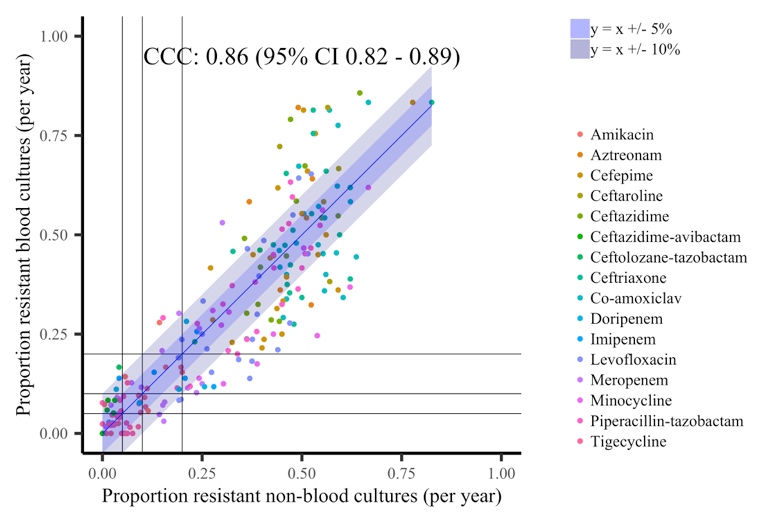

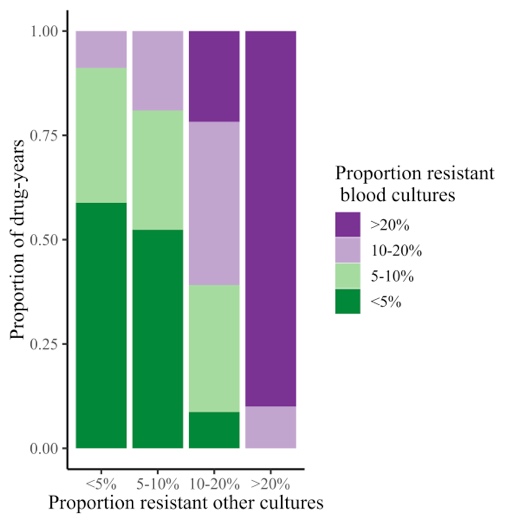

(B) *P. aeruginosa*

High-income countries

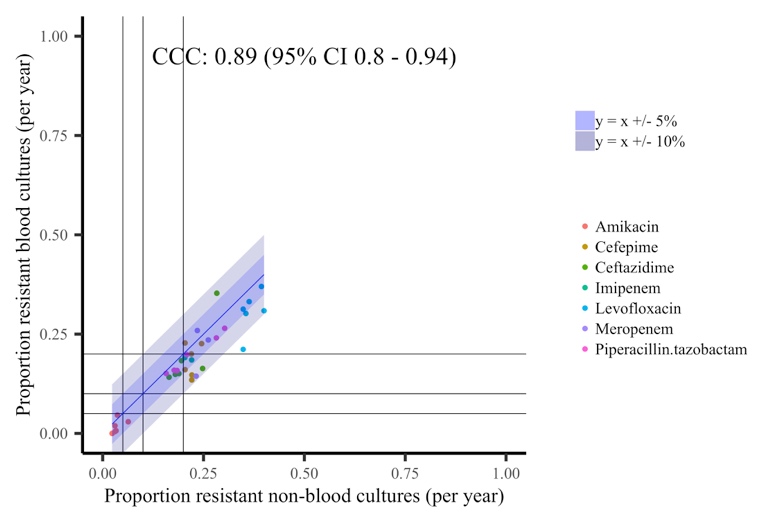

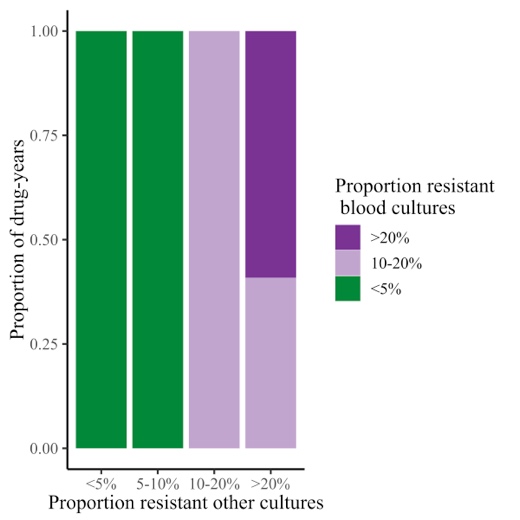

Middle-income countries

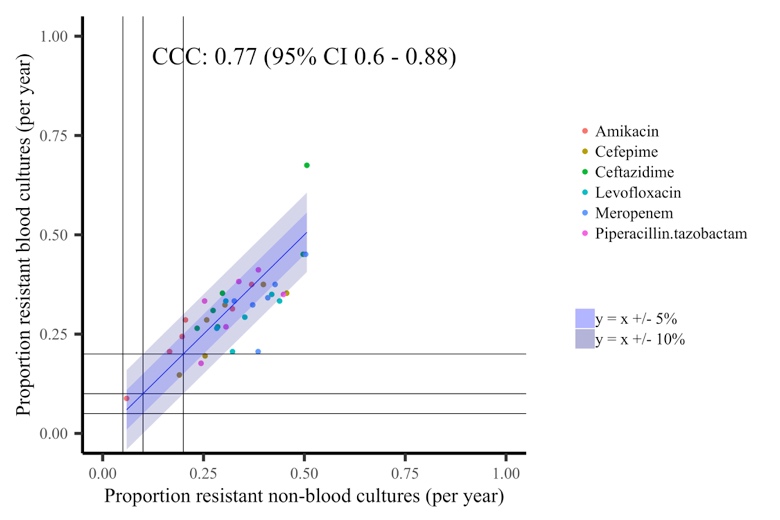

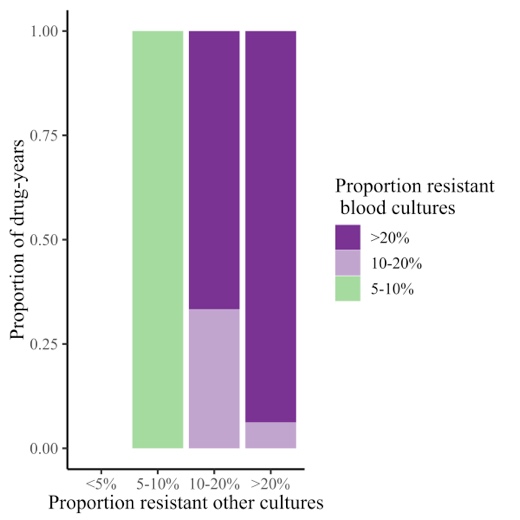

Supplementary Figure 17 Resistance prevalence in S. aureus in blood and non-blood cultures in three hospitals/programmes in LMICs

Cambodia	Laos	Thailand

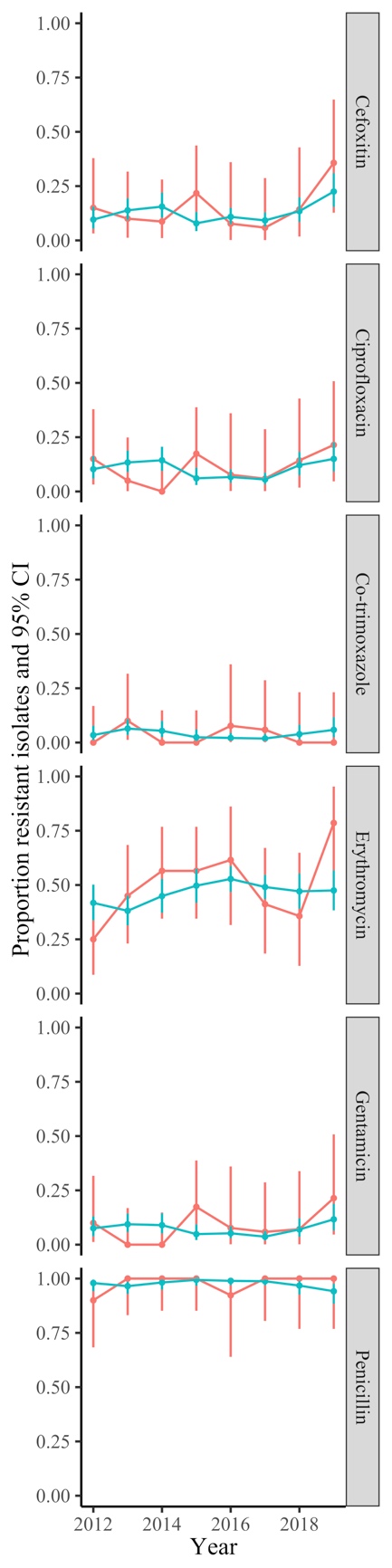

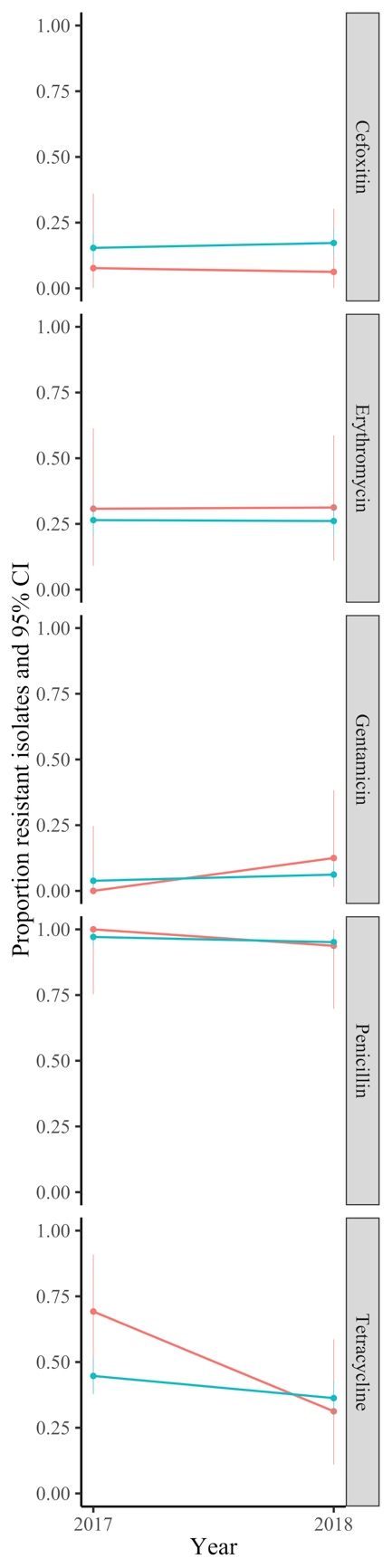

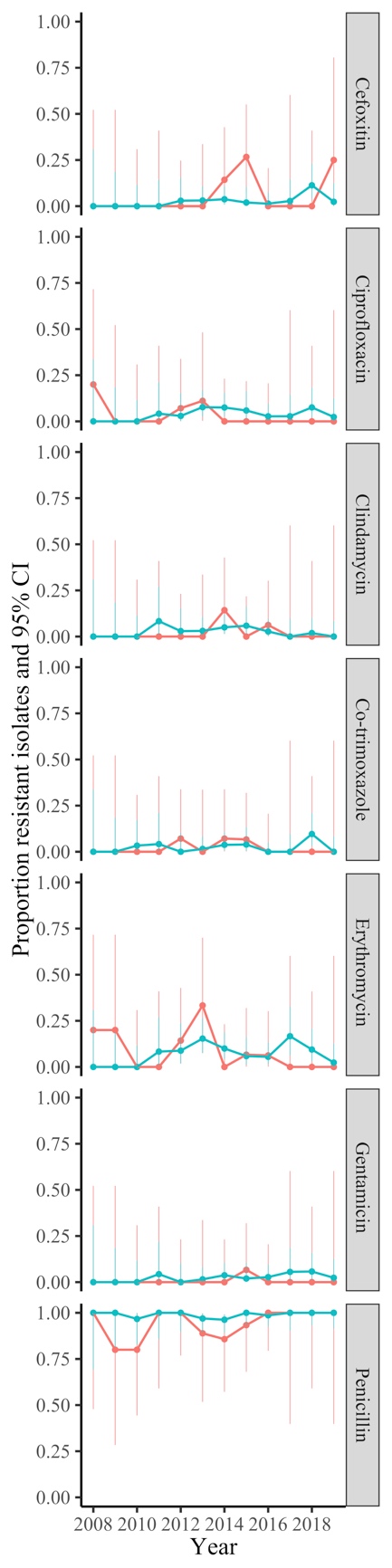

Supplementary Figure 18 Resistance in S. aureus blood and non-blood cultures in three hospitals/programmes in LMICs

(A) Cambodia

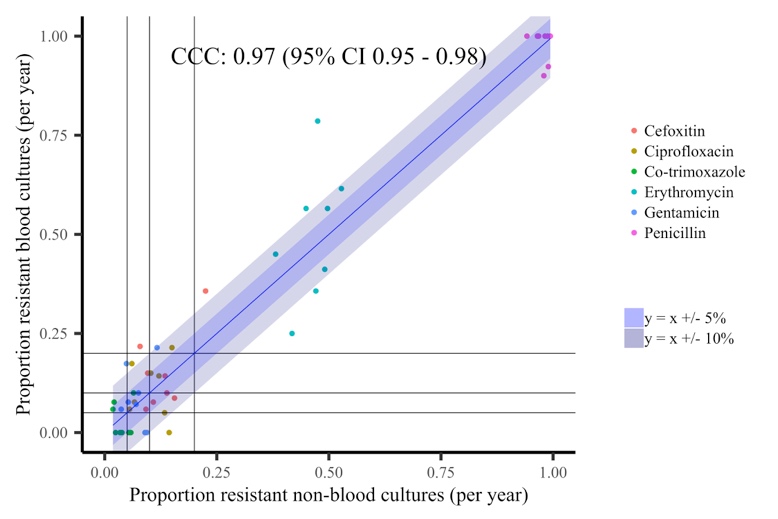

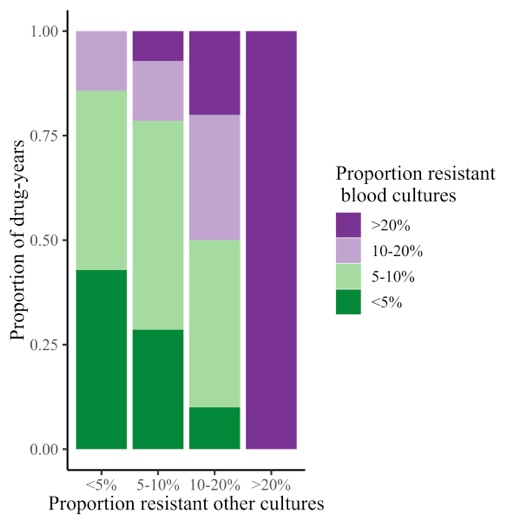

(B) Laos

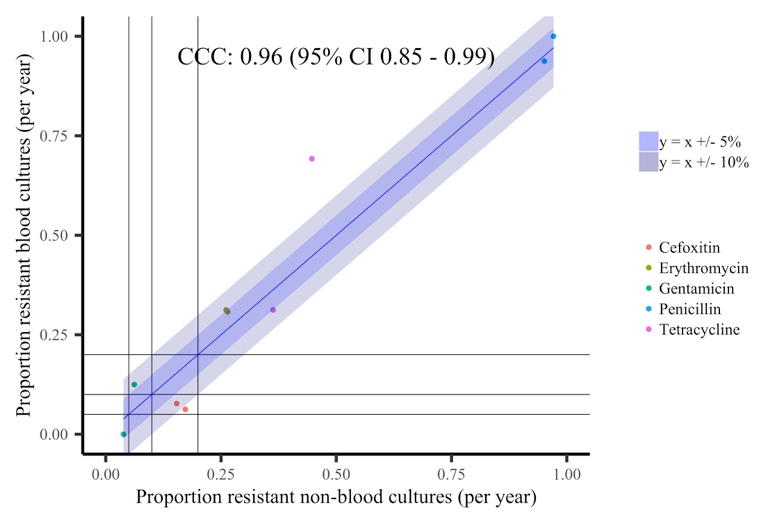

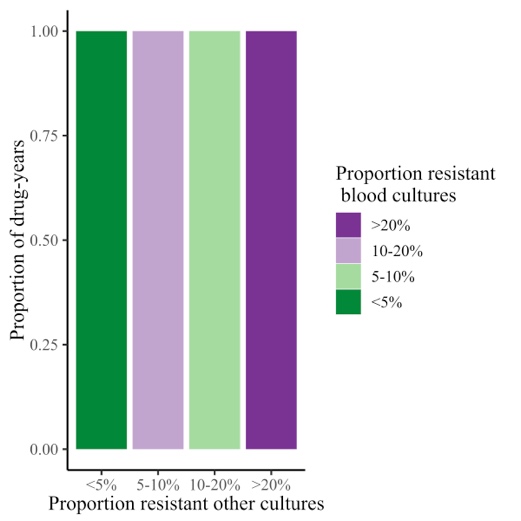

(C) Thailand

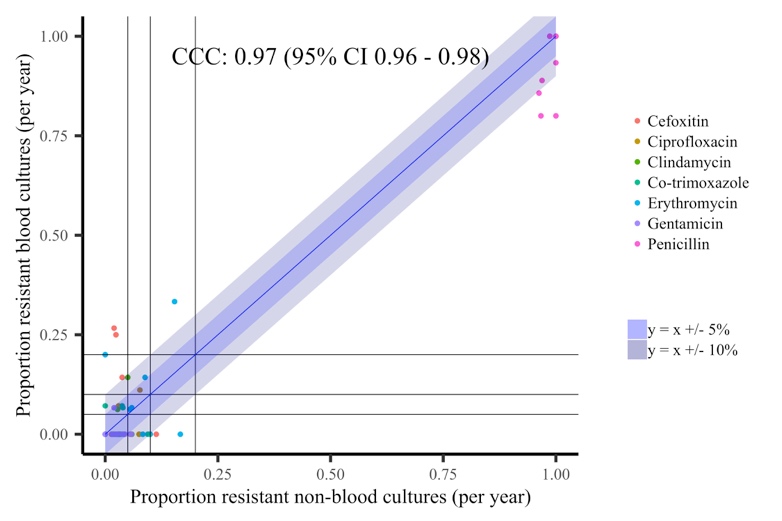

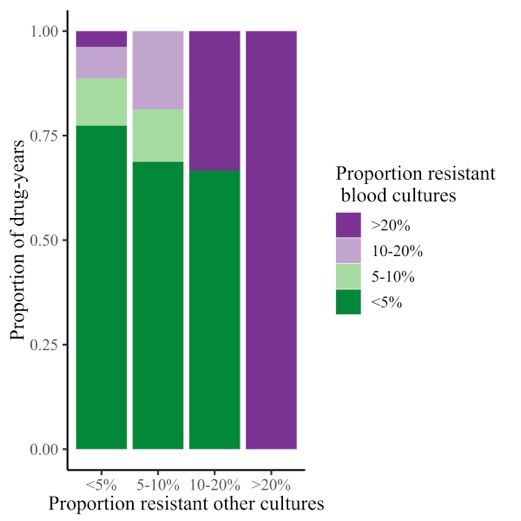

Supplementary Table 1 Summary of key results for agreement between proportions resistant in blood cultures vs other cultures, for all three datasets across all pathogens

|  | **Maximum**  **cross-correlation**  **(first column in supplementary figures)** | **Lin’s concordance coefficient (95% CI)** | **Pooled odds ratio (OR) for resistance in blood vs other samples* (95% CI)**  **(last column in supplementary figures)** | **Heterogeneity I^2^, p** | **Impact of calendar time on log OR (95% CI) p** | **Impact of log odds resistant in other samples on log odds resistant bloods (95% CI) p**  **(middle column in supplementary figures)** |
| --- | --- | --- | --- | --- | --- | --- |
| ***E. coli*** |  |  |  |  |  |  |
| Tested 1998-2018 |  |  |  |  |  |  |
| Amoxicillin | 0.77 at lag = 1 year | 0.17 (95% CI 0.06 - 0.28) | 1.55 (1.42,1.7) | 73%, p<0.0001 | 0.01 (-0.01,0.02), p=0.28 | 1.9 (1.15,2.65), p<0.0001 |
| Ciprofloxacin | 0.95 at lag = 0 year | 0.55 (95% CI 0.37 - 0.69) | 1.74 (1.59,1.9) | 37%, p=0.046 | 0.01 (-0.01,0.03), p=0.22 | 1.18 (0.97,1.39), p<0.0001 |
| Co-amoxiclav | 0.96 at lag = 0 year | 0.71 (95% CI 0.53 - 0.82) | 2.01 (1.8,2.24) | 75%, p<0.0001 | -0.01 (-0.03,0.01), p=0.30 | 0.87 (0.75,0.98), p<0.0001 |
| Trimethoprim | 0.80 at lag = 0 year | 0.77 (95% CI 0.53 - 0.9) | 0.92 (0.82,1.04) | 85%, p<0.0001 | 0.03 (0.01,0.04), p=0.0005 | 0.7 (0.46,0.95), p<0.0001 |
| ***S. aureus*** |  |  |  |  |  |  |
| Tested 1998-2018 |  |  |  |  |  |  |
| Ciprofloxacin | 0.95 at lag = 0 year | 0.81 (95% CI 0.69 - 0.89) | 1.25 (1.08,1.45) | 84%, p<0.0001 | -0.03 (-0.05,0), p=0.036 | 1.28 (1.06,1.5), p<0.0001 |
| Erythromycin | 0.93 at lag = 0 year | 0.75 (95% CI 0.61 - 0.84) | 1.15 (1.02,1.3) | 77%, p<0.0001 | -0.02 (-0.04,0), p=0.062 | 1.55 (1.27,1.84), p<0.0001 |
| Oxacillin | 0.98 at lag = 0 year | 0.73 (95% CI 0.58 - 0.83) | 1.55 (1.37,1.75) | 74%, p<0.0001 | -0.03 (-0.05,-0.01), p=0.0021 | 1.36 (1.23,1.5), p<0.0001 |
| Gentamicin | 0.54 at lag = 0 year | 0.48 (95% CI 0.12 - 0.72) | 1.71 (1.21,2.43) | 83%, p<0.0001 | -0.07 (-0.11,-0.02), p=0.0078 | 0.39 (0.13,0.64), p=0.0032 |
| Tetracycline | 0.69 at lag = -4 year | 0.13 (95% CI -0.03 - 0.29) | 0.47 (0.38,0.57) | 62%, p<0.0001 | 0.01 (-0.02,0.04), p=0.56 | 0.45 (-0.11,1.01), p=0.12 |
| Vancomycin | 0.60 at lag = 3 year | 0.02 (95% CI -0.01 - 0.04) | 10.68 (6.06,18.83) | 0%, p=1 | -0.02 (-0.11,0.08), p=0.70 | 0.27 (-0.58,1.13), p=0.53 |
| ***E. coli*** |  |  |  |  |  |  |
| Tested 2013-2018 |  |  |  |  |  |  |
| Aztreonam | 0.75 at lag = 0 year | 0.08 (95% CI -0.04 - 0.21) | 1.99 (1.61,2.46) | 62%, p=0.021 | 0.12 (0.05,0.2), p=0.0018 | 1.95 (0.39,3.51), p=0.014 |
| Ceftazidime | -0.7 at lag = -4 year | 0.02 (95% CI -0.04 - 0.08) | 2.22 (1.71,2.87) | 71%, p=0.0043 | 0.13 (0.04,0.23), p=0.0077 | 1.24 (-1.92,4.41), p=0.44 |
| Ceftriaxone | 0.72 at lag = 0 year | 0.06 (95% CI -0.04 - 0.15) | 1.88 (1.62,2.2) | 43%, p=0.12 | 0.08 (0.01,0.15), p=0.023 | 1.79 (0.11,3.46), p=0.036 |
| Co-trimoxazole | -0.8 at lag = 0 year | -0.36 (95% CI -0.78 - 0.28) | 1.13 (0.98,1.3) | 68%, p=0.0082 | 0.07 (0.02,0.13), p=0.011 | -1.23 (-2.38,-0.07), p=0.038 |
| Ertapenem | 0.81 at lag = -1 year | -0.06 (95% CI -0.56 - 0.47) | 2.68 (1.23,5.84) | 0%, p=0.44 | 0.01 (-0.51,0.53), p=0.98 | -0.07 (-1.1,0.96), p=0.90 |
| Gentamicin | 0.53 at lag = 1 year | 0.02 (95% CI -0.12 - 0.16) | 1.34 (1.03,1.73) | 73%, p=0.0021 | 0.14 (0.03,0.24), p=0.009 | 0.53 (-5.41,6.47), p=0.86 |
| Meropenem | 0.51 at lag = 0 year | 0.01 (95% CI -0.01 - 0.02) | 13.26 (5.03,34.92) | 0%, p=0.94 | 0.15 (-0.41,0.71), p=0.61 | 0.06 (-1.29,1.41), p=0.93 |
| Piperacillin-tazobactam | 0.76 at lag = -1 year | 0.09 (95% CI -0.08 - 0.25) | 2.61 (2.11,3.24) | 61%, p=0.026 | 0.06 (-0.06,0.19), p=0.33 | 0.48 (-0.06,1.02), p=0.081 |
| ***S. aureus*** |  |  |  |  |  |  |
| Tested 2013-2018 |  |  |  |  |  |  |
| Amoxicillin | 0.99 at lag = 0 year | 0.67 (95% CI 0.41 - 0.83) | 0.22 (0.06,0.75) | 42%, p=0.13 | -0.62 (-1.13,-0.12), p=0.015 | 0.27 (-0.16,0.69), p=0.22 |
| Clindamycin | 0.64 at lag = 0 year | 0.4 (95% CI -0.1 - 0.74) | 1.07 (0.88,1.3) | 41%, p=0.13 | 0.07 (-0.05,0.18), p=0.28 | 1.61 (-0.26,3.48), p=0.092 |
| Co-trimoxazole | -0.83 at lag = 0 year | -0.32 (95% CI -0.53 - -0.08) | 0.96 (0.64,1.44) | 51%, p=0.07 | -0.11 (-0.35,0.14), p=0.41 | -4.84 (-8.77,-0.91), p=0.016 |
| Daptomycin | 0.97 at lag = 0 year | 0.88 (95% CI 0.56 - 0.97) | 1.65 (0.79,3.44) | 0%, p=0.95 | 0.14 (-0.27,0.54), p=0.51 | 0.67 (-0.02,1.36), p=0.058 |
| Fusidic acid | -0.55 at lag = -1 year | 0.02 (95% CI -0.11 - 0.14) | 0.7 (0.54,0.93) | 52%, p=0.065 | 0.1 (-0.04,0.24), p=0.16 | 1.31 (-5.99,8.61), p=0.73 |
| Linezolid | 0.98 at lag = 0 year | 0.92 (95% CI 0.78 - 0.97) | 1.42 (0.67,3.02) | 0%, p=0.44 | 0.42 (-0.02,0.86), p=0.063 | 0.41 (-0.05,0.87), p=0.079 |
| Moxifloxacin | -0.7 at lag = -2 year | 0.31 (95% CI -0.19 - 0.69) | 1.38 (1.08,1.77) | 29%, p=0.22 | 0.09 (-0.04,0.21), p=0.18 | 1 (-0.32,2.33), p=0.14 |
| Mupirocin | 0.95 at lag = 0 year | 0.52 (95% CI 0.06 - 0.79) | 0.47 (0.26,0.83) | 0%, p=0.76 | -0.15 (-0.45,0.15), p=0.34 | 1.85 (0.52,3.19), p=0.0066 |
| Teicoplanin | 0.93 at lag = 0 year | 0.72 (95% CI 0.35 - 0.9) | 1.5 (0.75,2.99) | 0%, p=0.95 | 0.02 (-0.36,0.4), p=0.92 | 1.01 (0.19,1.82), p=0.015 |
| * from random effects meta-analysis treating each calendar year as an independent “study” vs urines for *E. coli and Klebsiella spp*, vs non-sterile site cultures for *S. aureus*, vs all other sites for other pathogens | | | | | | |
| ***Klebsiella* spp.** |  |  |  |  |  |  |
| Aztreonam | 0.88 at lag = 0 year | 0.32 (95% CI -0.01 - 0.58) | 1.45 (1.1,1.92) | 28%, p=0.22 | 0 (-0.18,0.18), p=0.99 | 2.13 (0.65,3.6), p=0.0047 |
| Ceftazidime | -0.53 at lag = -3 year | 0.15 (95% CI -0.17 - 0.44) | 1.47 (0.96,2.24) | 60%, p=0.03 | 0.11 (-0.13,0.36), p=0.37 | 1.14 (-1.72,4.01), p=0.43 |
| Ceftriaxone | 0.62 at lag = 0 year | 0.49 (95% CI -0.18 - 0.85) | 0.95 (0.72,1.24) | 31%, p=0.20 | 0.02 (-0.15,0.19), p=0.85 | 1.13 (-0.59,2.85), p=0.20 |
| Ciprofloxacin | 0.64 at lag = 0 year | 0.62 (95% CI 0.21 - 0.84) | 1.21 (0.92,1.59) | 51%, p=0.01 | 0.01 (-0.03,0.05), p=0.59 | 0.42 (0.07,0.77), p=0.018 |
| Co-amoxiclav | 0.89 at lag = 0 year | 0.77 (95% CI 0.58 - 0.88) | 1.13 (0.96,1.33) | 37%, p=0.069 | 0.03 (0.01,0.05), p=0.0013 | 1.36 (0.96,1.76), p<0.0001 |
| Co-trimoxazole | 0.54 at lag = 2 year | -0.1 (95% CI -0.58 - 0.44) | 1.03 (0.7,1.52) | 73%, p=0.0026 | 0.06 (-0.18,0.31), p=0.60 | -0.59 (-3.13,1.95), p=0.65 |
| Ertapenem | 0.84 at lag = -1 year | -0.04 (95% CI -0.66 - 0.61) | 2.09 (0.91,4.84) | 52%, p=0.064 | 0.23 (-0.28,0.73), p=0.38 | -0.07 (-0.81,0.66), p=0.84 |
| Gentamicin | 0.75 at lag = 0 year | 0.1 (95% CI -0.03 - 0.22) | 1.59 (0.9,2.8) | 75%, p=0.0015 | -0.04 (-0.45,0.37), p=0.84 | 4.7 (-0.99,10.4), p=0.11 |
| Meropenem | 0.67 at lag = -3 year | 0 (95% CI -0.01 - 0.01) | 16.69 (5.54,50.31) | 0%, p=1 | 0.08 (-0.58,0.74), p=0.81 | -0.14 (-5,4.71), p=0.95 |
| Piperacillin-tazobactam | -0.52 at lag = -2 year | 0.11 (95% CI -0.15 - 0.35) | 1.83 (1.45,2.31) | 29%, p=0.22 | 0.08 (-0.04,0.2), p=0.21 | 0.46 (-0.49,1.4), p=0.34 |
| ***S. pneumoniae*** |  |  |  |  |  |  |
| Amoxicillin | 0.50 at lag = 3 year | 0.15 (95% CI -0.35 - 0.58) | 0.79 (0.32,1.92) | 0%, p=1 | -0.05 (-0.58,0.47), p=0.84 | 0.22 (-3.56,4.01), p=0.91 |
| Ceftriaxone | 0.50 at lag = -2 year | 0.01 (95% CI -0.05 - 0.07) | 3.06 (1.01,9.24) | 0%, p=1 | -0.06 (-0.71,0.6), p=0.86 | 0.04 (-3.02,3.11), p=0.98 |
| Chloramphenicol | -0.64 at lag = 1 year | 0.04 (95% CI -0.15 - 0.23) | 3.38 (1.42,8.04) | 0%, p=0.47 | -0.43 (-0.89,0.03), p=0.067 | 0.08 (-1.41,1.58), p=0.91 |
| Co-trimoxazole | 0.63 at lag = -1 year | 0.06 (95% CI -0.68 - 0.74) | 0.96 (0.54,1.7) | 53%, p=0.06 | -0.23 (-0.52,0.07), p=0.13 | -0.02 (-1.39,1.35), p=0.98 |
| Erythromycin | 0.74 at lag = 4 year | 0.07 (95% CI -0.36 - 0.48) | 0.69 (0.49,0.97) | 30%, p=0.21 | -0.21 (-0.38,-0.04), p=0.015 | 0.52 (-1.95,2.98), p=0.68 |
| Linezolid | 0.84 at lag = 0 year | 0.04 (95% CI -0.02 - 0.11) | 3.63 (1.17,11.31) | 0%, p=1 | 0.02 (-0.64,0.69), p=0.95 | 0.63 (-2.79,4.05), p=0.72 |
| Meropenem | 0.85 at lag = 0 year | 0.04 (95% CI -0.02 - 0.09) | 3.54 (1.14,11.04) | 0%, p=1 | 0.01 (-0.65,0.68), p=0.97 | 0.71 (-3.15,4.57), p=0.72 |
| Moxifloxacin | 0.75 at lag = 0 year | 0.11 (95% CI -0.04 - 0.25) | 3.28 (1.88,5.71) | 5%, p=0.38 | 0.06 (-0.3,0.42), p=0.75 | 2.29 (0.05,4.53), p=0.045 |
| Teicoplanin | 0.86 at lag = 0 year | 0.04 (95% CI -0.02 - 0.1) | 3.71 (1.19,11.57) | 0%, p=1 | 0.01 (-0.65,0.68), p=0.97 | 0.72 (-2.95,4.39), p=0.70 |
| Tetracycline | 0.65 at lag = 3 year | 0.53 (95% CI 0.19 - 0.75) | 0.76 (0.59,0.98) | 12%, p=0.30 | -0.03 (-0.07,0.02), p=0.27 | 0.63 (0.24,1.03), p=0.0015 |
| Vancomycin | 0.70 at lag = 0 year | 0.02 (95% CI -0.01 - 0.05) | 3.5 (1.12,10.91) | 0%, p=1 | 0.04 (-0.63,0.7), p=0.91 | 0.43 (-3.69,4.55), p=0.84 |
| ***E. faecalis*** |  |  |  |  |  |  |
| Amoxicillin | 0.85 at lag = 2 year | -0.02 (95% CI -0.06 - 0.03) | 3.62 (1.56,8.42) | 0%, p=0.99 | 0.01 (-0.49,0.51), p=0.97 | -0.16 (-3.06,2.75), p=0.92 |
| Daptomycin | 0.77 at lag = 0 year | 0.77 (95% CI 0.04 - 0.96) | 1.1 (0.58,2.1) | 0%, p=0.65 | 0.15 (-0.21,0.51), p=0.42 | 0.52 (-0.31,1.36), p=0.22 |
| Gentamicin | 0.40 at lag = -4 year | 0.04 (95% CI -0.55 - 0.61) | 1.24 (0.89,1.73) | 24%, p=0.25 | 0.1 (-0.1,0.29), p=0.34 | 0 (-1.58,1.59), p=1 |
| Linezolid | 0.76 at lag = 3 year | 0 (95% CI -0.02 - 0.01) | 9.46 (3.83,23.35) | 0%, p=0.97 | -0.06 (-0.6,0.49), p=0.83 | -0.09 (-2.2,2.02), p=0.93 |
| Teicoplanin | -0.36 at lag = 0 year | -0.04 (95% CI -0.17 - 0.08) | 3.82 (1.83,7.98) | 0%, p=0.84 | 0.29 (-0.15,0.72), p=0.19 | -0.43 (-2.53,1.68), p=0.69 |
| Vancomycin | -0.48 at lag = -2 year | -0.05 (95% CI -0.16 - 0.07) | 4.29 (2.14,8.61) | 0%, p=0.77 | 0.33 (-0.1,0.75), p=0.13 | -0.5 (-2.58,1.57), p=0.63 |
| ***E. faecium*** |  |  |  |  |  |  |
| Amoxicillin | 0.51 at lag = 0 year | 0.46 (95% CI -0.36 - 0.88) | 1.12 (0.78,1.61) | 0%, p=0.80 | 0.05 (-0.16,0.26), p=0.67 | 0.72 (-0.83,2.26), p=0.36 |
| Vancomycin | -0.45 at lag = -3 year | 0.18 (95% CI -0.25 - 0.55) | 1.54 (1.04,2.3) | 72%, p=0.003 | -0.26 (-0.4,-0.13), p=0.0001 | 1.07 (-0.89,3.04), p=0.28 |

* from random effects meta-analysis treating each calendar year as an independent “study” vs urines for *E. coli and Klebsiella spp*, vs non-sterile site cultures for *S. aureus*, vs all other sites for other pathogens

Supplementary Table 2 Impact of log odds resistant in other samples on log odds resistant bloods, for all three datasets across all pathogens

| **Pathogen** | **Impact of log odds resistant in other samples on log odds resistant bloods (95% CI) p** | **Drug-years outside credible interval*** |
| --- | --- | --- |
| **Oxfordshire** | | |
| ***E. coli*** | +0.82 (+0.78,+0.86), p<0.0001 | 0/132 (0%) |
| ***Klebsiella* spp*.*** | +0.62 (+0.51,+0.72), p<0.0001 | 0/80 (0%) |
| ***S. aureus*** | +0.77 (+0.72,+0.82), p<0.0001 | 0/180 (0%) |
| ***S. pneumoniae*** | +0.55 (+0.44,+0.66), p<0.0001 | 0/81 (0%) |
| ***E. faecalis*** | +0.61 (+0.48,+0.73), p<0.0001 | 0/36 (0%) |
| ***E. faecium*** | +0.88 (+0.69,+1.07), p<0.0001 | 0/12 (0%) |
| **ATLAS high-income countries** | | |
| ***E. coli*** | +0.93 (+0.9,+0.96), p<0.0001 | 4/401 (1%) |
| ***S. aureus*** | +0.82 (+0.79,+0.85), p<0.0001 | 0/155 (0%) |
| ***K. pneumoniae*** | +0.95 (+0.89,+1.02), p<0.0001 | 3/186 (2%) |
| ***P. aeruginosa*** | +1 (+0.85,+1.15), p<0.0001 | 0/35 (0%) |
| **ATLAS middle-income countries** | | |
| ***E. coli*** | +0.83 (+0.79,+0.87), p<0.0001 | 1/268 (0%) |
| ***S. aureus*** | +0.72 (+0.69,+0.76), p<0.0001 | 5/344 (1%) |
| ***K. pneumoniae*** | +0.86 (+0.79,+0.93), p<0.0001 | 0/228 (0%) |
| ***P. aeruginosa*** | +0.74 (+0.49,+0.98), p<0.0001 | 1/36 (3%) |
| ***E. cloacae*** | +0.94 (+0.85,+1.03), p<0.0001 | 0/61 (0%) |
| **Cambodia** | | |
| ***E. coli*** | +0.67 (+0.52,+0.81), p<0.0001 | 0/48 (0%) |
| ***S. aureus*** | +0.75 (+0.65,+0.86), p<0.0001 | 0/48 (0%) |
| **Laos** | | |
| ***E. coli*** | +0.90 (+0.66,+1.13), p<0.0001 | 0/18 (0%) |
| ***S. aureus*** | +0.86 (+0.57,+1.15), p<0.0001 | 0/10 (0%) |
| **Thailand** | | |
| ***E. coli*** | +0.70 (+0.62,+0.77), p<0.0001 | 1/108 (1%) |
| ***S. aureus*** | +0.57 (+0.48,+0.66), p<0.0001 | 0/84 (0%) |

*no overlap between exact 95% confidence interval of resistance prevalence in bloodstream infections and credible interval

Supplementary Table 3 Summary of numbers included in analysis of the ATLAS dataset for E. coli and S. aureus in high-income countries and middle-income countries

| **Pathogen** | **Countries** | **Country-drug-year** | **Blood cultures** | | | | **Non-blood cultures** | | | |
| --- | --- | --- | --- | --- | --- | --- | --- | --- | --- | --- |
|  | | | **Total** | **Minimum** | **Mean** | **Maximum** | **Total** | **Minimum** | **Mean** | **Maximum** |
| ***E. coli*** | **High income** | 401 (>100) | 86149 | 101 | 215 | 670 | 208734 | 149 | 520 | 1496 |
|  | **Middle income** | 293 (>30) | 14582 | 30 | 50 | 99 | 45574 | 43 | 155 | 305 |
| ***S. aureus*** | **High income** | 155 (>100) | 27947 | 101 | 180 | 457 | 150178 | 248 | 743 | 2226 |
|  | **Middle income** | 350 (>30) | 17717 | 30 | 51 | 94 | 87019 | 42 | 249 | 684 |

Supplementary Table 4 Summary of numbers from the three hospitals/programmes from LMICs for E. coli and S. aureus

| **Country** | **Period** | **Pathogen** | **Blood cultures/year**  **min-max** | **Non-blood cultures/year**  **min-max** |
| --- | --- | --- | --- | --- |
| **Cambodia** | **2012-2018** | ***E. coli*** | 7-19 | 71-127 |
|  |  | ***S. aureus*** | 14-23 | 149-327 |
| **Laos** | ***2017-2018*** | ***E. coli*** | 73-82 | 82-83 |
|  |  | ***S. aureus*** | 13-16 | 208-226 |
| **Thailand** | ***2008-2019*** | ***E. coli*** | 6-30 | 72-268 |
|  |  | ***S. aureus*** | 5-16 | 15-80 |

Supplementary Table 5 Within patient comparisons of susceptibility results in other sites of infections as a marker for bloodstream infections, Oxfordshire 1998-2018

|  | **Susceptible bloodstream infections, numbers** | | **Resistant bloodstream infections, numbers** | | **Major error**  **(susceptible blood, resistant other)** | **Very major error (resistant blood, susceptible other)** | **McNemar p (test for symmetry in discordance)** | **Agreement (same result in blood and other sites)** | **Positive predictive value (% resistant other site resistant in blood)** | **Negative predictive value (% susceptible other site susceptible in blood)** |
| --- | --- | --- | --- | --- | --- | --- | --- | --- | --- | --- |
|  | Susceptible  other site | Resistant  other site | Susceptible  other site | Resistant  other site |  |  |  |  |  |  |
| ***Concurrent E. coli*** |  |  |  |  |  |  |  |  |  |  |
| Tested 1998-2018 |  |  |  |  |  |  |  |  |  |  |
| Amoxicillin | 640 | 55 | 84 | 1071 | 8% | 7% | p=0.018 | 92% | 95% | 88% |
| Co-amoxiclav | 1301 | 71 | 129 | 372 | 5% | 26% | p<0.0001 | 89% | 84% | 91% |
| Ciprofloxacin | 1600 | 11 | 6 | 250 | 1% | 2% | p=0.33 | 99% | 96% | 100% |
| Trimethoprim | 837 | 169 | 88 | 727 | 17% | 11% | p<0.0001 | 86% | 81% | 90% |
| Tested 2013-2018 |  |  |  |  |  |  |  |  |  |  |
| Aztreonam | 519 | 2 | 4 | 63 | 0% | 6% | p=0.68 | 99% | 97% | 99% |
| Ceftazidime | 642 | 5 | 8 | 102 | 1% | 7% | p=0.58 | 98% | 95% | 99% |
| Ceftriaxone | 608 | 7 | 5 | 113 | 1% | 4% | p=0.77 | 98% | 94% | 99% |
| Co-trimoxazole | 390 | 14 | 5 | 187 | 3% | 3% | p=0.066 | 97% | 93% | 99% |
| Ertapenem | 623 | 1 | 0 | 0 | 0% | NA | p=1 | 100% | 0% | 100% |
| Gentamicin | 668 | 15 | 3 | 100 | 2% | 3% | p=0.0095 | 98% | 87% | 100% |
| Piperacillin-tazobactam | 629 | 70 | 18 | 62 | 10% | 22% | p<0.0001 | 89% | 47% | 97% |
| **Previous *E. coli*** |  |  |  |  |  |  |  |  |  |  |
| Tested 1998-2018 |  |  |  |  |  |  |  |  |  |  |
| Amoxicillin | 217 | 33 | 94 | 461 | 13% | 17% | p<0.0001 | 84% | 93% | 70% |
| Co-amoxiclav | 477 | 47 | 105 | 194 | 9% | 35% | p<0.0001 | 82% | 80% | 82% |
| Ciprofloxacin | 620 | 18 | 33 | 147 | 3% | 18% | p=0.05 | 94% | 89% | 95% |
| Trimethoprim | 290 | 83 | 77 | 335 | 22% | 19% | p=0.69 | 80% | 80% | 79% |
| Tested 2013-2018 |  |  |  |  |  |  |  |  |  |  |
| Aztreonam | 242 | 13 | 14 | 36 | 5% | 28% | p=1 | 91% | 73% | 95% |
| Ceftazidime | 280 | 20 | 12 | 58 | 7% | 17% | p=0.22 | 91% | 74% | 96% |
| Ceftriaxone | 276 | 21 | 12 | 65 | 7% | 16% | p=0.16 | 91% | 76% | 96% |
| Co-trimoxazole | 165 | 15 | 26 | 99 | 8% | 21% | p=0.12 | 87% | 87% | 86% |
| Gentamicin | 306 | 18 | 9 | 68 | 6% | 12% | p=0.12 | 93% | 79% | 97% |
| Piperacillin-tazobactam | 289 | 43 | 18 | 40 | 13% | 31% | p=0.0021 | 84% | 48% | 94% |
| **Non-sterile site *S. aureus*** |  |  |  |  |  |  |  |  |  |  |
| Tested 1998-2018 |  |  |  |  |  |  |  |  |  |  |
| Ciprofloxacin | 868 | 32 | 14 | 454 | 4% | 3% | p=0.012 | 97% | 93% | 98% |
| Erythromycin | 847 | 28 | 16 | 320 | 3% | 5% | p=0.097 | 96% | 92% | 98% |
| Gentamicin | 1198 | 15 | 15 | 41 | 1% | 27% | p=1 | 98% | 73% | 99% |
| Oxacillin | 920 | 23 | 11 | 433 | 2% | 2% | p=0.059 | 98% | 95% | 99% |
| Tetracycline | 1164 | 4 | 2 | 32 | 0% | 6% | p=0.68 | 100% | 89% | 100% |
| Tested 2013-2018 |  |  |  |  |  |  |  |  |  |  |
| Ampicillin | 207 | 0 | 0 | 10 | 0% | 0% | NA | 100% | 100% | 100% |
| Clindamycin | 96 | 0 | 0 | 20 | 0% | 0% | NA | 100% | 100% | 100% |
| Co-trimoxazole | 117 | 0 | 0 | 4 | 0% | 0% | NA | 100% | 100% | 100% |
| Fusidic acid | 191 | 4 | 0 | 16 | 2% | 0% | p=0.13 | 98% | 80% | 100% |
| Linezolid | 171 | 1 | 1 | 0 | 1% | 100% | p=1 | 99% | 0% | 99% |
| Moxifloxacin | 109 | 1 | 1 | 10 | 1% | 9% | p=1 | 98% | 91% | 99% |
| Mupirocin | 142 | 2 | 1 | 1 | 1% | 50% | p=1 | 98% | 33% | 99% |
| **Non-blood site *S. aureus*** |  |  |  |  |  |  |  |  |  |  |
| Tested 1998-2018 |  |  |  |  |  |  |  |  |  |  |
| Ciprofloxacin | 1439 | 41 | 19 | 696 | 3% | 3% | p=0.0067 | 97% | 94% | 99% |
| Erythromycin | 1498 | 37 | 18 | 548 | 2% | 3% | p=0.015 | 97% | 94% | 99% |
| Gentamicin | 2065 | 16 | 24 | 60 | 1% | 29% | p=0.27 | 98% | 79% | 99% |
| Oxacillin | 1561 | 28 | 11 | 662 | 2% | 2% | p=0.01 | 98% | 96% | 99% |
| Tetracycline | 1977 | 10 | 6 | 67 | 1% | 8% | p=0.45 | 99% | 87% | 100% |
| Tested 2013-2018 |  |  |  |  |  |  |  |  |  |  |
| Amoxicillin | 0 | 0 | 0 | 296 | NA | 0% | NA | 100% | 100% | NA |
| Ampicillin | 561 | 3 | 2 | 67 | 1% | 3% | p=1 | 99% | 96% | 100% |
| Clindamycin | 283 | 2 | 1 | 43 | 1% | 2% | p=1 | 99% | 96% | 100% |
| Co-trimoxazole | 316 | 0 | 0 | 17 | 0% | 0% | NA | 100% | 100% | 100% |
| Daptomycin | 371 | 1 | 2 | 0 | 0% | 100% | p=1 | 99% | 0% | 99% |
| Fusidic acid | 560 | 3 | 2 | 41 | 1% | 5% | p=1 | 99% | 93% | 100% |
| Linezolid | 384 | 4 | 3 | 0 | 1% | 100% | p=1 | 98% | 0% | 99% |
| Moxifloxacin | 308 | 1 | 2 | 24 | 0% | 8% | p=1 | 99% | 96% | 99% |
| Mupirocin | 408 | 2 | 1 | 3 | 0% | 25% | p=1 | 99% | 60% | 100% |
